# Ageing and Decision-making: A systematic review and meta-analysis

**DOI:** 10.1101/2024.10.09.24315136

**Authors:** Nicole Ee, Brooke Brady, Craig Sinclair, Kaarin J. Anstey, Ruth Peters

## Abstract

**Objectives:** This systematic review aimed to synthesise the evidence on potential differences in financial, social, health and safety-related decision-making between younger and older adults.

**Methods:** Trial, experimental, and prospective studies including older (60+) and younger adults that reported on quantitative decision-making outcome measures (i.e., performance in relation to achieving a specific prespecified goal) were included.

**Results:** Decision-making was significantly poorer (i.e., further from prespecified goals) in older compared to younger adults (k = 57, *d*_random_ = -0.17, 95% CI -0.29, -0.04, *I^2^* = 92.92%), with high heterogeneity between studies. Age differences were observed for financial and social but not health decision-making domains.

**Discussion:** Older adults performed more ‘poorly’ on financial and social decision-making than younger adults. Reasons for observed differences may vary (e.g., different motivation and values) and require exploration in future research. This has implications for how people of different ages are supported, especially at times of important decision-making.

## Introduction

Decision-making refers to the cognitive processes that result in a final belief, choice, or course of action (Wang and Ruhe 2007). It is integral to daily life and to successful ageing. Since decision-making is influenced by multiple factors including performance in cognitive domains that are thought to change with ageing (e.g., learning, memory, information processing speed) it has been suggested that decision-making performance may change with increasing age (Del Missier et al. 2020; Lim and Yu 2015; Phillips et al. 2016). However, the degree, directionality, and implications of this remain unclear (Bruine de Bruin et al. 2012; E. Peters et al. 2007). Older age has also been associated with positive decision-making tendencies such as a reduced influence of irrelevant options on final choice (Kim and Hasher 2005) and avoidance of sunk cost bias (e.g., the tendency to continue investments with poor return) (Bruine de Bruin et al. 2007; Strough et al. 2008). Conversely, negative associations between age and decision-making have also been observed, including inconsistent application of decision rules (Bruine de Bruin et al. 2007), worse decision-making with increasing options (Besedeš et al. 2012) and susceptibility to framing effects (Bruine de Bruin et al. 2007; Finucane et al. 2005). Alongside this there is a growing awareness of the factors that might impact upon decision-making and the need to take this into account in a societal setting (Fontaine et al. 2021; Weissberger et al. 2021).

Increasing attention is also being paid to capacity assessments and the development of frameworks for assisted and supported decision-making in clinical populations (Moye et al. 2006). Understanding this potential change in decision-making with age is important in the context of our ageing populations. Building upon this knowledge will be critical to developing appropriate and well-targeted information, interventions, and policies for adults at different stages of life. Whilst the assessment of this is complex and there are multiple factors that might impact on decision-making, (Kusev et al. 2017; Lerner et al. 2015) the use of experimental decision-making tasks allows a level of standardisation in this area and the potential to examine age effects or to compare performance at different ages on the same or similar standard tasks. Furthermore, experimental decision-making studies provide important insights into the possible mechanisms underpinning age differences in cognitive task performance (Pachur et al. 2017). In particular tasks that enable proxy measurement of the mental processes underpinning real-world decision-making and instrumental activities of daily living and the assessment of decision-making under conditions simulating those that would likely accompany real-world financial, social, health and safety decisions (e.g., risk and uncertainty), given relevant goals (e.g., increasing monetary profit, social benefit). For example, the widely used Iowa Gambling Task (IGT), measures how averse or inclined individuals are to risk using a card game involving selecting cards from one of four decks.

Two of the decks either yield a penalty ($100) or high monetary reward ($250) and the other two decks are low risk ($50 deduction) low reward ($50). Participants are instructed to maximise their winnings over multiple selections. Examples of other well-known and widely used tasks include the Ultimatum game (UG), which assesses generosity and reciprocity through participants deciding how to split a monetary sum between themselves and another person and the game of dice task which requires participants to predict a dice roll through selecting high risk high reward or low risk low reward odds (full details in Online Resource: Supplementary Table VI).

Several reviews target ageing and decision-making, but few have adhered to systematic review methodology or have drawn existing data together quantitatively. Three have been systematic with the most recent published in 2015 (Best and Charness 2015; Mata et al. 2011; Thornton and Dumke 2005). Thornton and Dumke (2005) explored age differences in everyday problem solving and decision-making in 28 studies published until 2003. The authors observed a reliable deterioration of everyday problem-solving and decision-making effectiveness with older age. Mata et al. (2011) explored age differences in risky decision-making, including an exploration of framing effects. They reported no overall or framing differences between younger and older adults on risky moral and financial decision-making. More recently, Best and Charness included publications until 2013 and found younger adults were more likely to make risky decisions in positively framed (but not negatively framed) conditions (Best and Charness 2015). Moderator analysis revealed age differences were exclusive to small-reward financial and large-reward morality scenarios in positively framed conditions. Both reviews highlighted the potential impact of task characteristics on age-related decision-making. Overall, these reviews provide inconsistent evidence for age-related variability in decision-making and serve to highlight important gaps in the extant literature. These include limited generalisability of current evidence due to an underrepresentation of studies on non-risky decision-making or decisions in real-world contexts (e.g., financial decision-making, health, and safety decision-making), the need for consideration of potential moderators (such as task characteristics, demographic factors) in research design and interpretation of results, and where possible, appropriate statistical analyses accounting for these factors.

To address current knowledge gaps and improve translatability of available research, an updated, comprehensive review is required. The primary aim of this review was to synthesise the trial, experimental and prospective evidence for the impact of adult ageing on decision-making with applicability to real world outcomes and in key domains, including financial, social, health and safety-related decisions. The secondary aim of this review was to explore if task characteristics influence observed effects.

## Method

Three databases, Embase, Medline and PsycINFO, were searched from inception to 18 January 2019, using the following search terms: (exp *ageing/ or exp *cognitive ageing) AND (exp *decision-making/ OR cognitive ageing) supplemented with an additional targeted search in title, abstract and keyword fields (((ageing OR ageing) AND cognitive) AND (decision making OR decision)). Reference lists of reviews on ageing and decision-making were manually searched and field experts consulted to identify additional publications. Non-English publications were translated using the Google Translate web-based software.

### Study Selection

Two independent reviewers (NE and RP) carried out title and abstract screening for all publications returned by the searches and identified through reference lists followed by subsequent full text screening. Discrepancies in study selection were resolved through discussion and consensus at both stages.

### Eligibility Criteria and Rationale

To capture the widest possible evidence, liberal eligibility criteria were used.

Experimental, observational studies, and trials including older adults aged over 60 years and reporting on the relationship between decision-making and age were included. Studies reporting on age-related changes in adulthood (within-subject comparison) or age-related differences between a younger and older adult group (between-subjects comparison) were included. Studies of only paediatric, adolescent, animal or clinical populations were excluded. We also chose to focus on published original research and excluded editorials, commentaries, reviews, dissertation, and conference abstracts.

The literature covers a wide range of studies exploring age differences in decision-making outcomes, tendencies, and underlying mechanisms, but many do not allow for the determination of whether older age is associated with better or worse overall decisions making. While real-world decisions are often incredibly nuanced and multifaceted, in balance, better outcomes generally are those that result in better financial standing, are socially favourable e.g., more altruistic, improve health and safety, or avoid physical harm and damage or emotional distress. In these terms, this review specifically focuses on evidence relating to whether older adults are likely to be worse real-world decision-makers than their younger counter parts. To ensure applicability to real world outcomes, only studies where decisions could be quantified as positive or negative in relation to a task-specific goal were included. These primarily comprised of decision-making tasks which resulted in a final amount of points or money earned or lost. Studies exclusively investigating neutral decision response tendencies (i.e., preferencing certain options over others e.g., colour or brand preference; informational presentation type) were excluded. For example, studies investigating older adults’ car colour preferences would be excluded, as there is no way to ascertain whether choosing a blue over a red car is indicative of superior decision-making.

Conversely, studies investigating older adults’ decisions to purchase a car based on set of constraints or goals would be included, as some car choices would take them closer to the goal parameters for the task than others.

Eligible studies were included in meta-analyses if they reported effect sizes in the form of correlation coefficients (Pearson’s *r,* Spearman’s ρ), unstandardised regression coefficients (β), Cohen’s *d*, or provided sufficient data (e.g., means and standard deviations, *t* or *F* statistics, Chi-squared tests, Mann-Whitney U tests, two-way ANOVA) to calculate Cohen’s *d.* Where multiple publications were derived from the same dataset, the most comprehensive data was included to eliminate duplication.

### Data Extraction

Data were extracted by a team of four reviewers (NE, RP, CS, BB) into an *a priori* study extraction table. All the entries were then checked by a second reviewer, identified errors were corrected and checked by a third reviewer against the original publication. Data was collected on study name, publication year, sample size, mean age and standard deviation, proportion female, decision-making task and domain, type of statistical analyses, control variables, and a summary of results. If multiple age comparisons were reported, the older (aged 60+) versus younger (aged 30-60) comparisons were preferentially extracted. The classification of performance as better/poorer was based on the classification reported in the constituent studies and the prespecified goals that the studies used in their experimental decision-making tasks. Where a significant relationship between age and decision-making was reported, any secondary analyses investigating potential mechanisms were also extracted. Multiple studies within a single publication were numbered for differentiation, and publications with overlapping data sources were identified and labelled. Data on effect size were extracted. Unstandardised regression coefficients (β) (Borenstein et al. 2011) and Spearman’s rank correlations (ρ) corresponding to large sample sizes were directly substituted as *r* statistics. Imputing missing correlations with β and ρ have been found to produce relatively accurate and precise population-effects estimates, while also reducing sampling bias within meta-analyses (Peterson and Brown 2005; Rupinski and Dunlap 1996). Average effect sizes were calculated for studies reporting multiple conditions of the same task (e.g., gains and loss conditions, familiar and non-familiar conditions) to ascertain the overall age effects. Effect size direction was standardised such that a negative effect size represented poorer decision-making as a function of increasing age. Multiple rows of data were extracted if the same participants performed more than one decision-making task, and a weighted average effect size computed for tasks of the same decision-making domain. Each outcome was coded for study characteristics including decision-making tasks (Iowa Gambling Task (IGT), Ultimatum Game (UG), Game of Dice Task (GDT), Balloon Analogue Risk Task (BART), other), decision-making domain (financial, social, health, safety, other), additional task demands (altruism, risk, intertemporal judgement, inferencing, or exploration/exploitation factors), and independent age variable (categorical/continuous).

### Meta-analysis

Meta-analysis was performed with Jamovi MAJOR, with Correlations Coefficients, and variance for age effects on decision-making outcomes as the principal outcome variable. Due to variability in sampling characteristics and methodology of included studies, random effects Restricted Maximum Likelihood models were employed. Heterogeneity of outcomes was quantified with the I*^2^* index, funnel plots and summary effect size confidence intervals, with values closer to 100 per cent and a wider interval indicating greater heterogeneity, respectively (Borenstein et al. 2010; Borenstein et al. 2017; Sterne and Egger 2001) (Online Resource: Supplementary Fig. II)

Meta-analyses were carried out pooling all compatible studies, and, where possible by subgroup of decision-making domain (i.e., financial, social, health, safety). The risk of overestimation of effect sizes from small samples sizes (*N* < 50) associated with use of Cohen’s *d* was examined with post-hoc sensitivity analyses. Sensitivity analyses were performed with Jamovi MAJOR. These assessed the potential impact of moderators on the relationship between age and decision-making (publication year, mean age, task type, presence of additional task demands), whether age was measured as a categorical or continuous variable, adjustment for socio-economic factors, and effect size conversion.

### Risk of Bias Assessment

The review team developed a quality rating tool informed by the Australian National Health and Medical Research Council’s Guidelines for Assessing Risk of Bias (National Health and Medical Research Council, 2019). The quality rating tool contained 11 items which critically appraised risk of bias in five assessment domains: Selection bias (age range, sex distribution, sample size, screening), confounding bias (task administration), attrition bias, statistical analyses (appropriateness, adjustment for confounding effects), study design (suitability, clear description) (detailed in Online Resource: Supplementary Table III and IV).

Level of bias was rated low, moderate, or high for each item and overall risk of bias for the body of evidence summarised as a percentage of items rated low, moderate, or high bias per domain. A numerical scoring scheme was not employed as this can lead to a loss of subtlety when assessing quality (Jüni et al. 1999). Each study was assessed by two independent reviewers (NE, RP, CS, BB). Any discrepancies between bias ratings were resolved by discussion and consensus. The systematic review is reported in accordance with the PRISMA checklist (Online Resource: Supplementary Table V).

## Results

A total of 1591 abstracts were screened from the first search and 467 from the second, with 213 publications assessed at the full text stage (Online Resource: Supplementary Fig. I). One hundred and thirty-seven full texts were deemed ineligible for reasons provided in Online Resource: Supplementary Table I. A final 76 publications containing 87 studies met the review eligibility criteria. Thirty-eight per cent of the included publications were published within the last five years, and 75 per cent within the last decade. Sample sizes were reasonable across studies, with only 10 of the 87 studies reporting small sample sizes (*N* < 50). The mean sample ages ranged from 18.7 to 83.5, and most studies included both male and female participants, albeit in different proportions (Table I shows the characteristics of the included studies).

**Table I.**
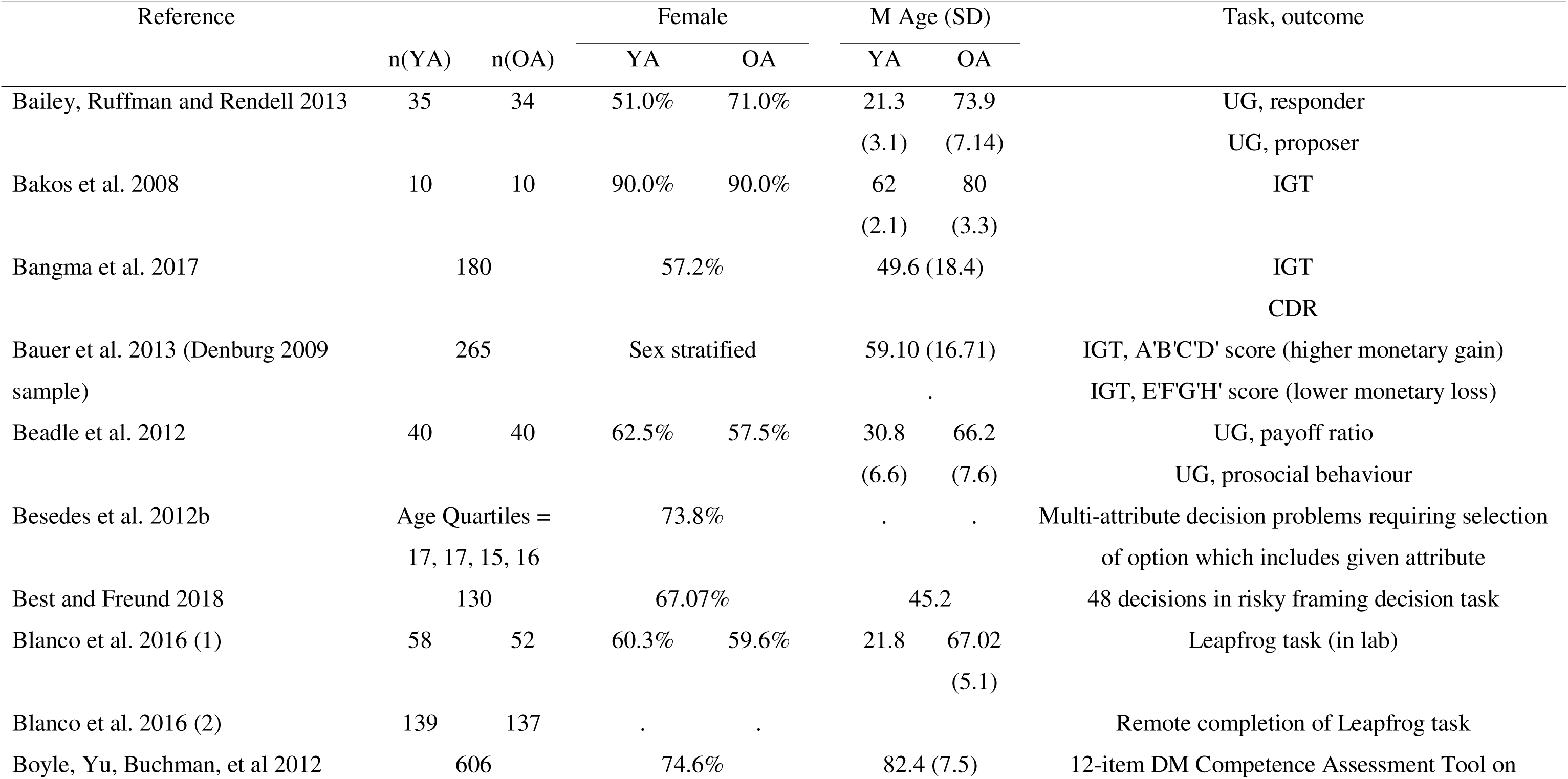
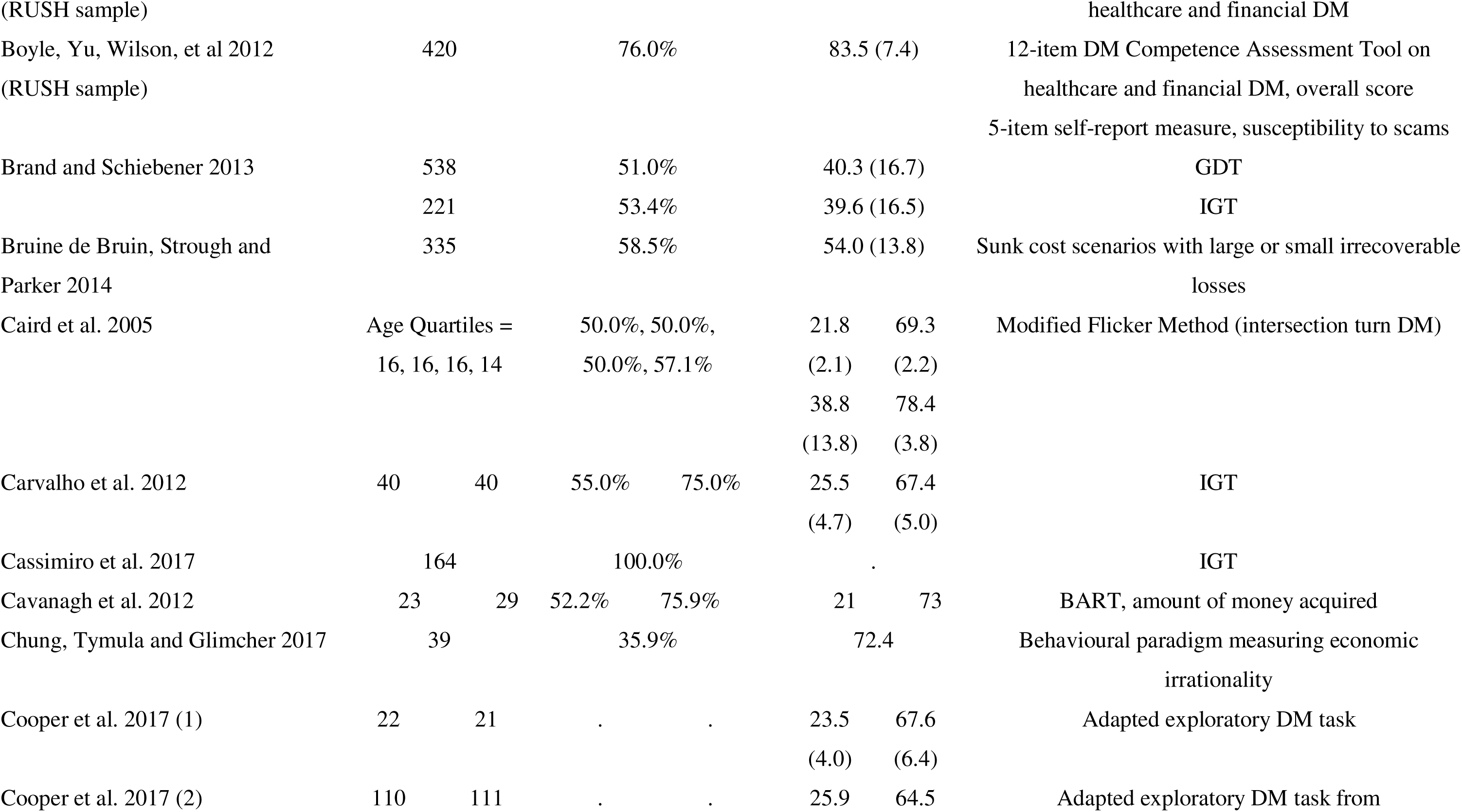
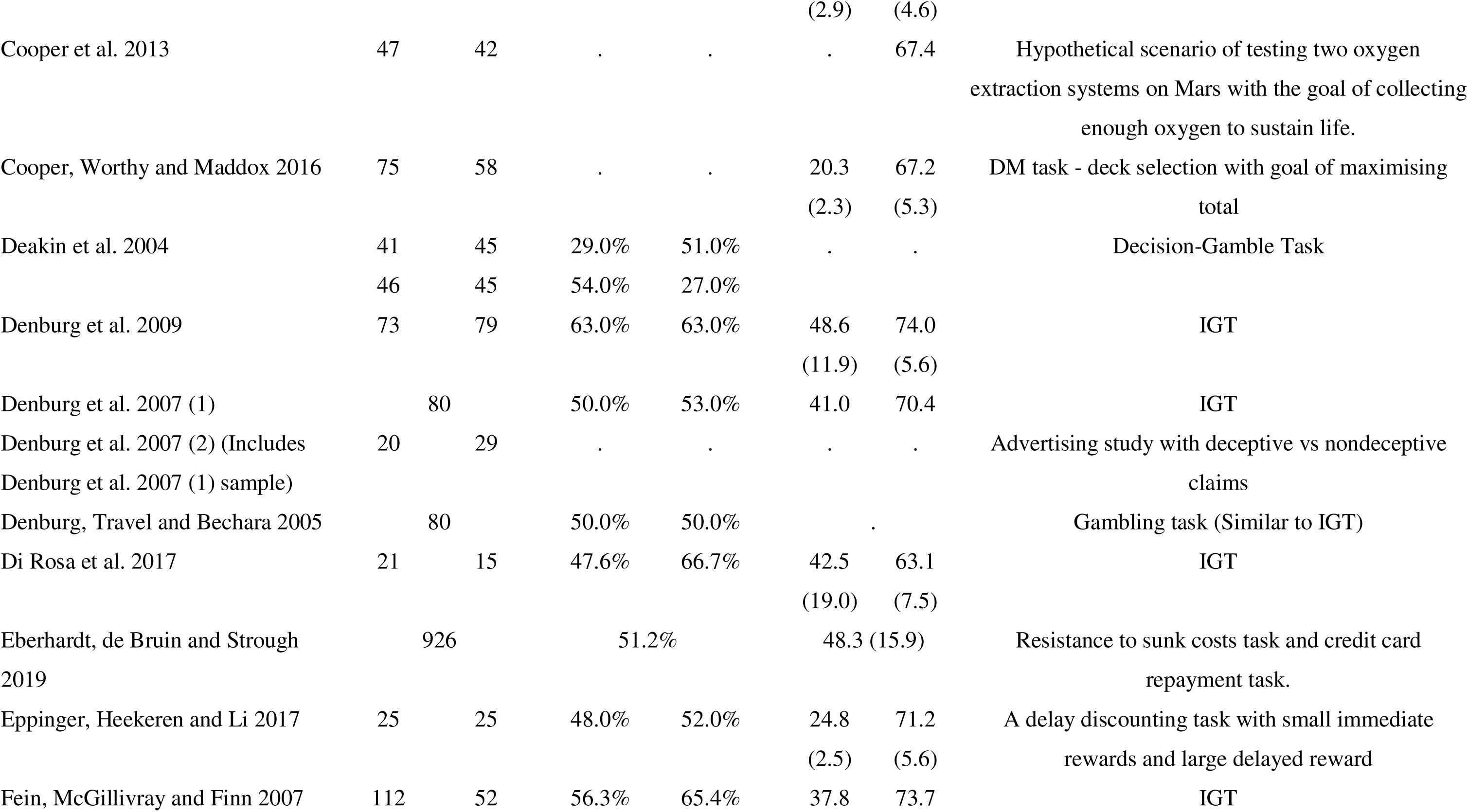
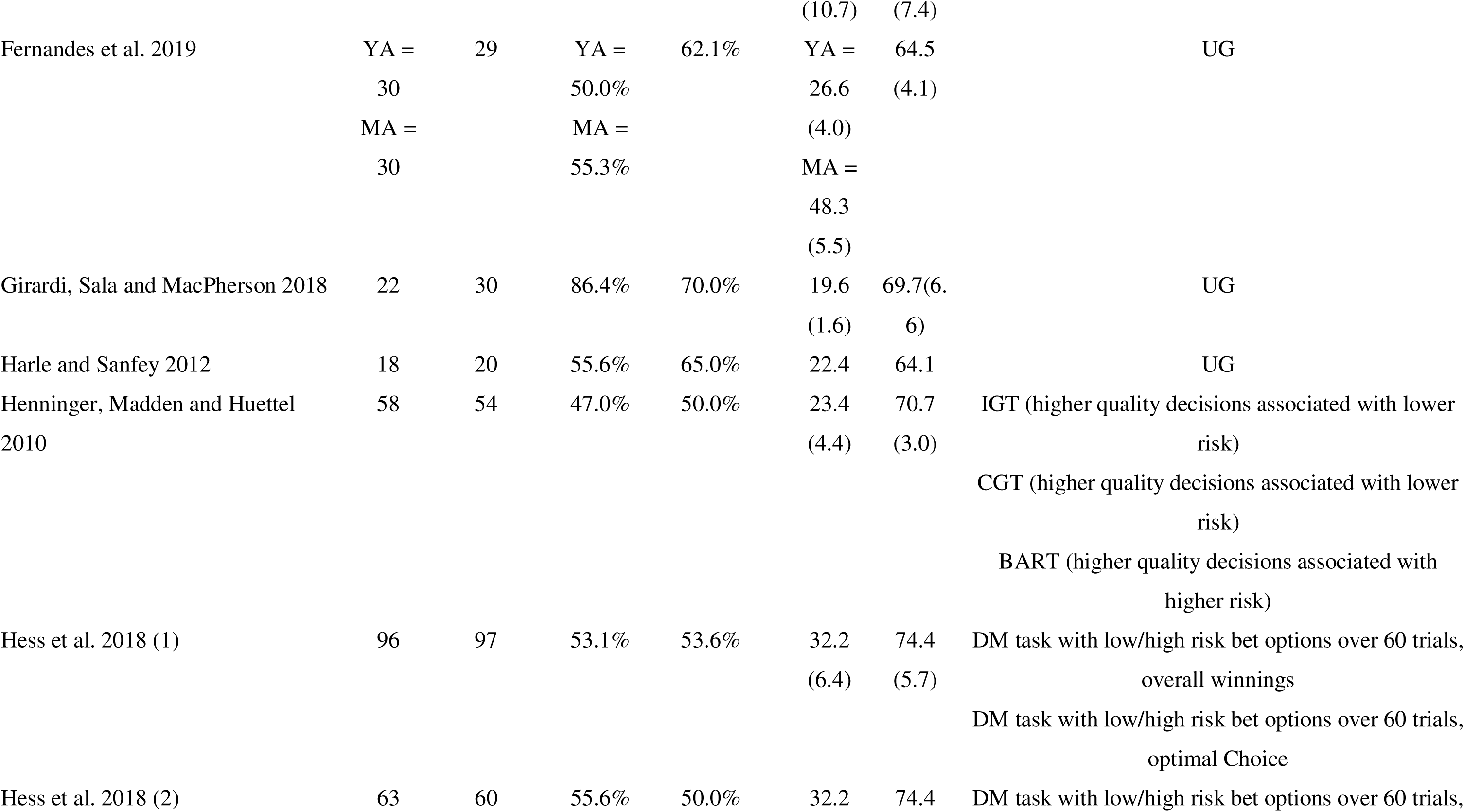
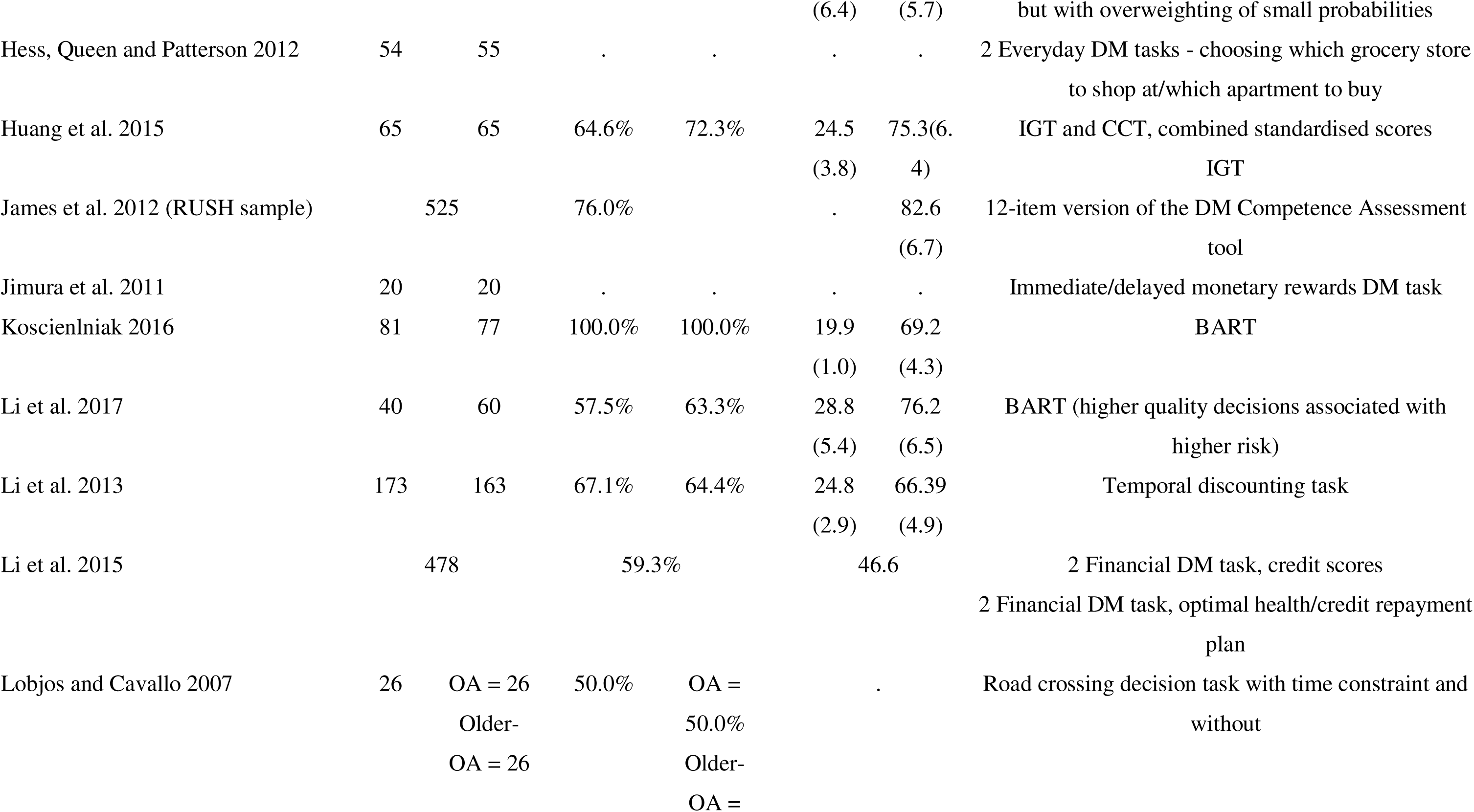
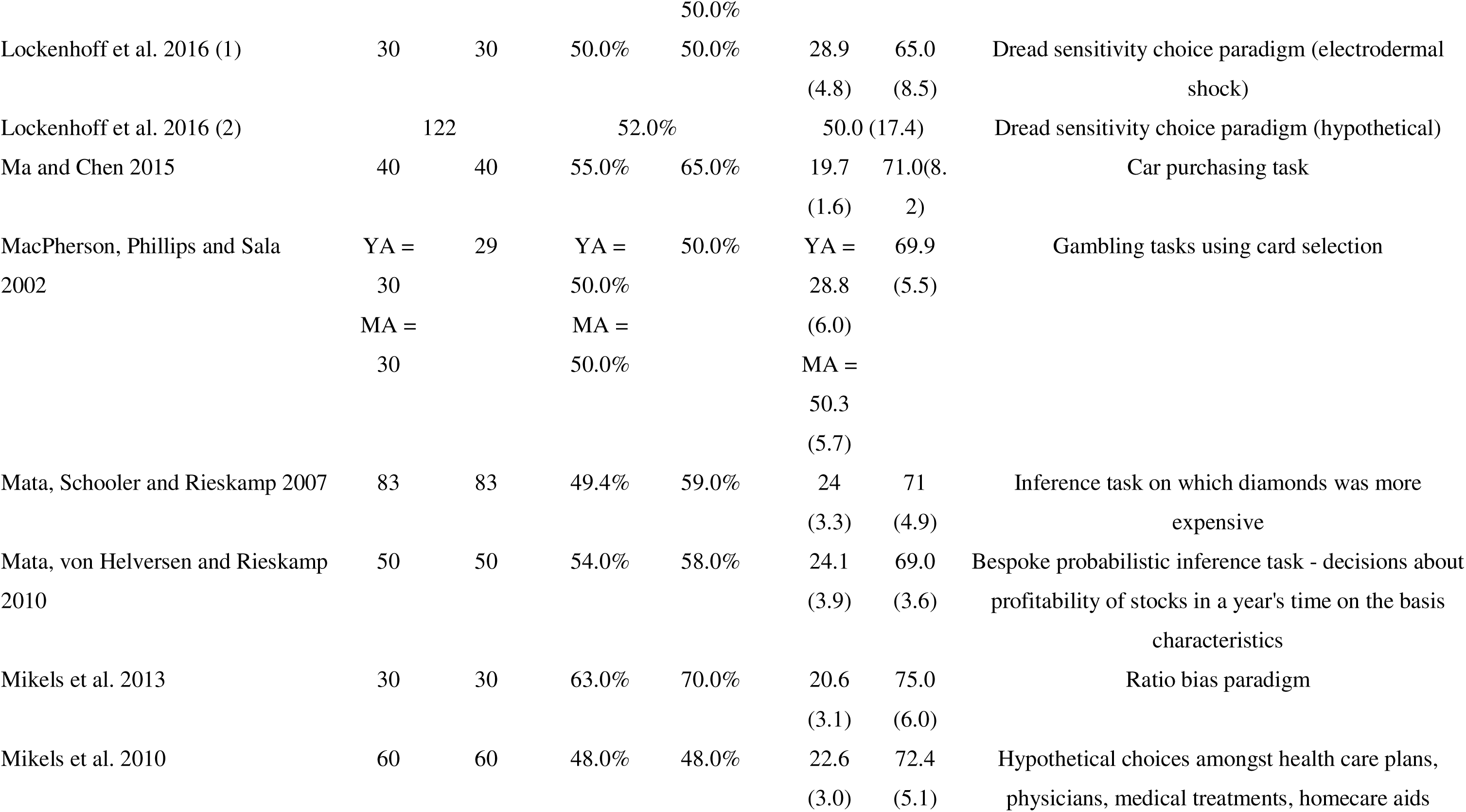
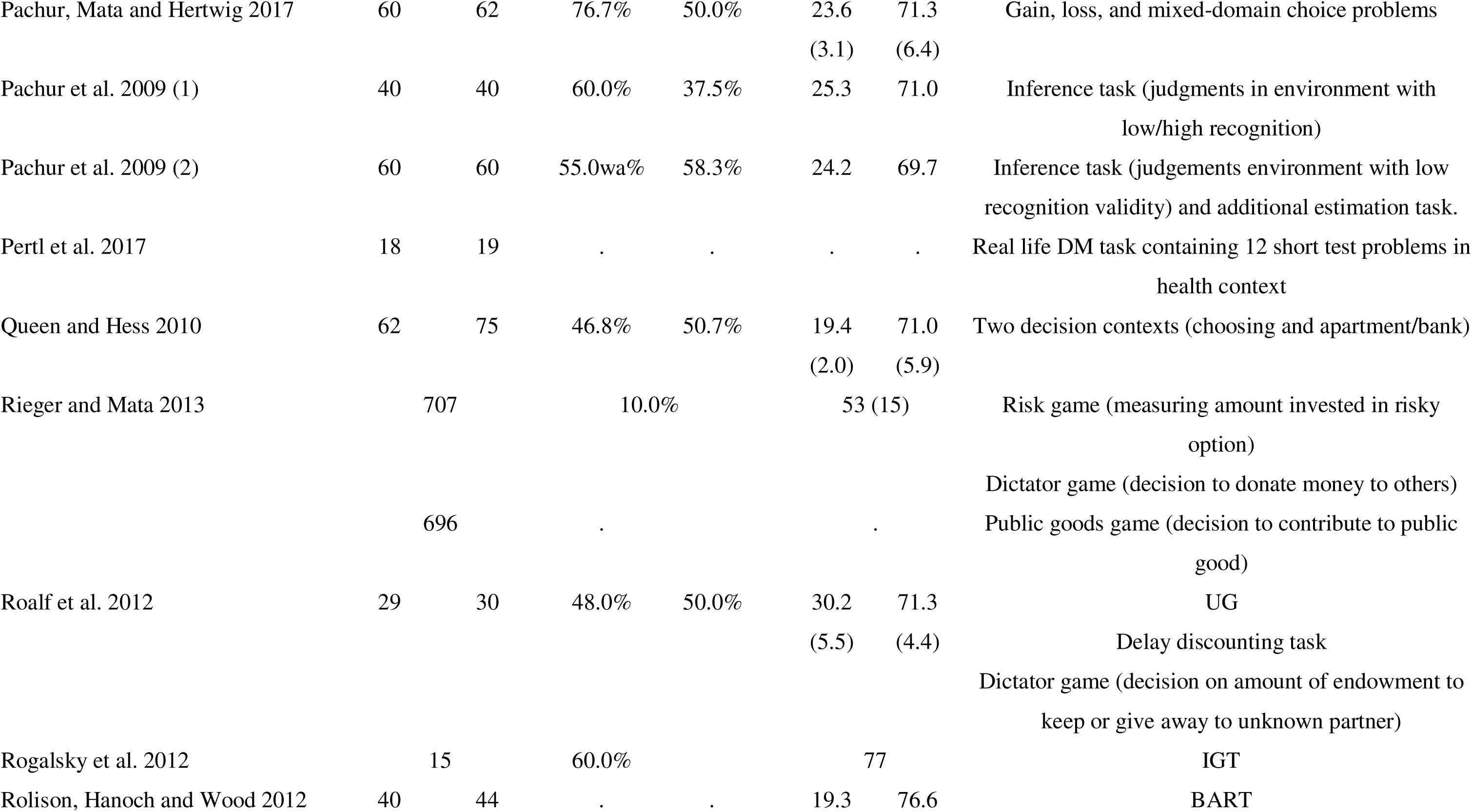
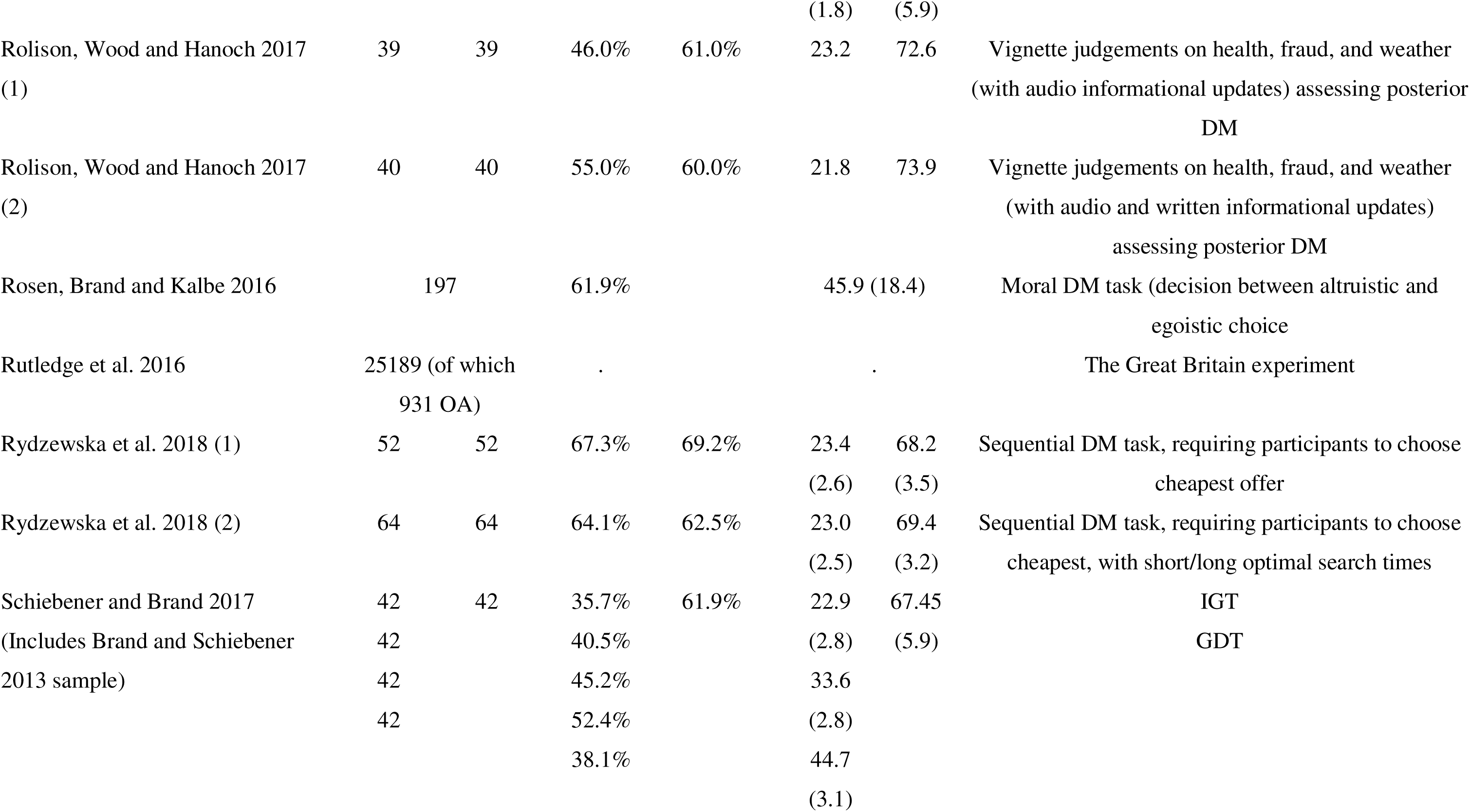
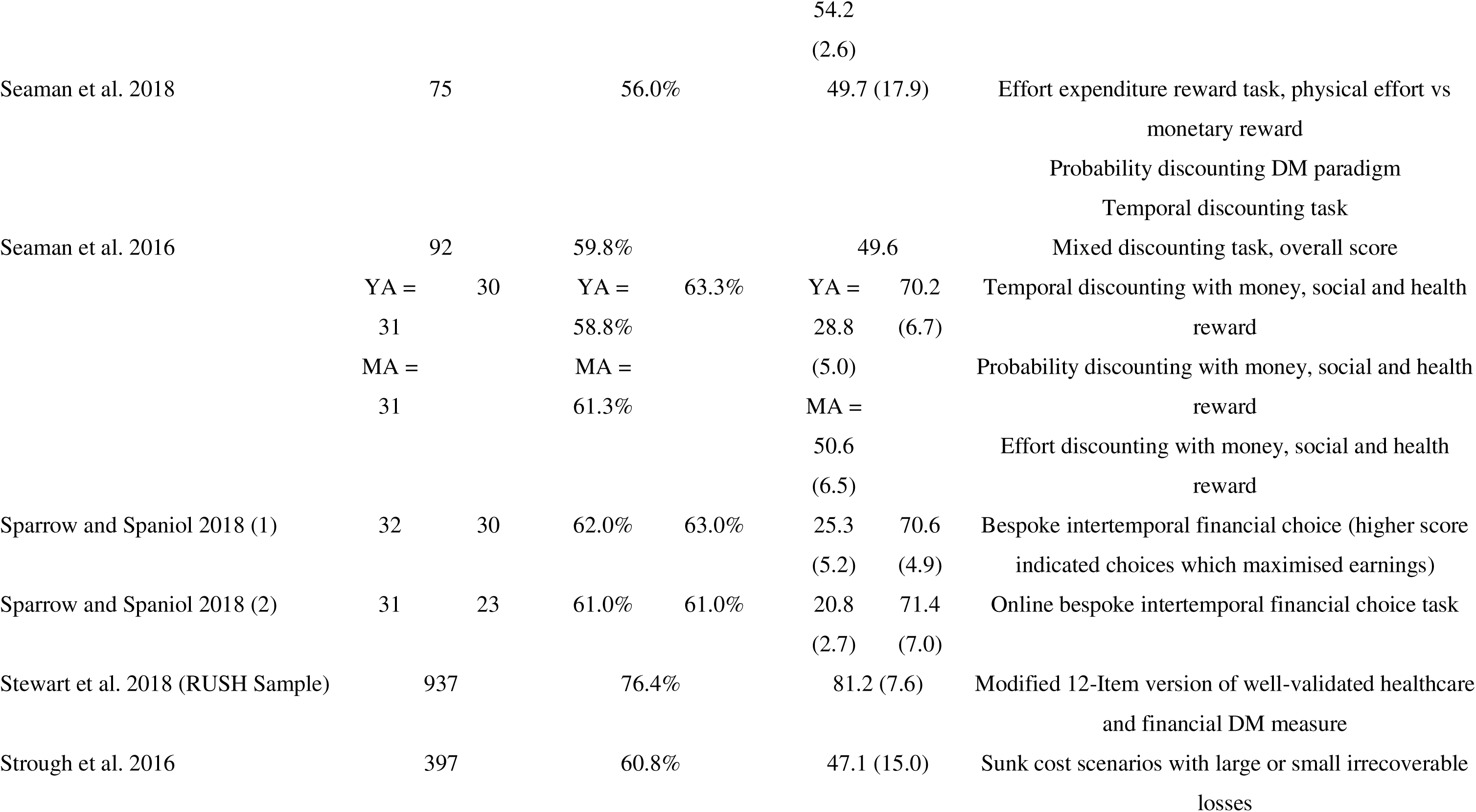
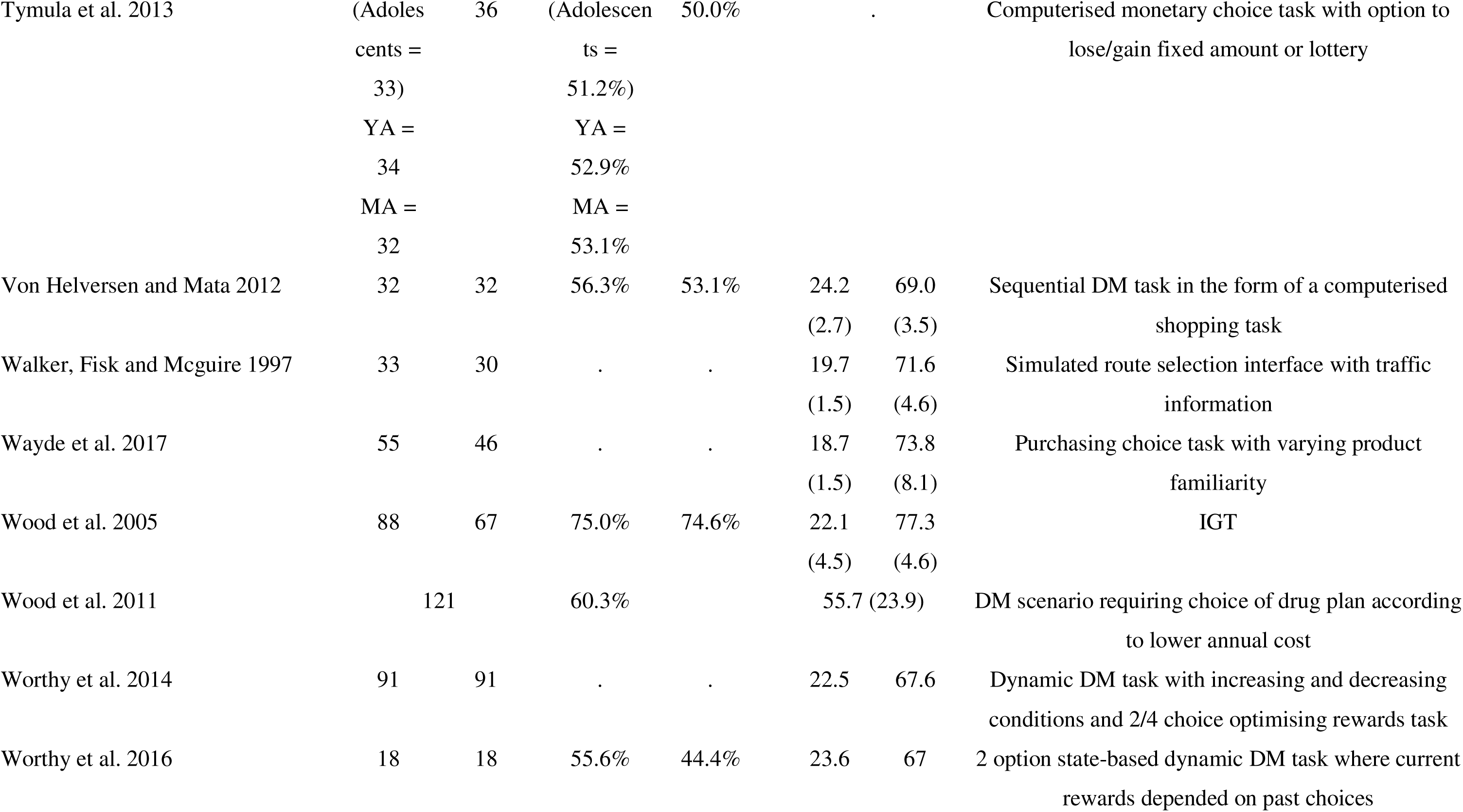
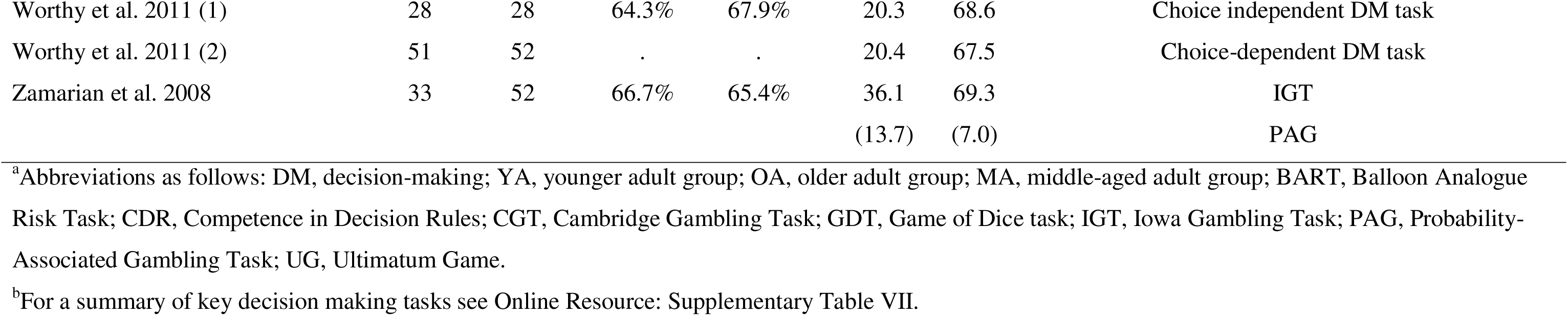
Summary Characteristics of Included Studies.

Most studies employed tasks that involved financial decision-making (*k* = 36). There were limited numbers of studies with tasks representing social (*k* = 14), health (*k* = 5), and safety (k = 4) decision-making processes. Twenty-five studies employed tasks that did not fall into financial, social, health or safety domains. Twenty-eight studies involved tasks covering two or more decision-making domains (e.g., financial and social).

Table II shows the results and analyses in detail. Overall, the majority of the 87 studies found that older age was associated with poorer decision-making on one or more outcome measure. Twenty-two studies reported no significant associations between age and decision-making, and 16 reported better decision-making amongst older as compared to younger adults on one or more measures. Three studies provided narrative summaries of overall age differences without numerical results, preventing their inclusion into meta-analysis (Cavanagh et al. 2012; Hess et al. 2012; Ma and Chen 2015). The relationship between age and decision-making was primarily investigated using a between-subjects design, with only two studies reporting on intra-individual age-related changes in decision-making (Bauer et al. 2013; Boyle, Yu, Buchman, et al. 2012; Li et al. 2013; Li et al. 2015). A total of 57 studies from 53 publications were compatible for meta-analyses and the data were pooled to allow for estimates of age differences on decision-making tasks.

**Table II.**
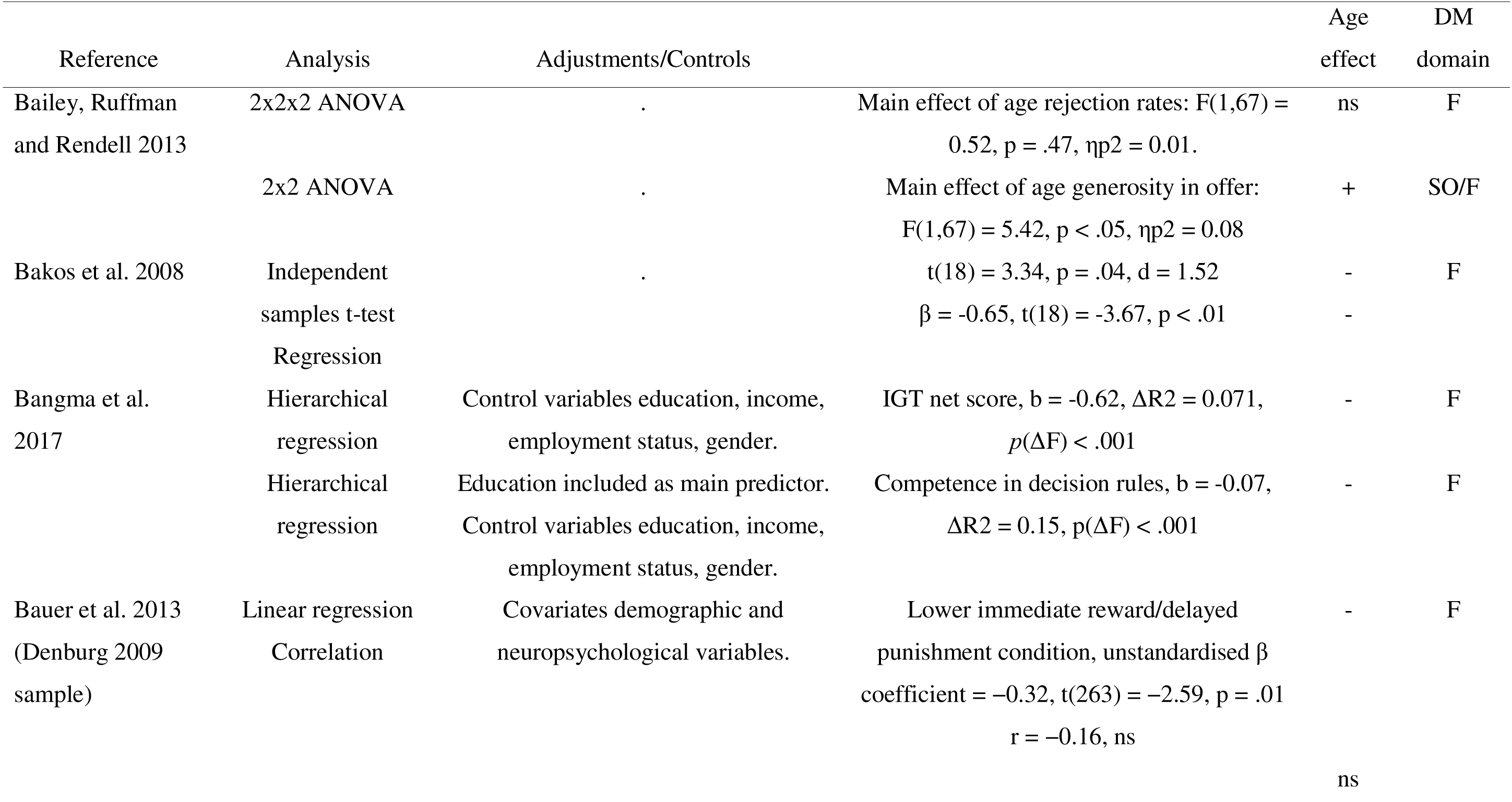
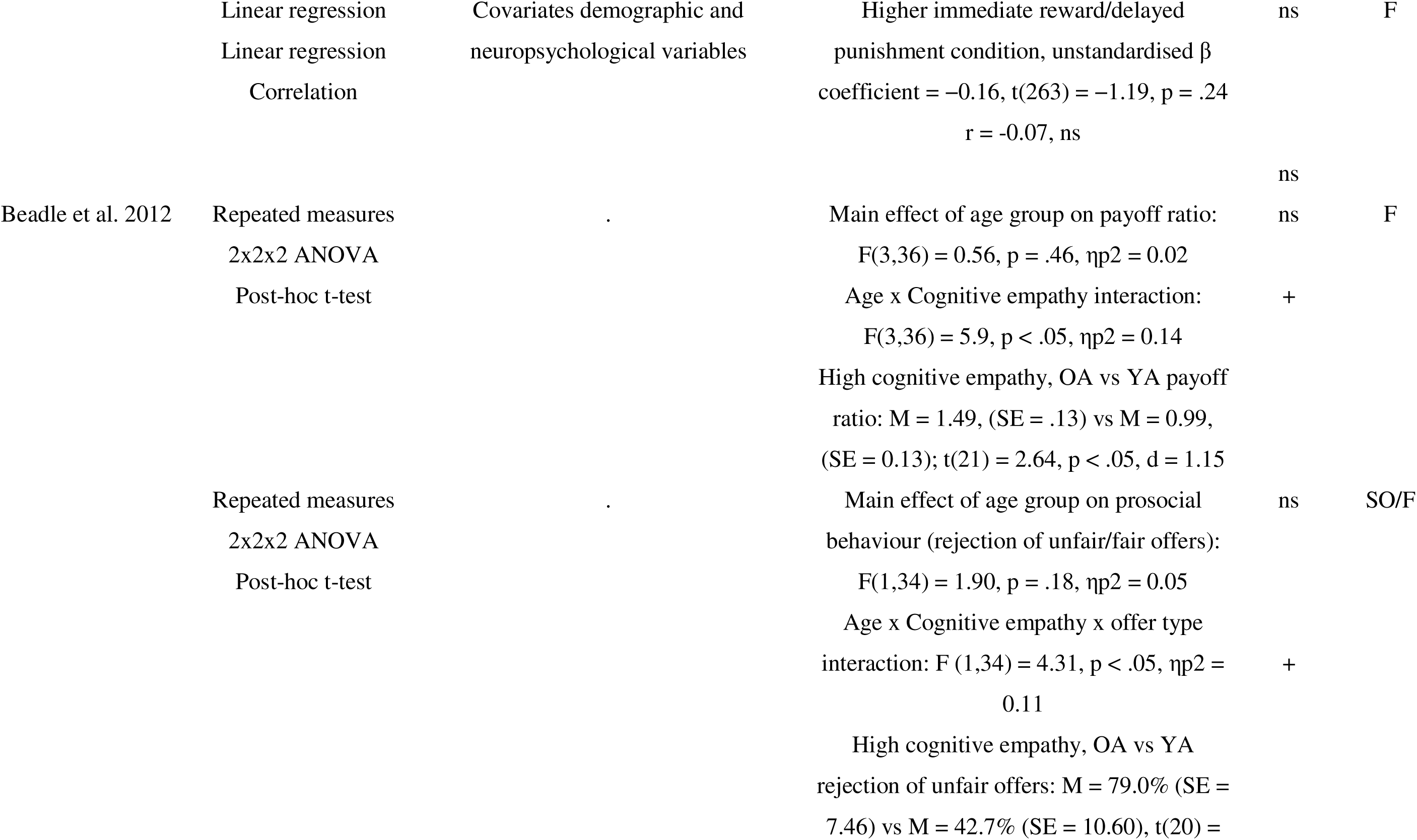
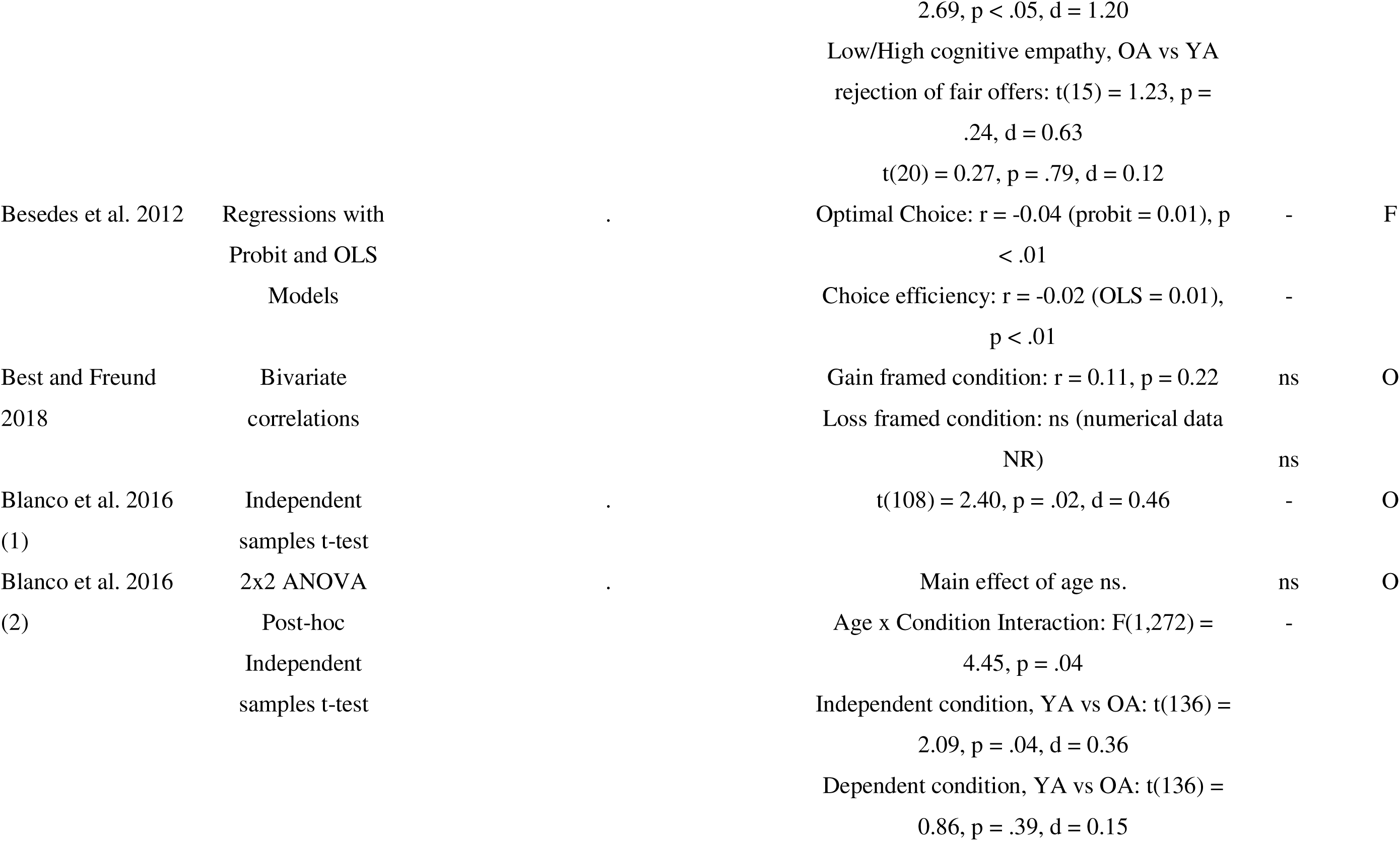
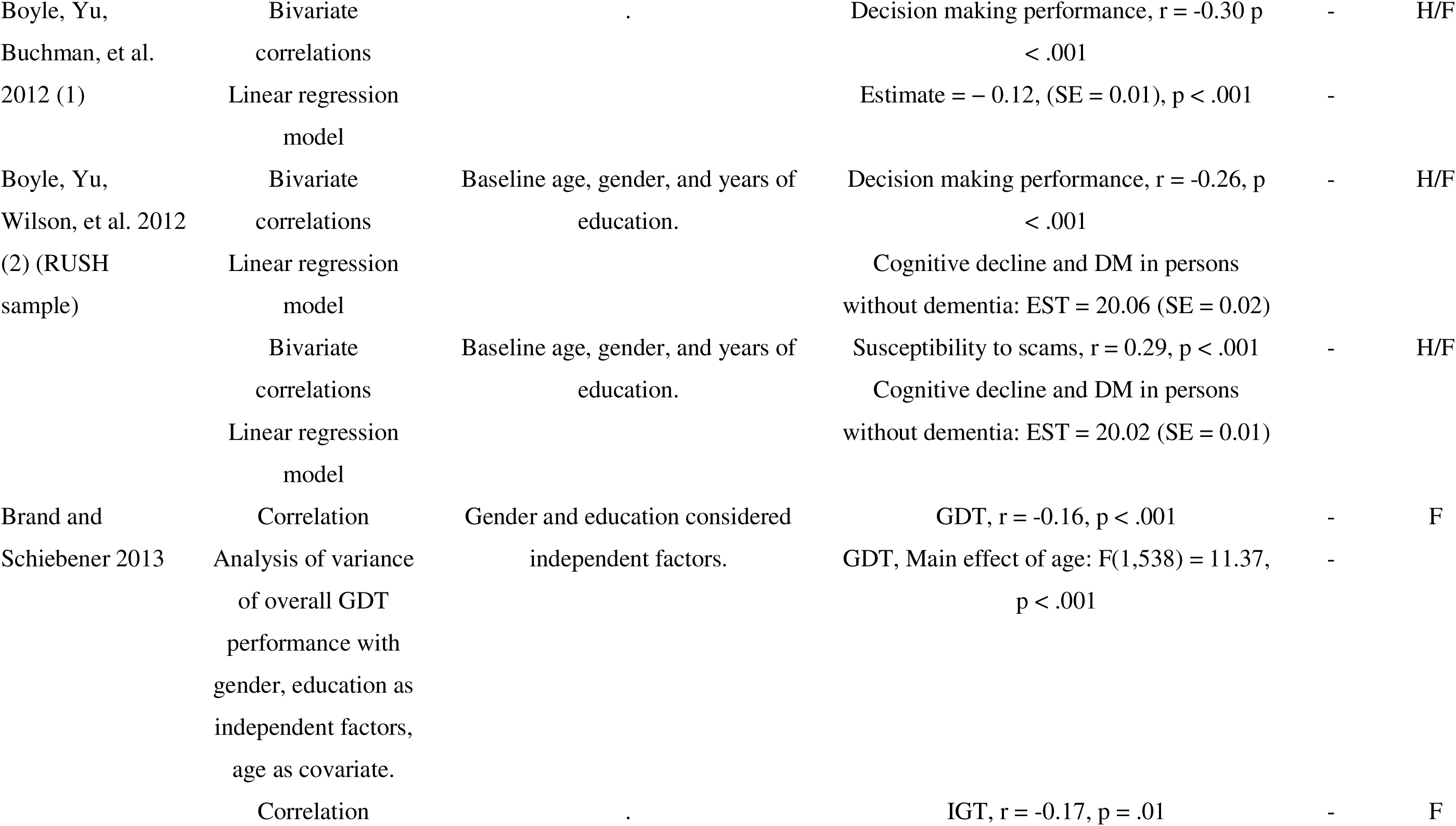
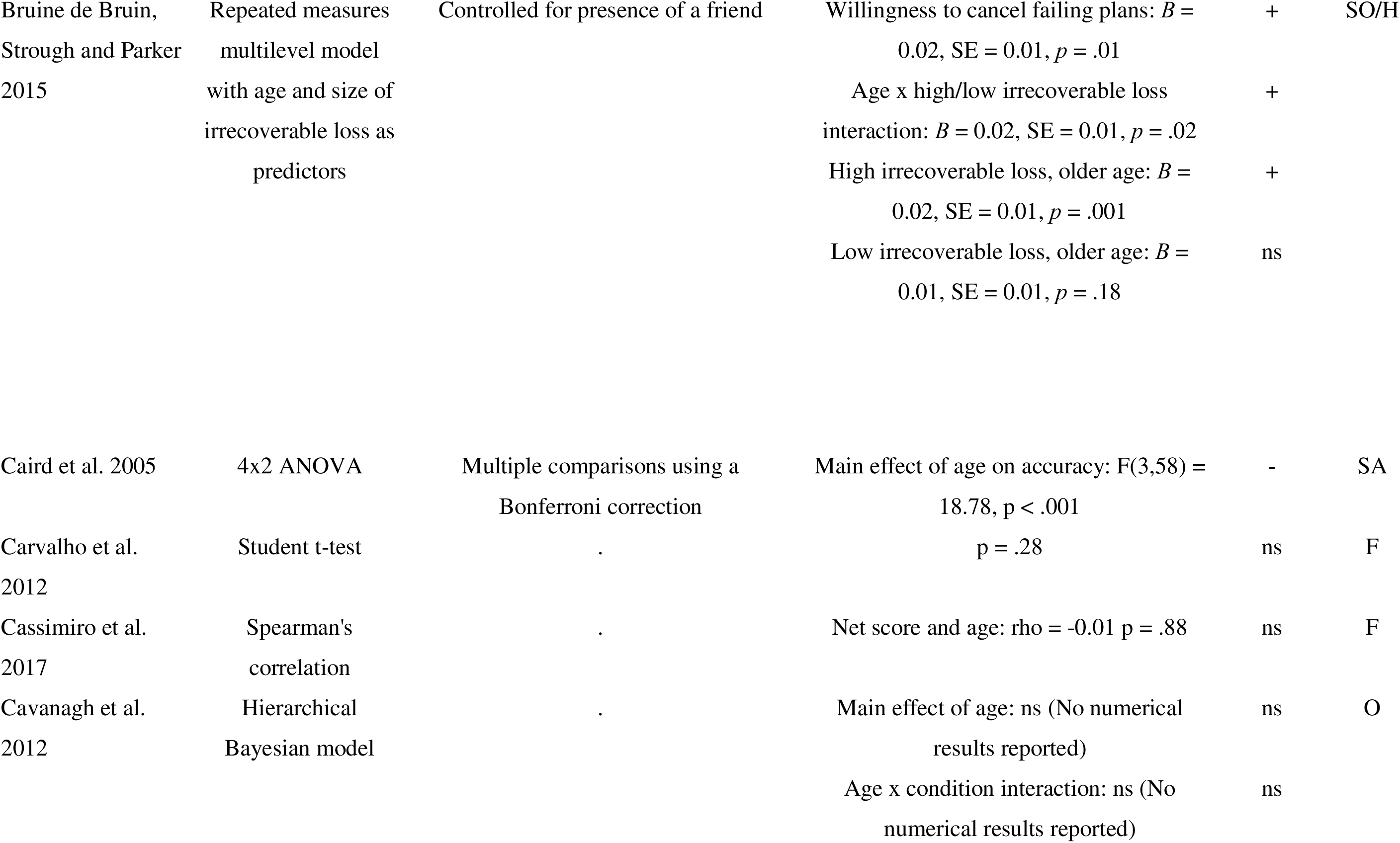
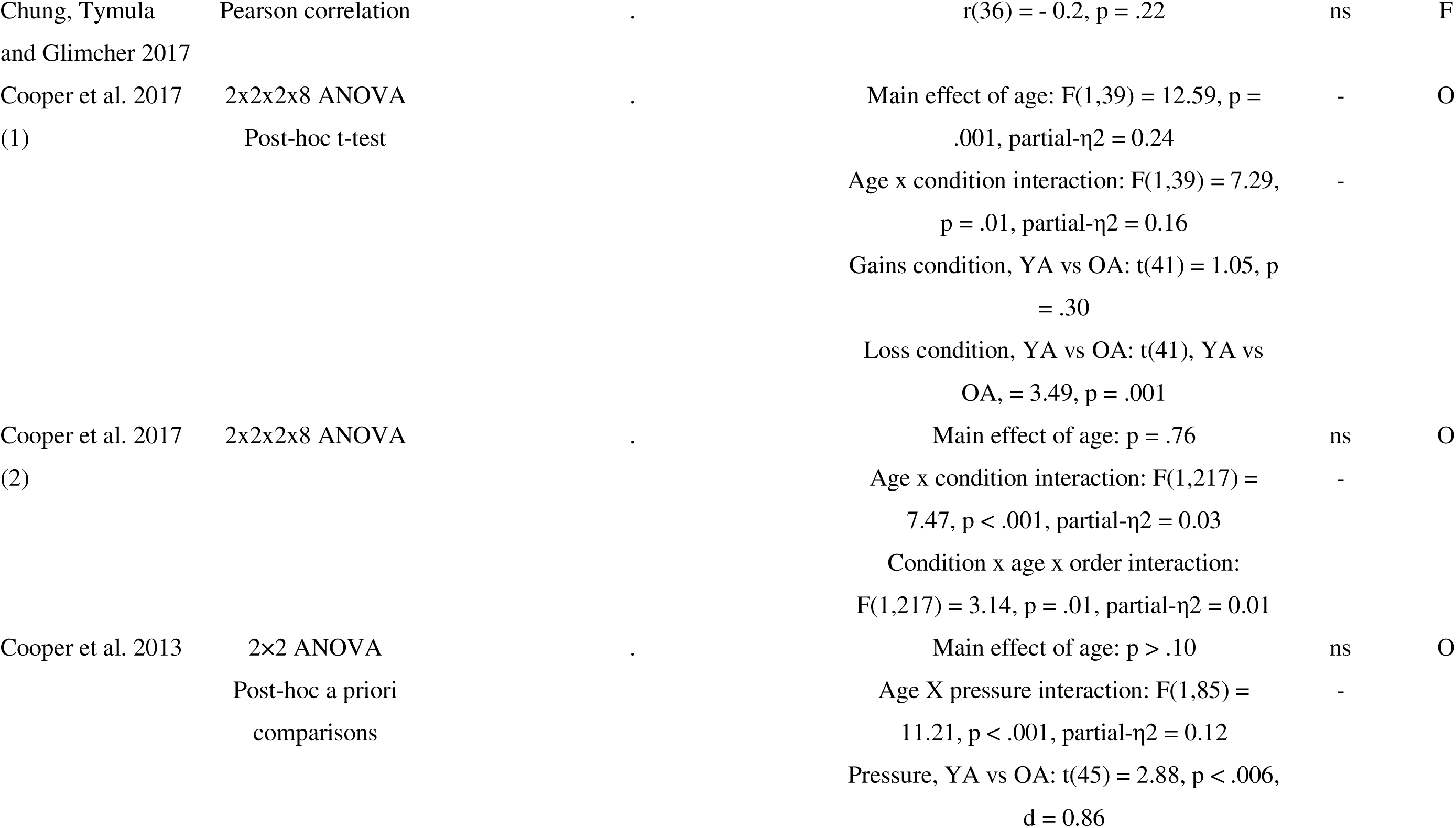
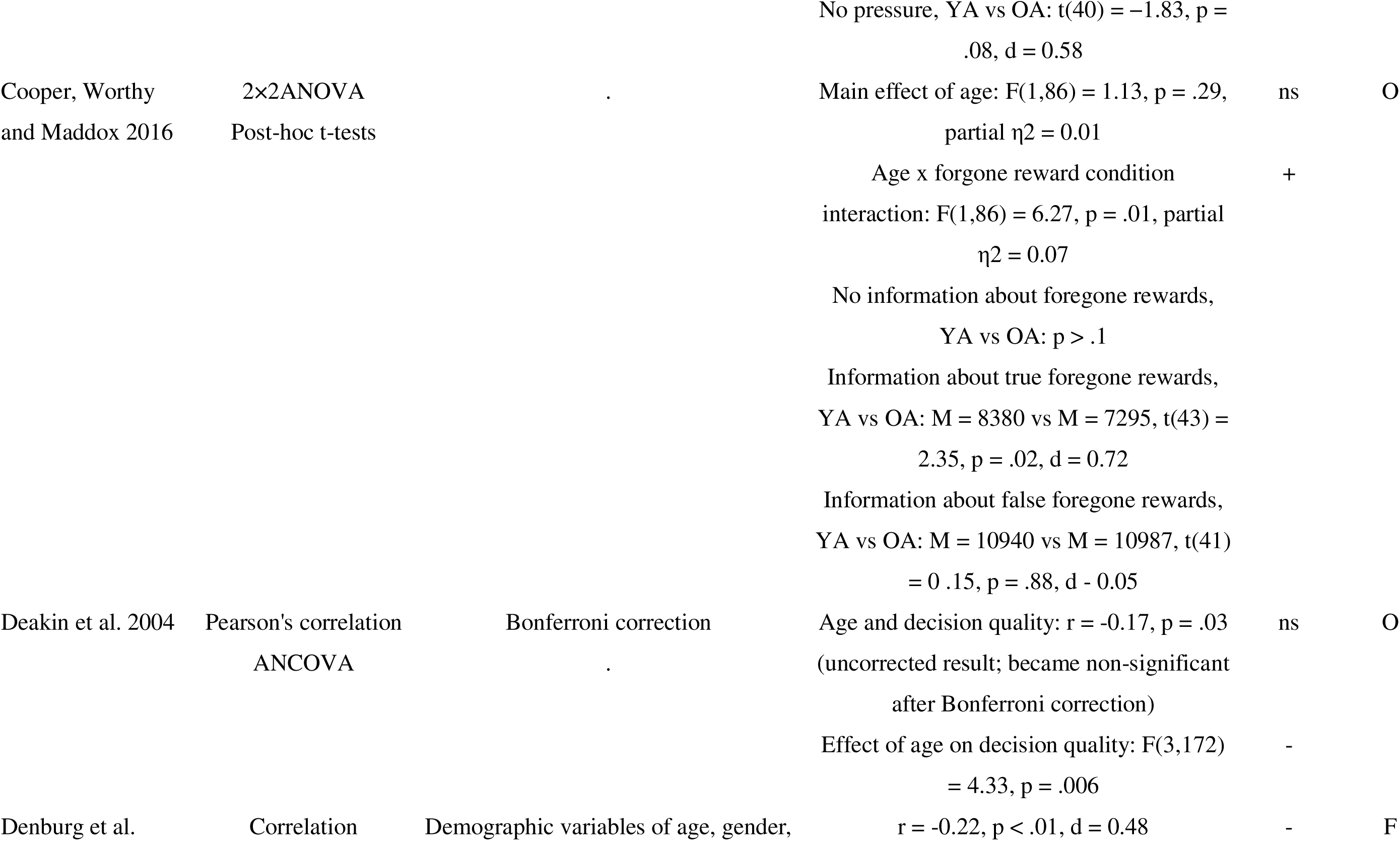
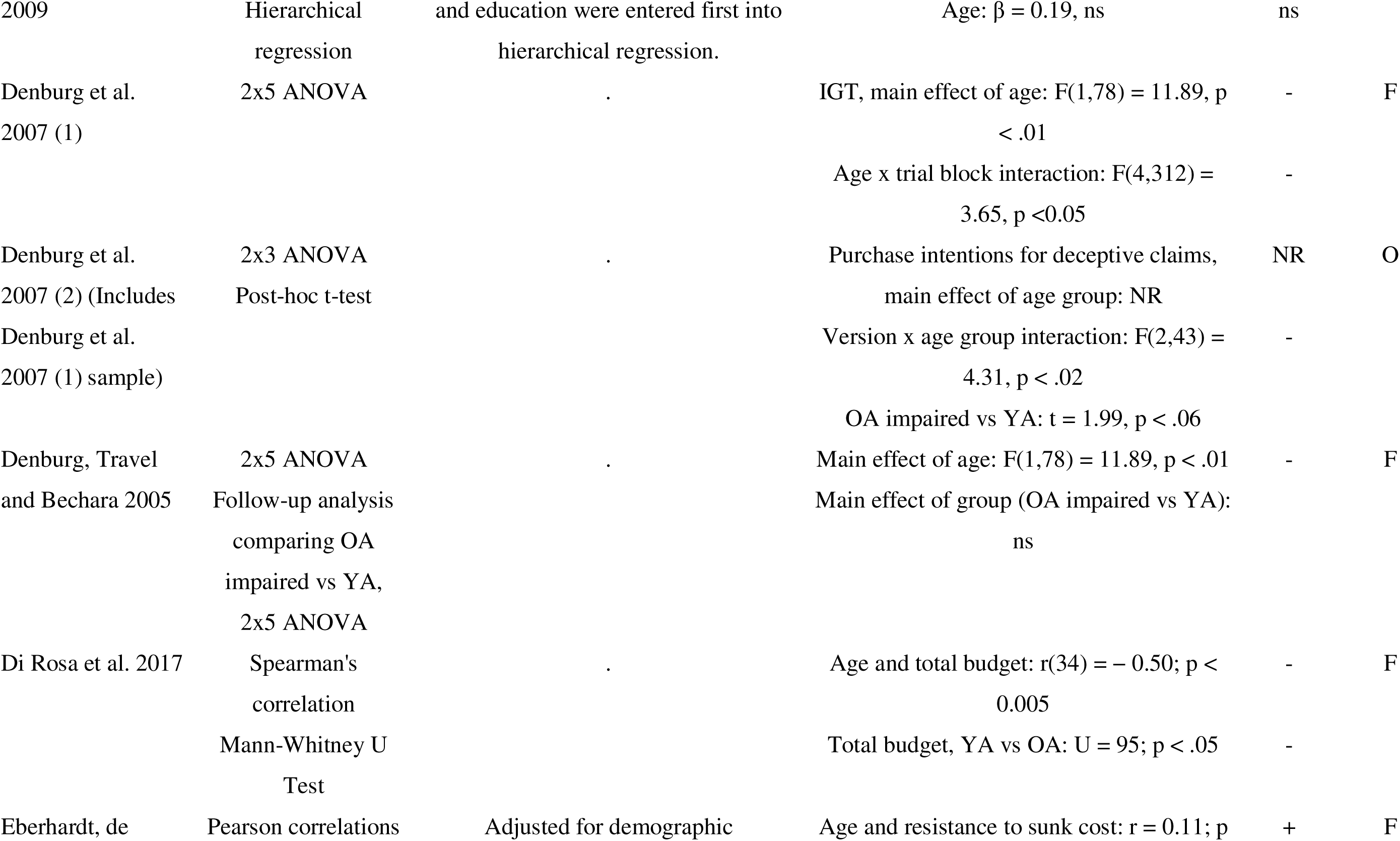
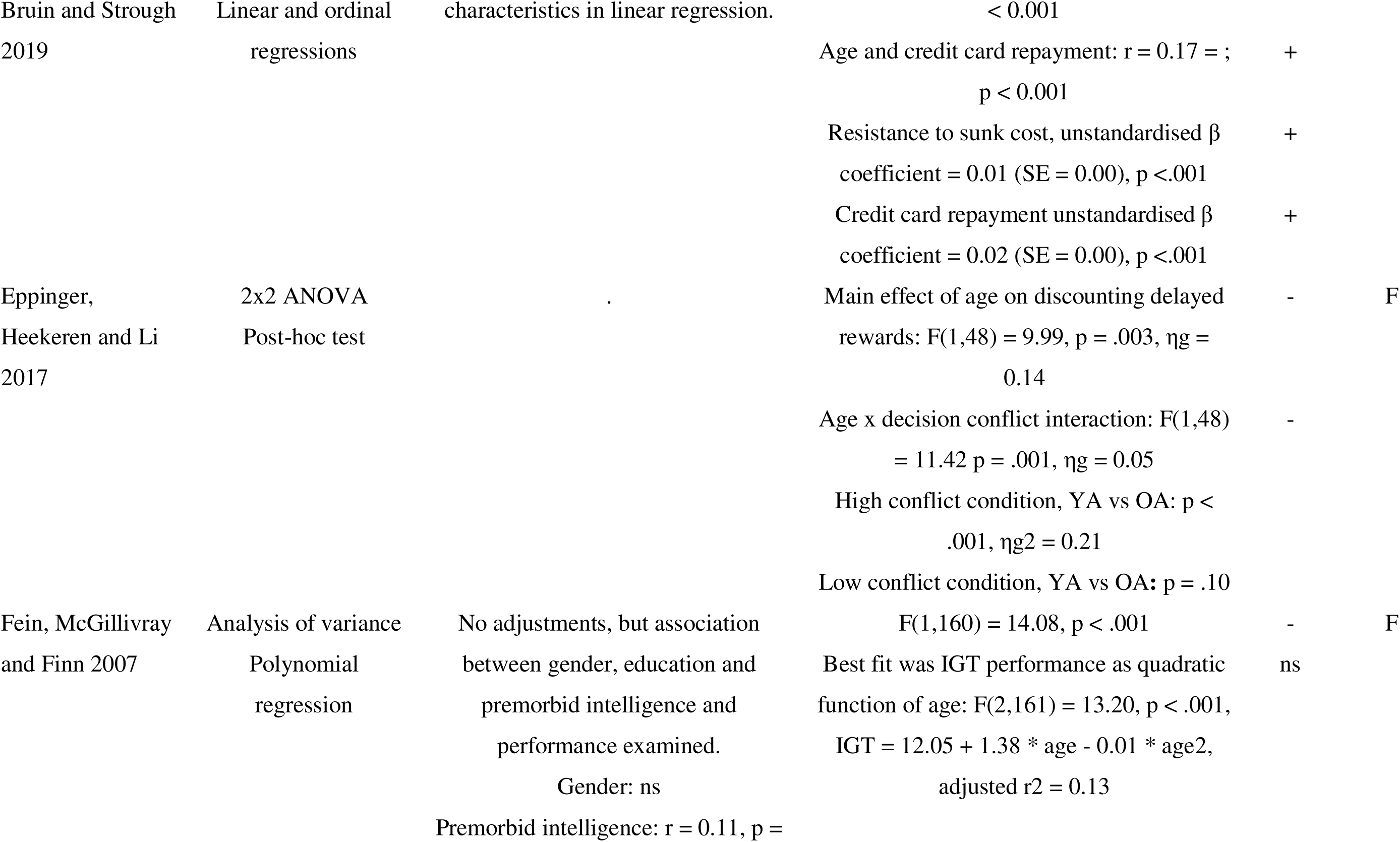
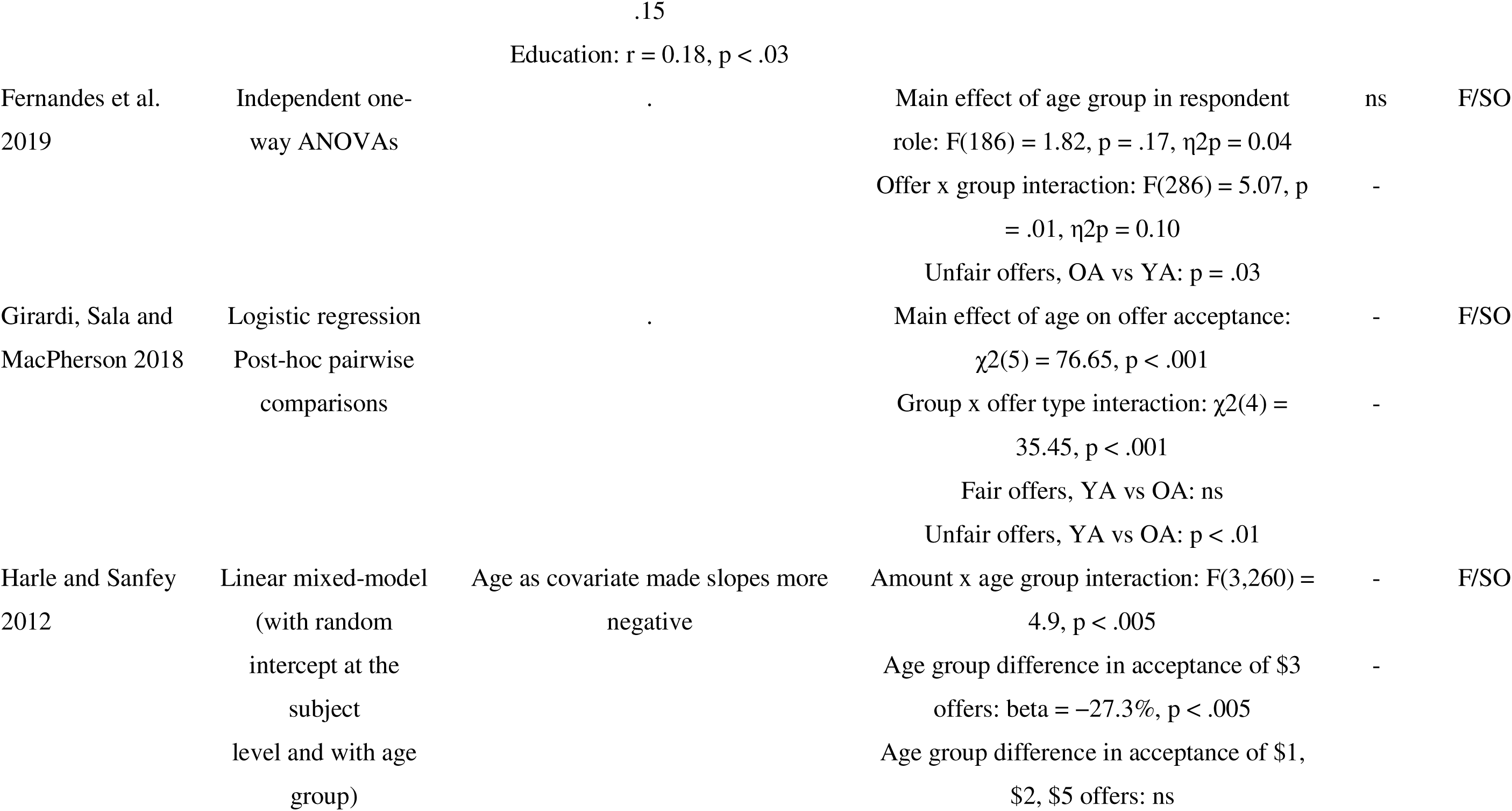
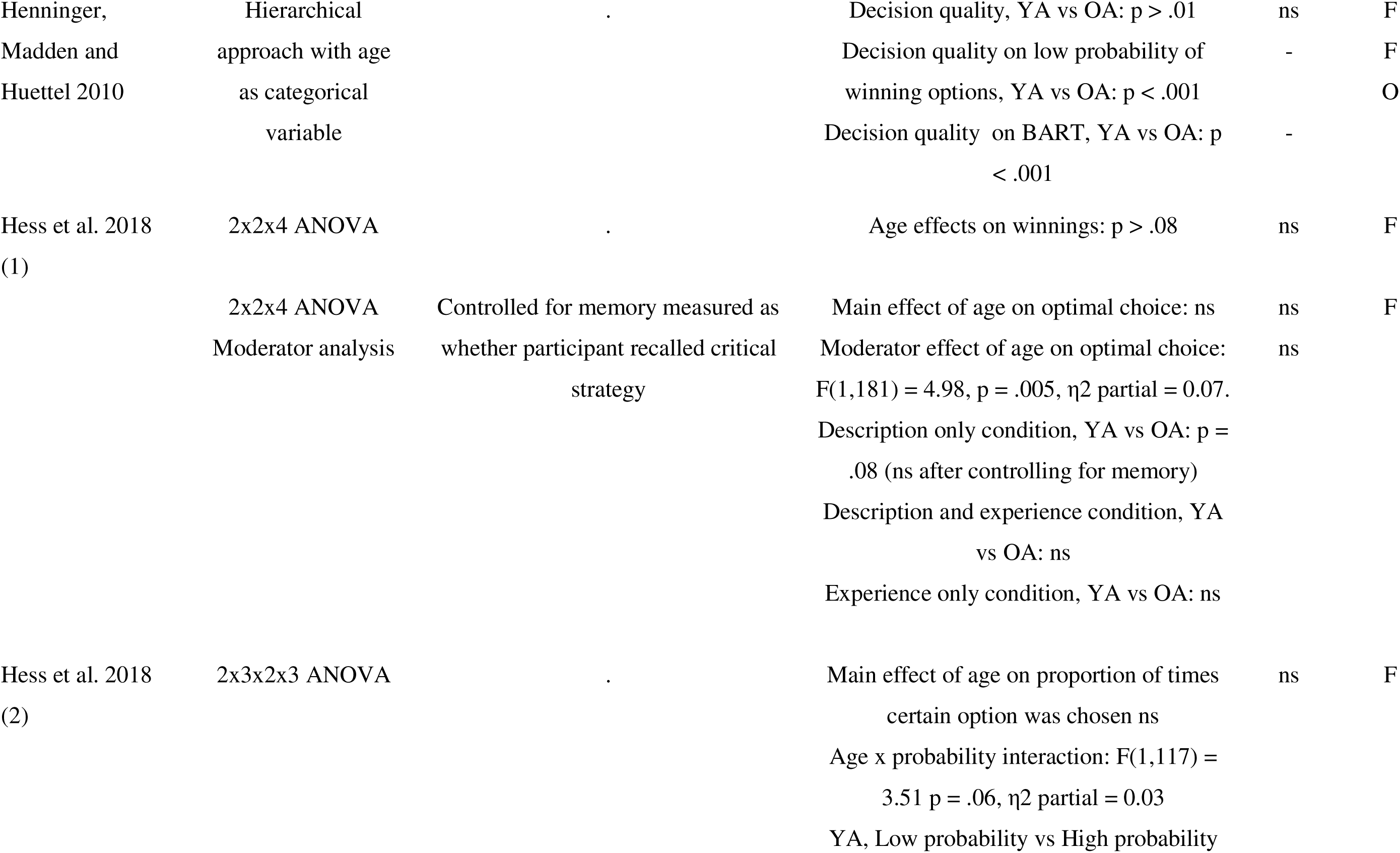
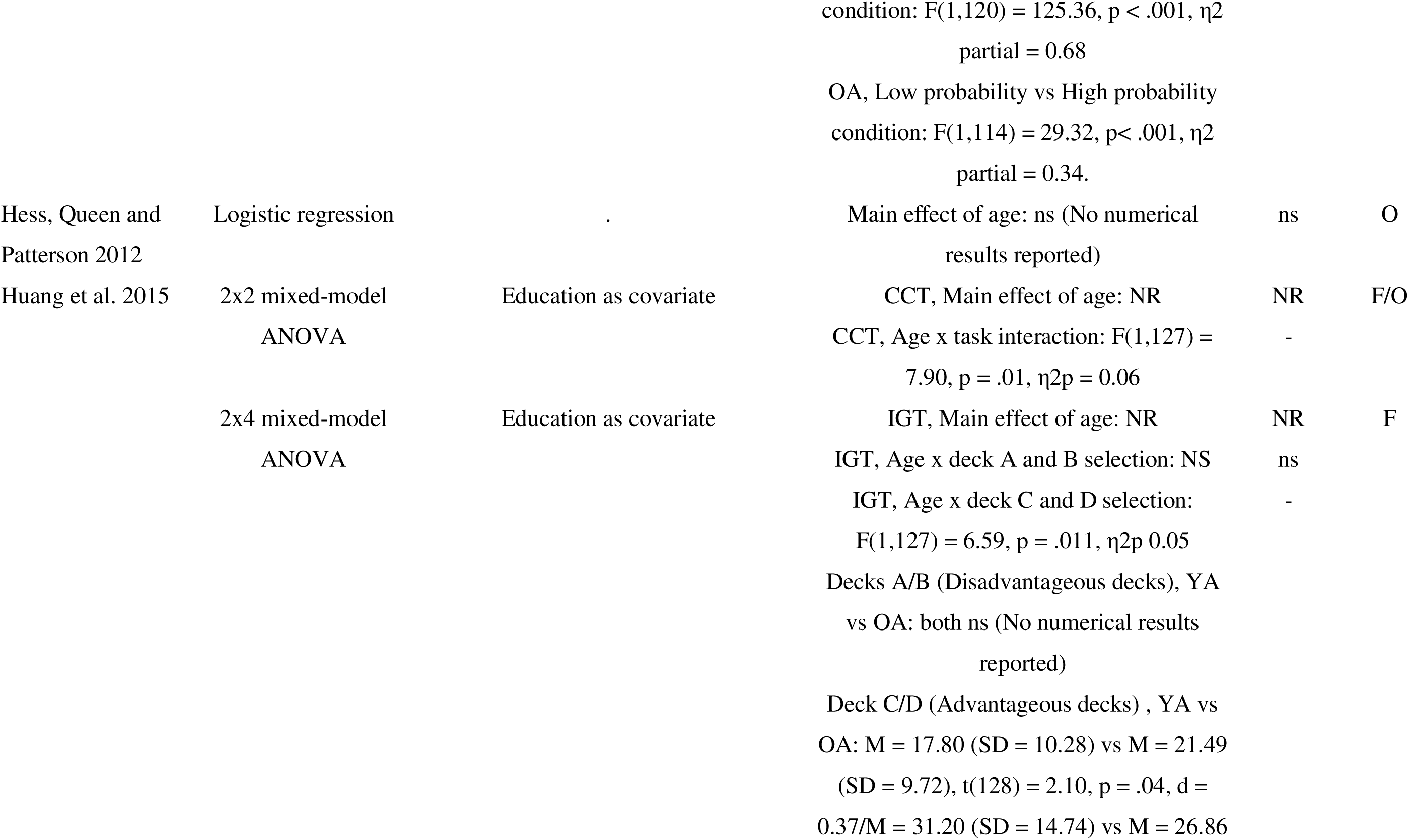
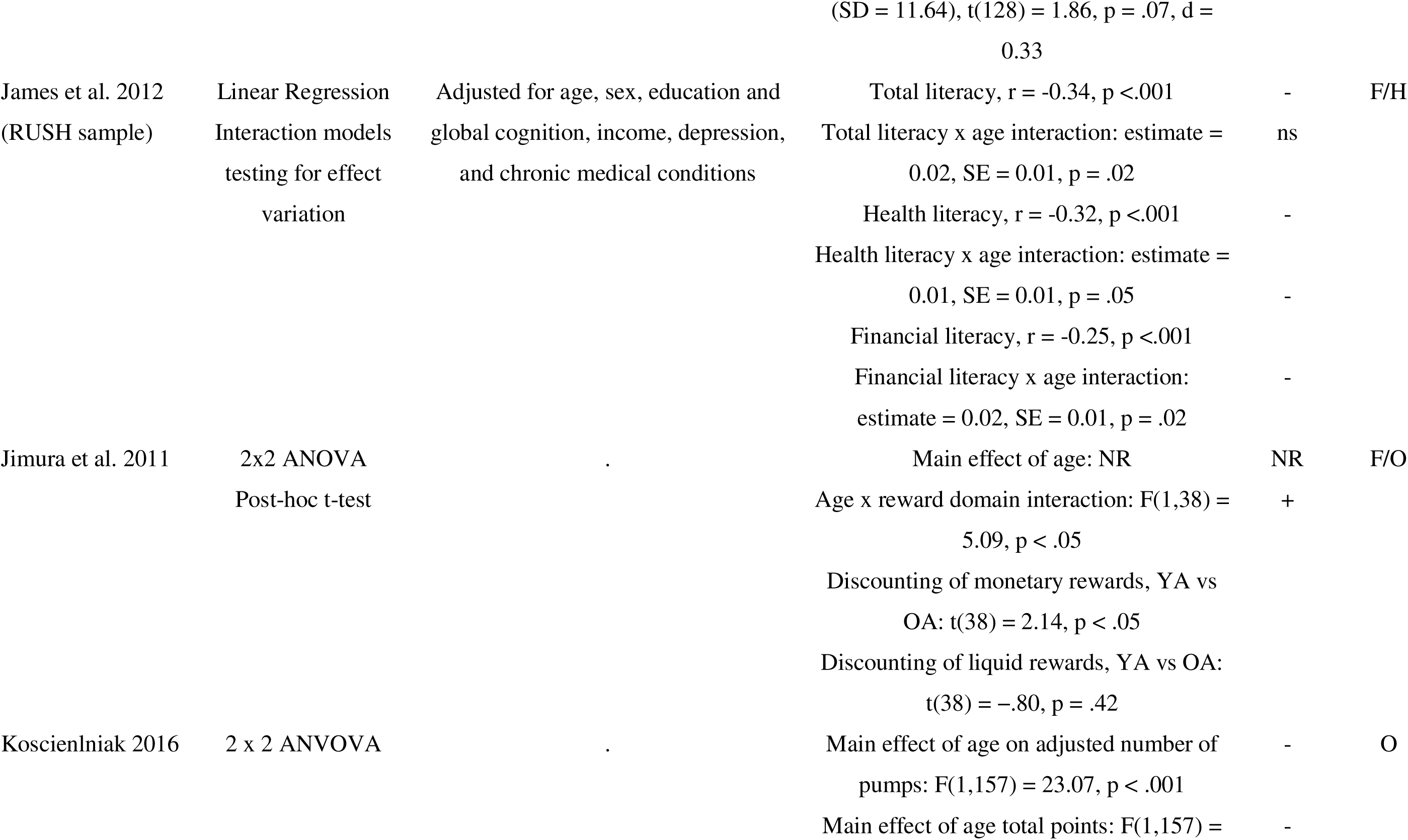
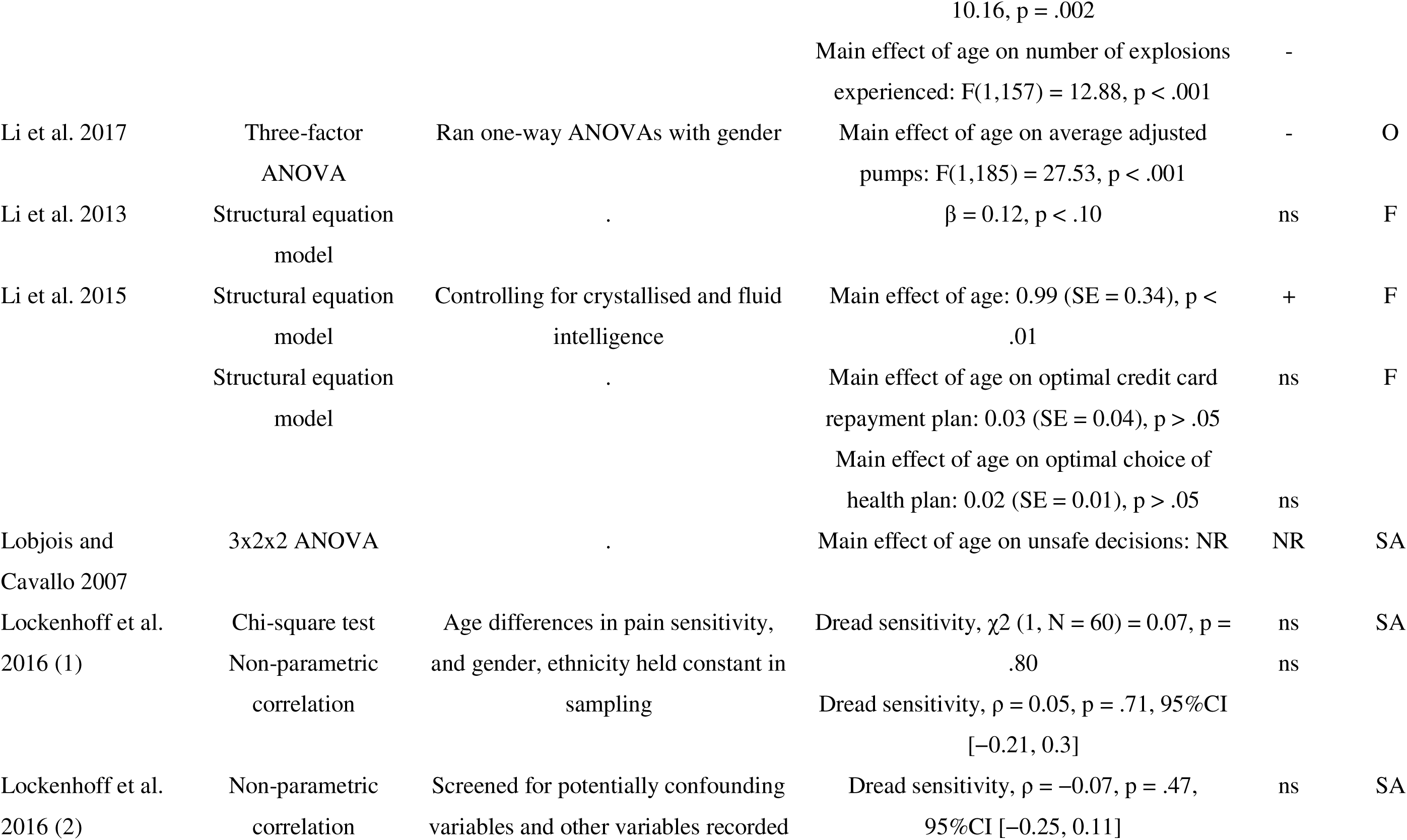
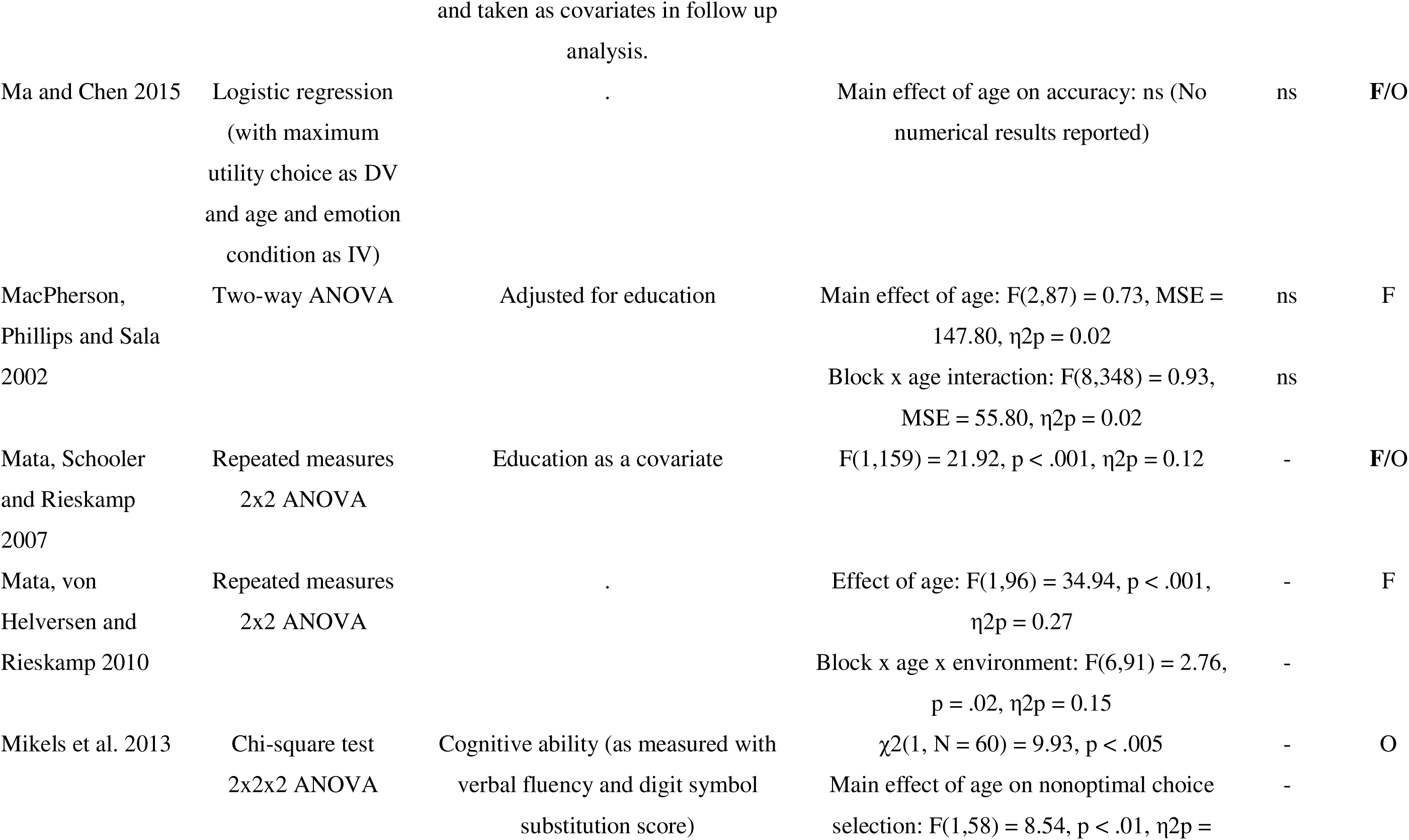
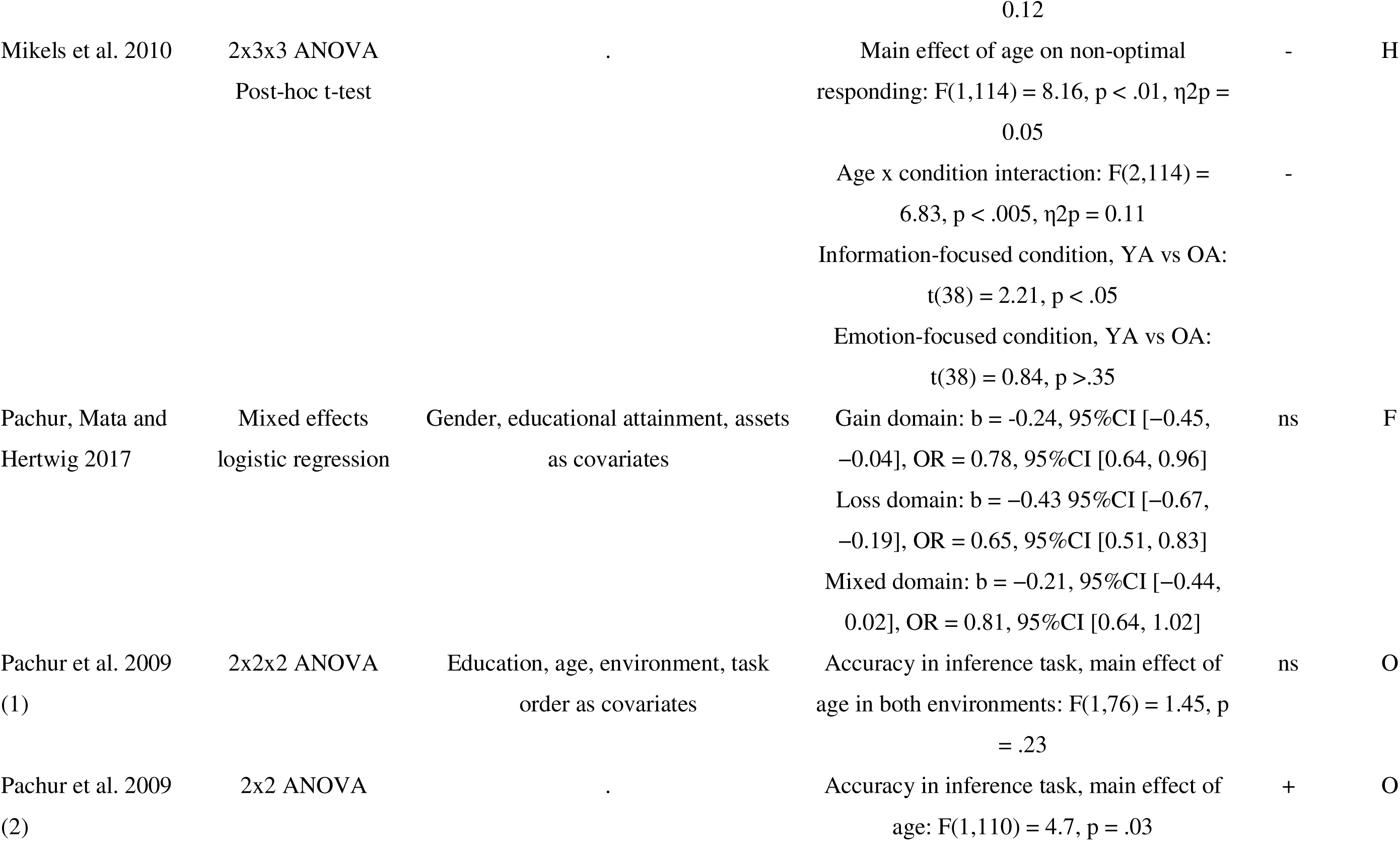
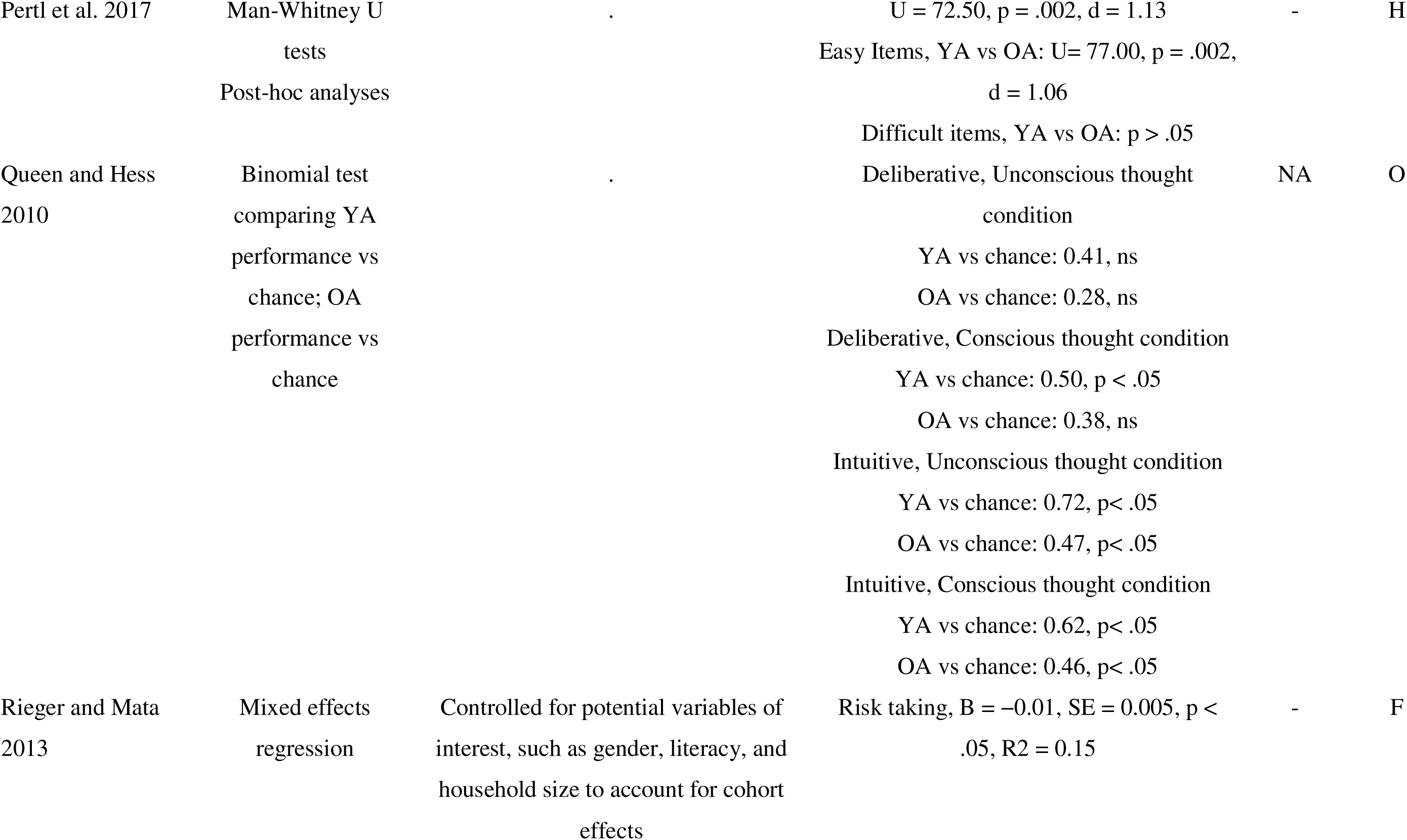
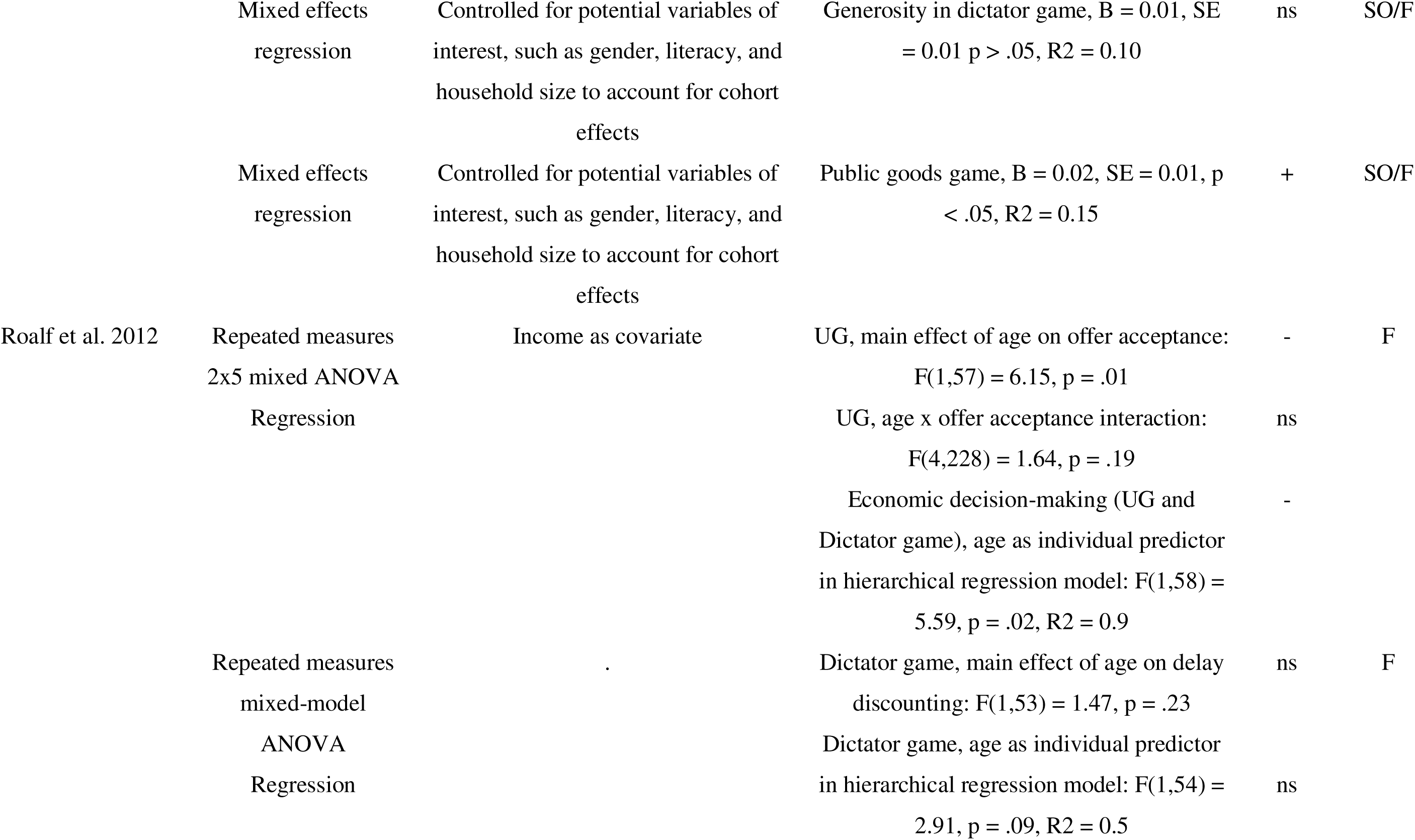
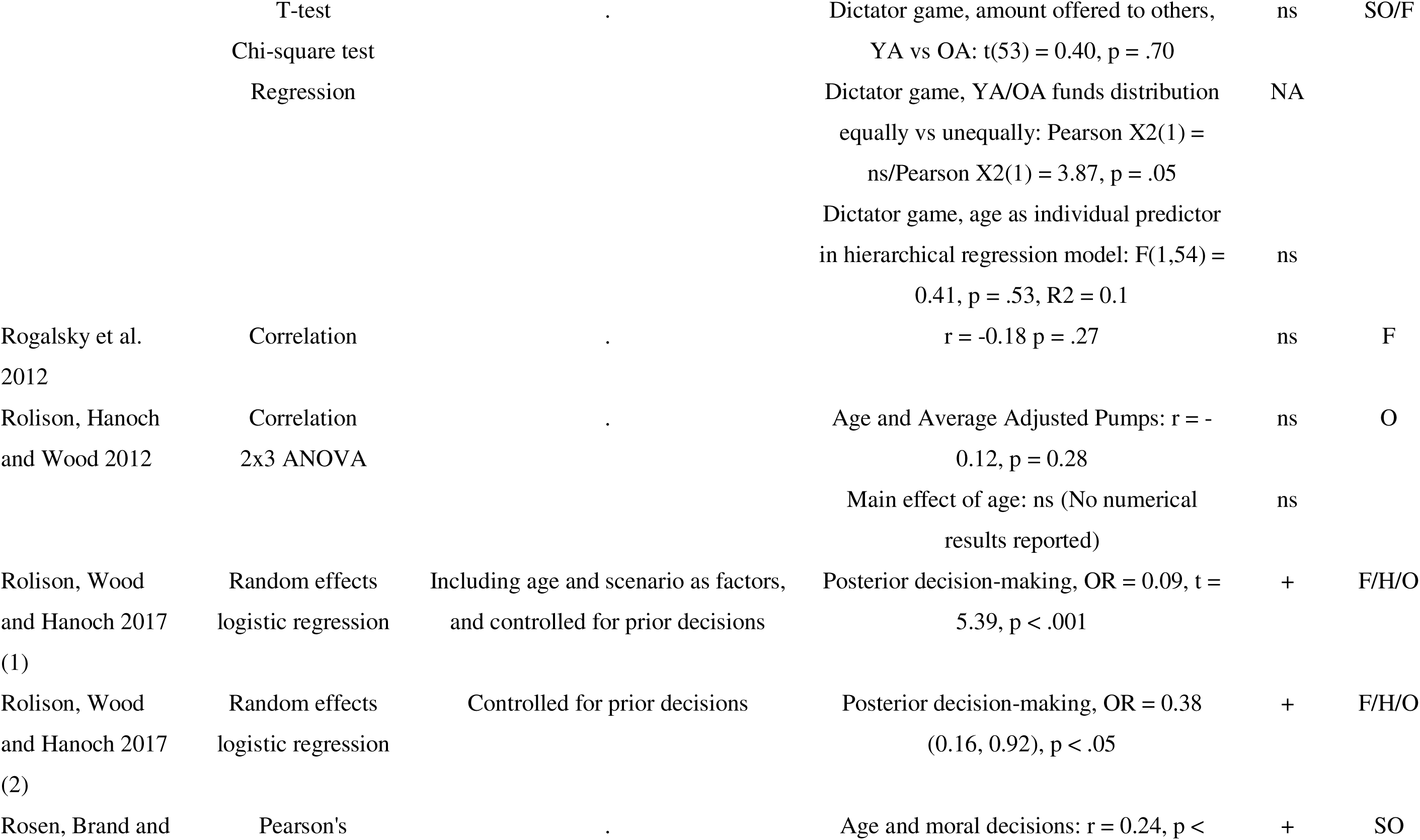
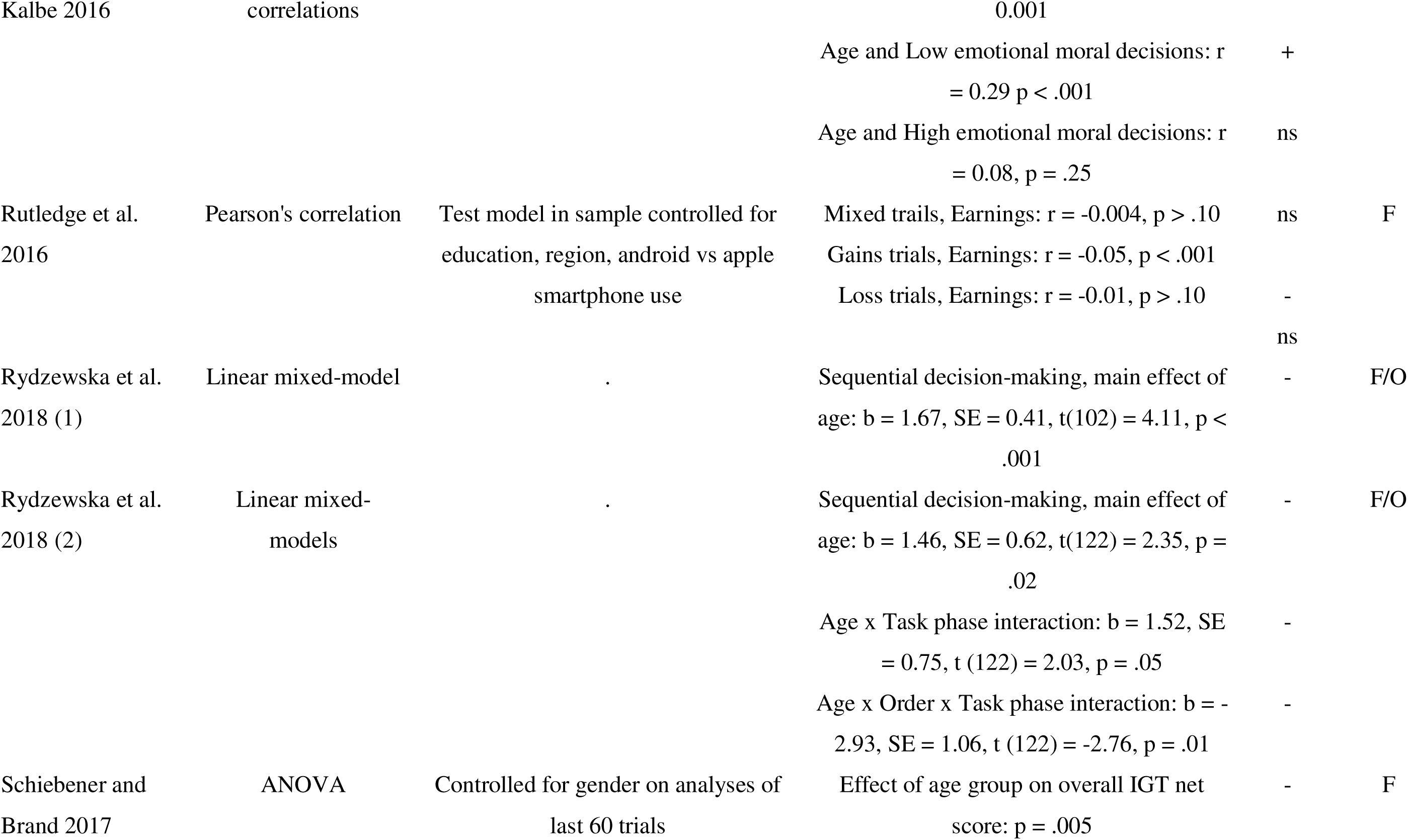
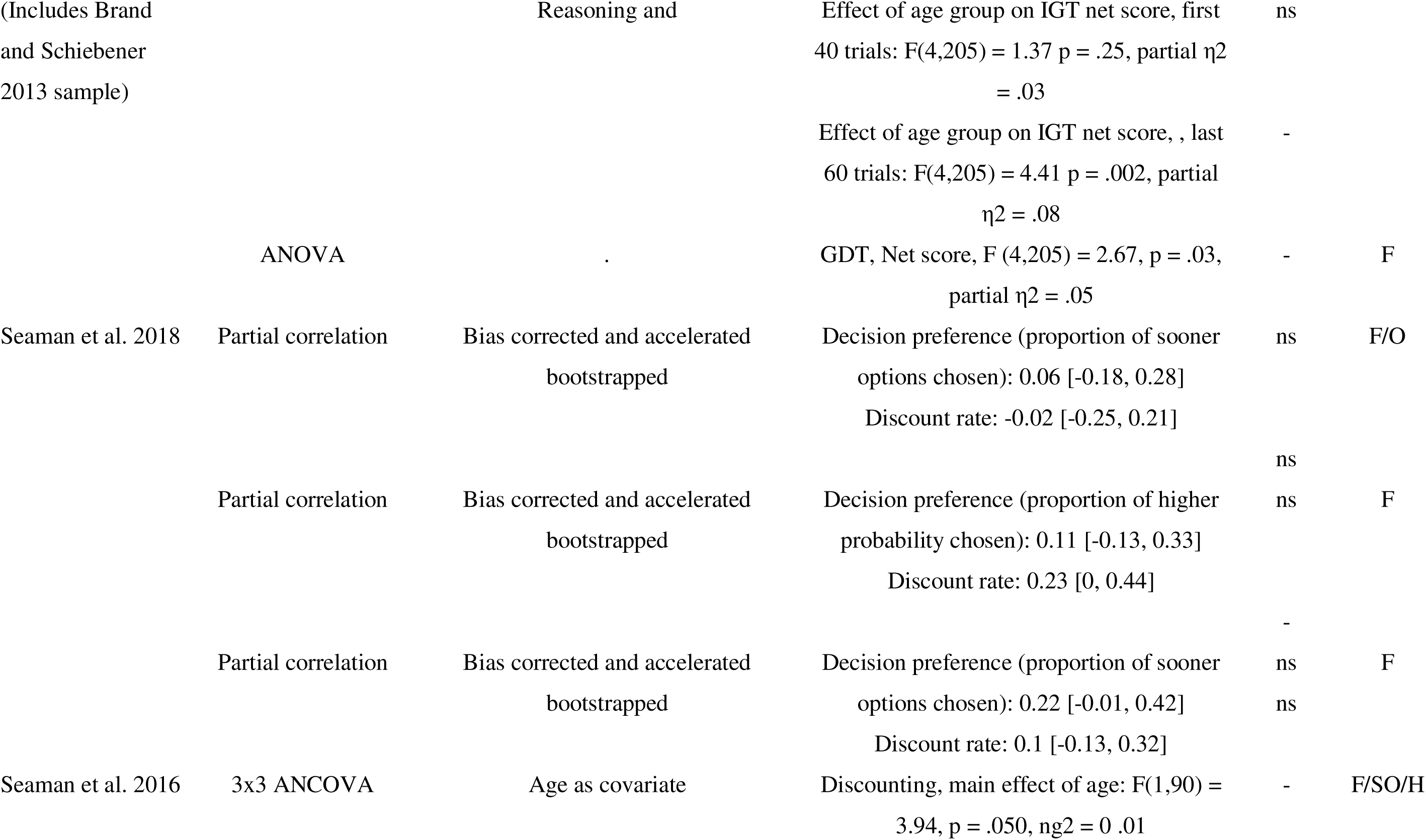
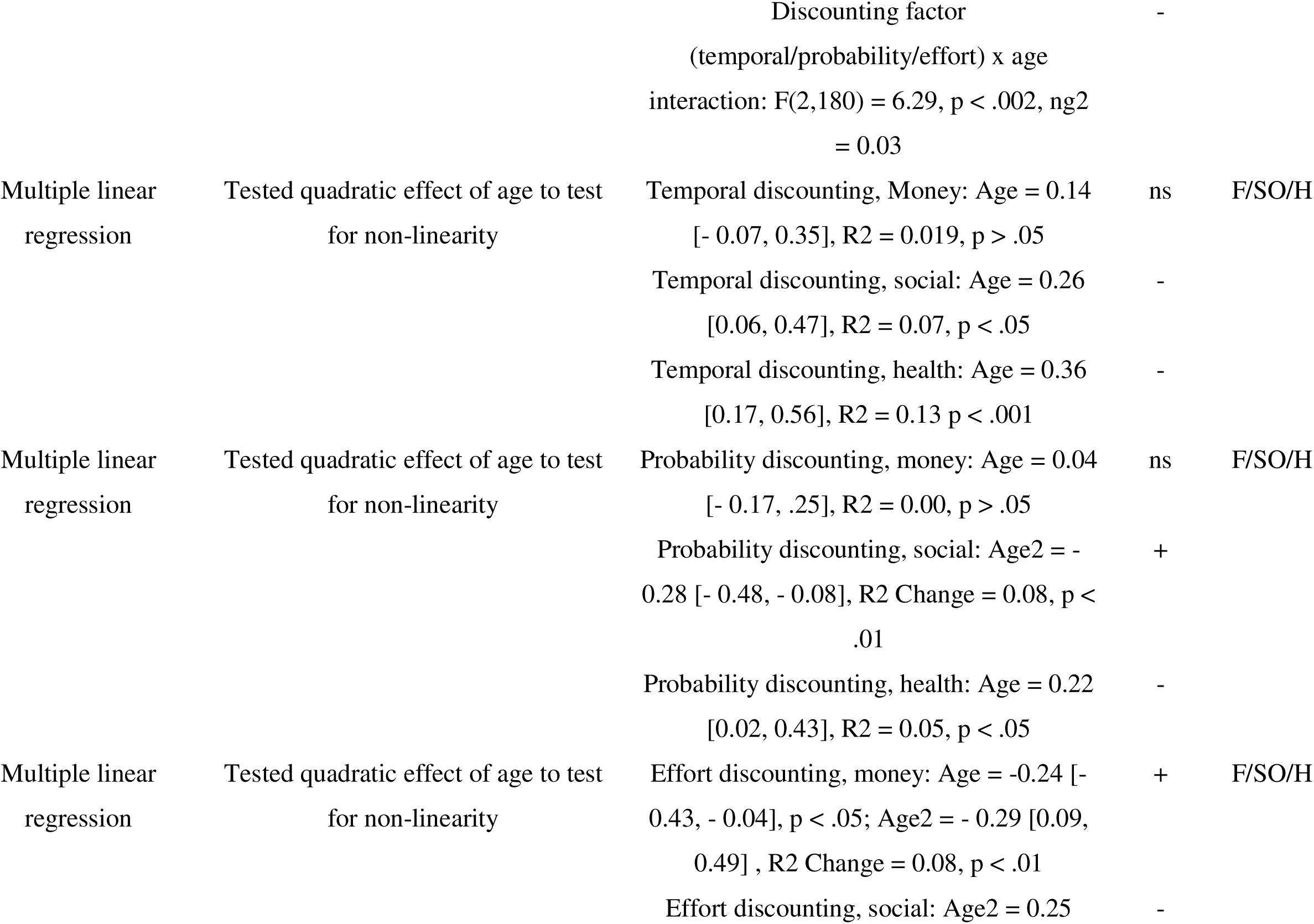
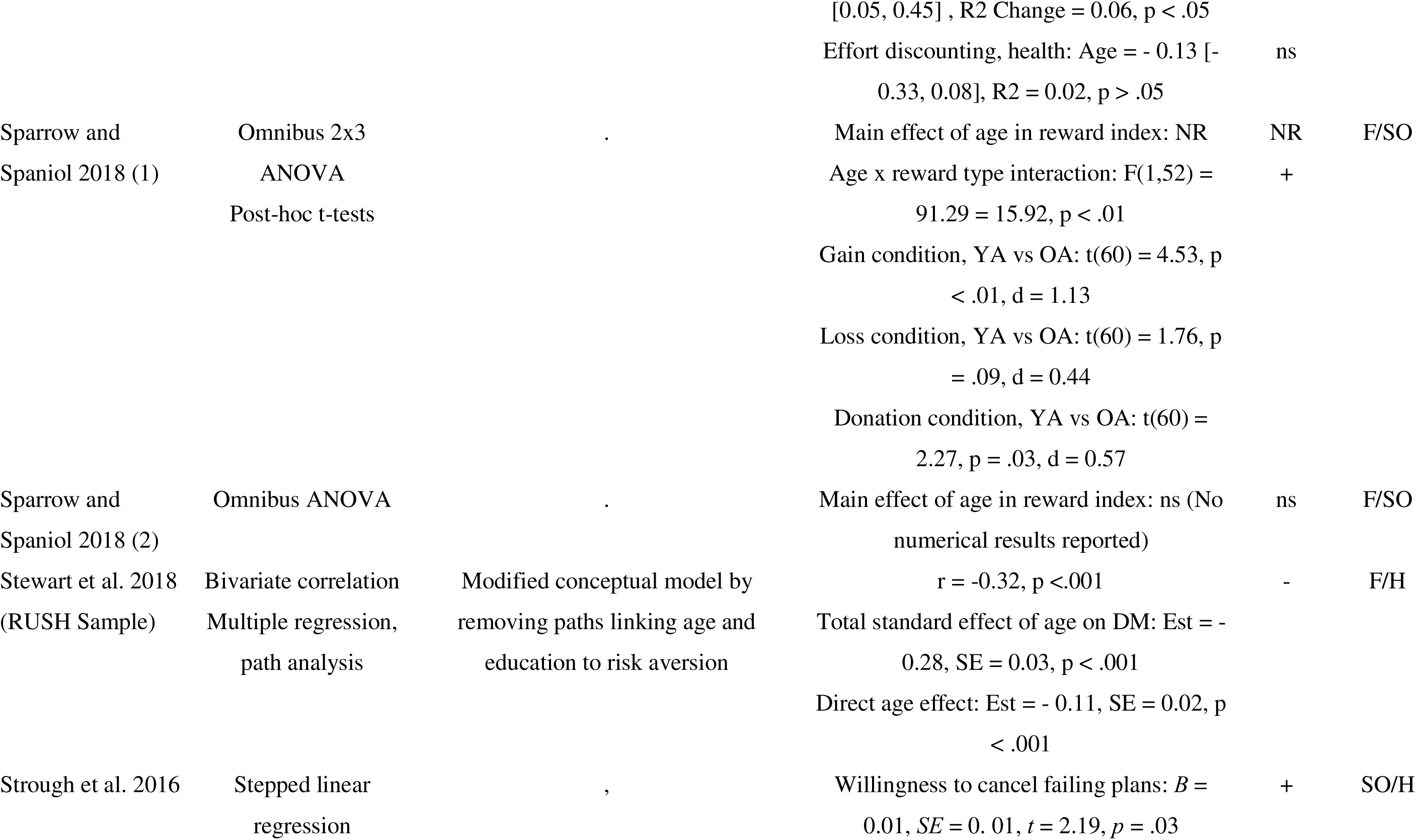
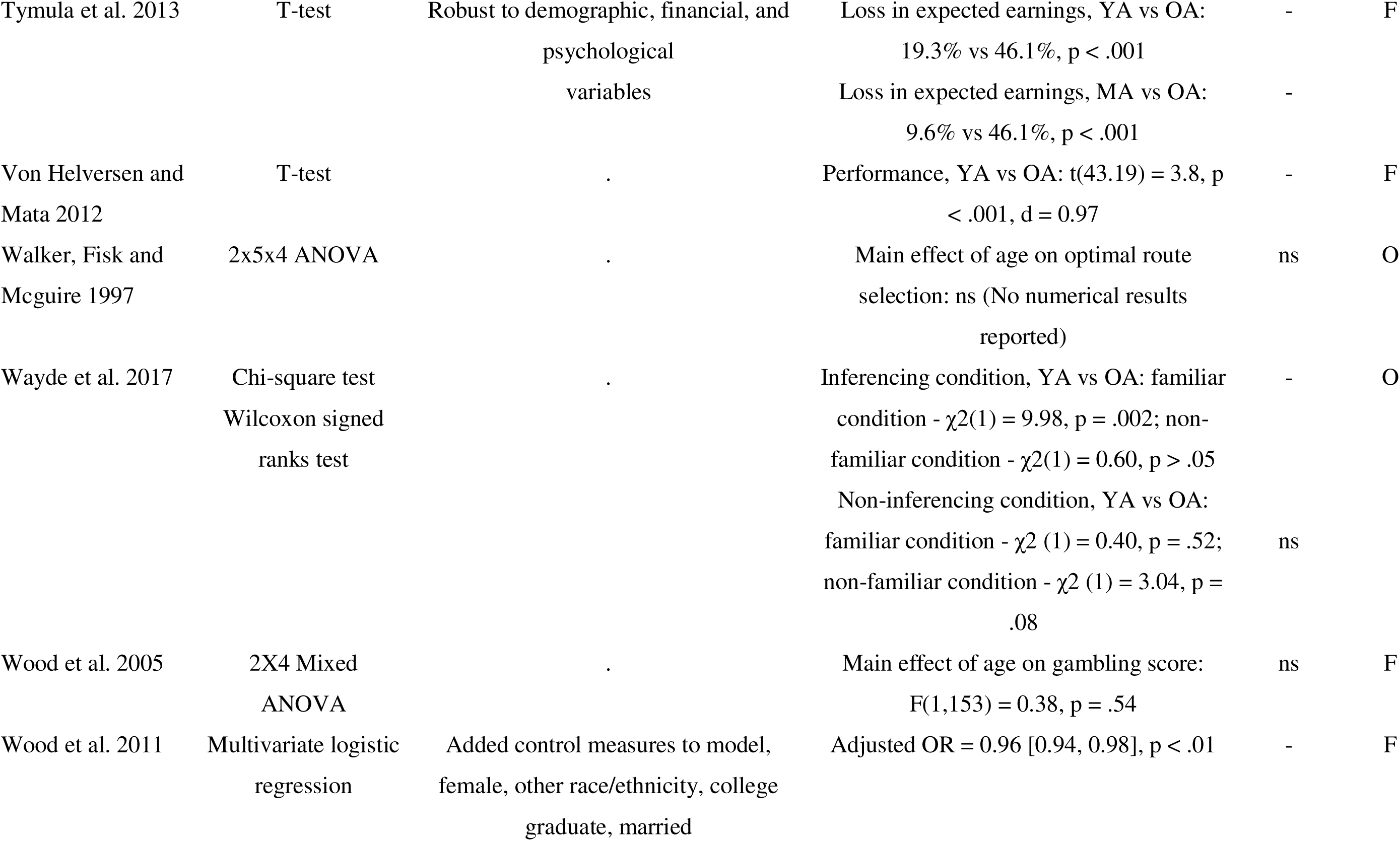
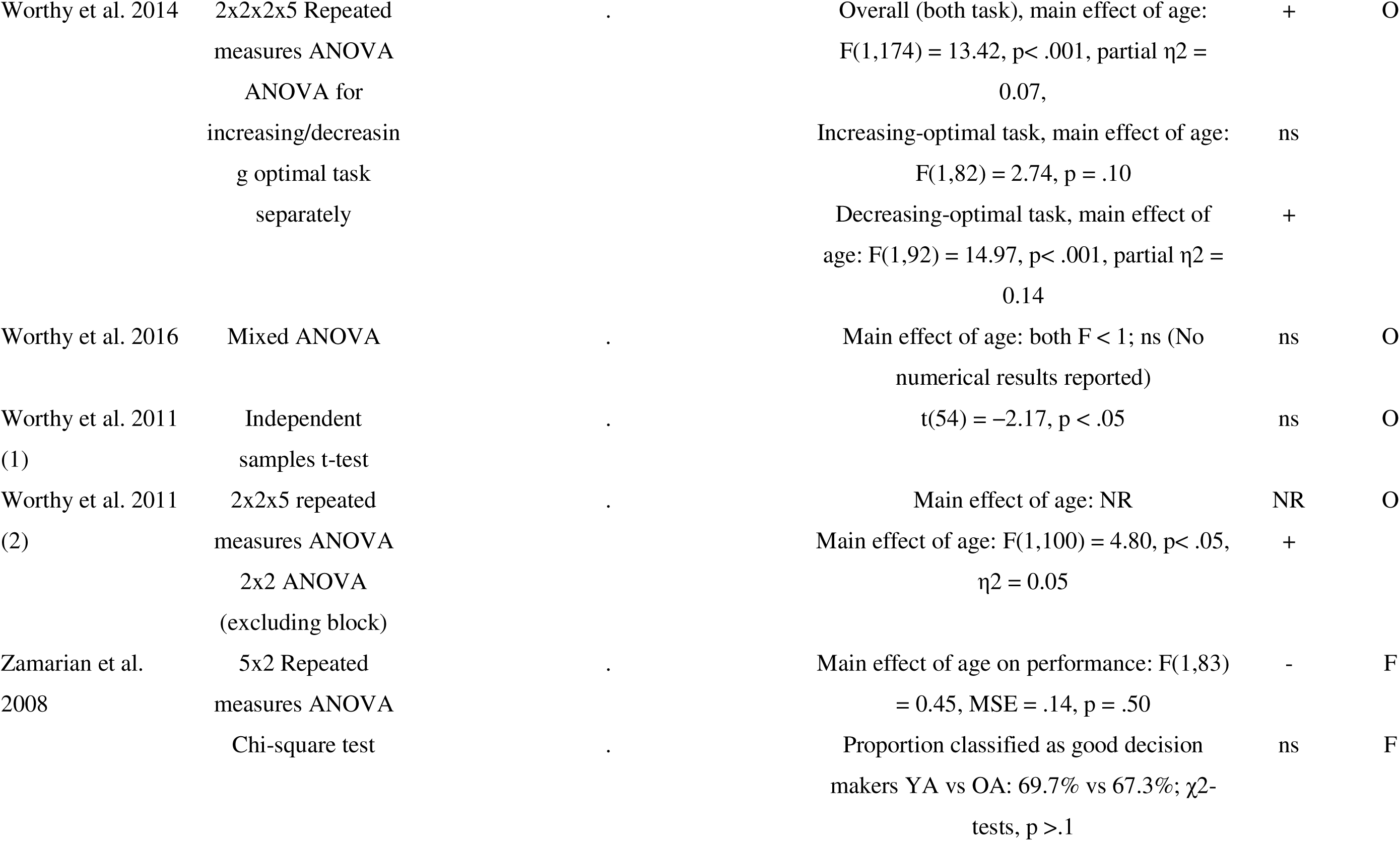
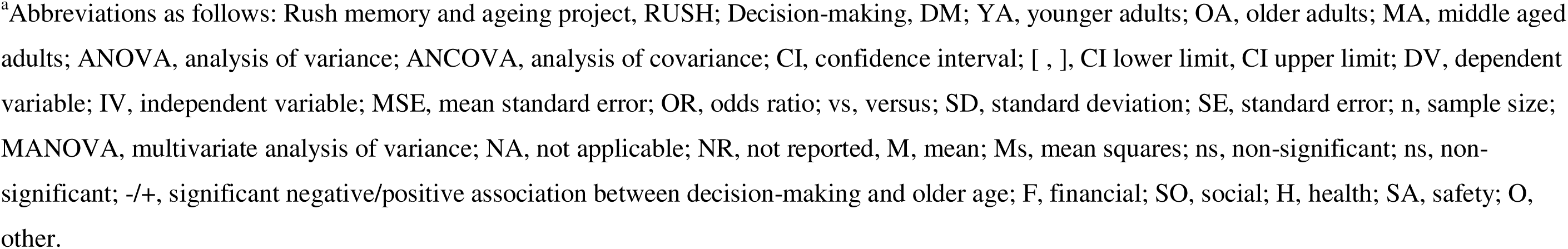
Summary of results and analyses of included studies.

### Meta-analysis of studies comparing decision-making in older and younger adult

When meta-analysis was carried out the combined estimate was statistically significant in indicating poorer decision-making in older as compared to younger adults (k = 57, *d*_random_ = -0.17, 95% CI -0.29, -0.04, *I^2^* = 92.92%) (shown in Fig. I) however, heterogeneity was high and individual studies varied considerably (for funnel plots refer to Online Resource: Supplementary Fig. III). This finding was magnified when studies of intertemporal judgement tasks were excluded (*k* = 46, *d* _random_ = -0.24, 95% CI -0.39, -0.09, *I^2^* = 93.38%) (Online Resource: Supplementary Table II).

**Figure I.**
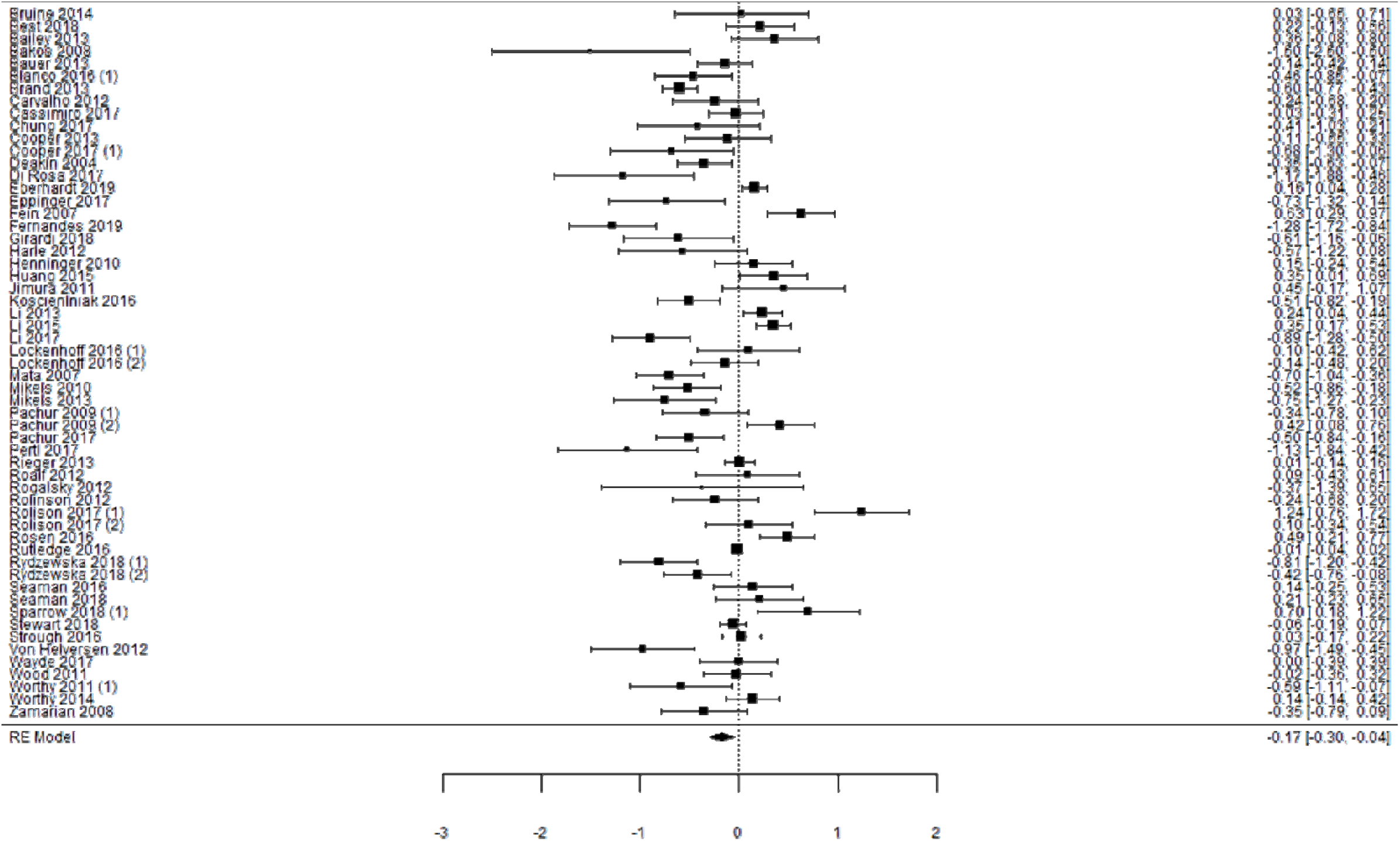
Forest plot showing individual and summary effects for 55 studies on age and overall decision-making. *Note.* Cohen’s d effects summary effects and 95% CI represented, with positive effects representing better decision making in older adults as compared to younger adults. A restricted maximum likelihood model was employed.

### Older compared to younger adults, decision-making domain

Meta-analysis by decision-making domain (financial, social or health related decisions) found combined estimates showing poorer financial (*k* = 36, *d* _random_ = -0.25, 95% CI -0.45, -0.04, *I^2^* = 94.68%) and social decision-making (*k* = 14, *d* _random_ = -0.38, 95% CI - 0.72, -0.03, *I^2^* = 92.9%) in older adults as compared to younger adults, as shown in Online Resource: Supplementary Fig. II). However, these were not significant for health decision-making (*k* = 5, *d* _random_ = 0.34, 95% CI -0.31, 0.99, *I^2^* = 95.02%; Online Resource: Supplementary Fig. II). Task demands, specifically risk and independent variable type moderated the effect of age on social decision-making. Post-hoc sensitivity analyses excluding one study of risky choice resulted in non-significant negative summary effect, (*d* _random_ = -0.26, 95% CI -0.54, 0.25, *I^2^* = 92.84%). However, the exclusion of studies which employed age as continuous variable did not materially affect the results, (*d* _random_ = -0.59, 95% CI -1.01, -0.18, *I^2^* = 88.5%). There were insufficient studies (*k* = 2) to allow for meaningful meta-analysis of safety-related decision-making. Moderators including mean sample age, task type, additional task demands, and adjustment for socio-economic factors did not materially alter the results for the financial and health decision-making domains (see Online Resource: Supplementary Table IV).

### Assessment of bias

Included studies were broadly similar in their overall reviewer-rated risk of bias (Online Resource: Supplementary Table IV). High risk of bias generally resulted from inadequate sample size, no a priori sample size calculation, a lack of statistical adjustment for potential confounders, and the use of a cross-sectional design. On the other hand, many studies reported using validated decision-making tasks (e.g., IGT, UG, GDT, BART), recruiting roughly even numbers of older and younger adults and exposing them to the same experimental conditions.

## Discussion

This review systematically synthesizes the available evidence comparing decision-making in earlier and later life across three real-world decision-making domains. More than half of the studies included in the systematic review provided support for an age-related change in decision-making with older adults performing more poorly when judged against achieving prespecified goals in standard decision tasks. Differences in studies could be explained to some extent by the nature of the decision tasks. In particular, positive associations between age and decision-making were observed across decision-tasks that involved altruism or prosocial considerations (Bailey et al. 2013; Beadle et al. 2012; Rosen et al. 2016; Sparrow and Spaniol 2018), response to risk (Rolison et al. 2017), inferencing (Pachur et al. 2009), resistance to sunk cost (Strough et al. 2015) and real-world credit scores (Li et al. 2015). Age differences in overall and financial decision-making were robust, with older adults performing more poorly than their younger counterparts although individual studies estimates varied widely.

Previous reviews (Best and Charness 2015; Thornton and Dumke 2005) have identified interpersonal considerations as an important moderator of age-differences in decision-making. Thornton and Dumke observed that age-associated declines in everyday problem-solving and decision-making effectiveness were attenuated when decisions had interpersonal significance. In their review, older adults appear to perform more favourably when outcomes resulted in mutual benefit. Broader research has also shown that older age is linked to increased altruism, better interpersonal problem solving, and prioritisation of social considerations in decision-making (Best and Charness 2015; Blanchard-Fields et al. 2007), Best and Charness similarly highlighted the important interplay between decisions involving interpersonal factors and age. Older adults were less risk-seeking than younger adults in positively framed morality-based scenario conditions.

Whilst the focus of our review was to summarise the existing evidence rather than to delve into the potential drivers behind the differences, we highlight two potential explanatory theories that may provide insight to help to explain age differences in decision outcomes.

The Selection, Optimisation and Compensation model (SOC) and Socioemotional Selectivity Theory (SST). SOC hypothesises that individuals use selection (elective and loss-based selection), optimisation and compensation based strategies to function effectively in different situations (Baltes and Carstensen 1996). Increasing age may be associated with better use of strategies for dealing with changing situations. However, in older age, structural changes in the prefrontal cortex and associated declines in cognitive abilities and processing speed may mean greater dependence on strategies like selection and optimisation, and less use of effortful, deliberative decision-making processes (Kahneman 2003). Reduced reliance upon deliberative processing may have stronger effects in financial decision-making tasks, particularly those completed under explicit risk conditions (Bangma et al. 2017; Ellen Peters et al. 2007). SST by contrast posits that shrinking time horizons in later life lead older adults to increasingly prioritise emotionally meaningful goals with short term rewards (e.g., charity) over goals facilitating self-sustenance and long-term payoff (e.g., building wealth) (Lockenhoff et al. 2016). The prioritisation of socio-emotional goals may result in lower motivation to perform on tasks with individual financial reward and increased drive to act in favour of a decision partner, even if this strategy results in negative individual consequences. The role of age-related neurobiological changes on individual and prosocial decision-making is less clear (Han et al. 2015; James et al. 2015).

In summary, older adults perform marginally worse that younger adults overall in decision-making tasks with a level of applicability to real-world outcomes. This appears to be driven by an overrepresentation of financial compared to social and health decision-making studies and greater consistency across summary effects within the financial domain. The small number of studies that have explored age-differences in social, health and safety decision-making have produced inconsistent results.

### Strengths and Limitations

There are several methodological strengths and limitations of the present research. The final search terms were selected based on sensitivity and specificity and applied to three leading scientific databases; however, we did not include the ‘grey’ literature and it is therefore possible some studies were not captured. To minimise this risk, the reference list of existing systematic reviews was also searched. The inclusive eligibility criteria led to heterogeneity in the study populations but allowed us to capture the widest possible scope of evidence resulting in a substantial body of data with representation across financial, social, health and non-risky decisions which was further explored using a series of potential moderators (Doi et al. 2011). However, we were inevitably limited by the published evidence, the types of decision-making paradigms that were available in the literature and the quality of constituent studies. One possible alternative for future consideration would be to conduct an individual participant data-analysis, which, while time-intensive, allows for verification of published results, the inclusion of unreported findings, and standardisation of statistical analysis albeit at the cost of standardising disparate outcomes. Evidence on decision-making response tendencies or preferences that could not be categorised as better or worse were deemed as less relevant to the interpretation of the results, and thus excluded.

Overall, the evidence base in this area is also lacking published prospective data. While experimental research facilitates greater methodological rigour, replicability, and a closer approximation of causal relationships, prospective studies have been argued to be better suited for investigating gradual and long-term processing changes across the life course. Furthermore, the scope of this high-level synthesis on age differences in financial, social, health and safety decision-making prohibited investigation into the mechanisms underlying observed age effects. Nevertheless, this review summarises valuable high-level evidence regarding age differences in decision-making, illuminates methodological issues and knowledge gaps and points to future directions for the field.

### Future Directions

As highlighted by the overrepresentation of financial decision-making evidence, more studies of social, health and safety-related decision-making with greater ecological validity would be a welcome addition to the literature. Secondly, the consistent application of validated decision-making measures across future studies will be critical to overcoming issues of generalisability, and replicability. Relative to clinical or neuroimageing tools, decision-making tasks often require minimal supervision and clinical interpretation, which may enhance their utility and accessibility across different contexts. Thirdly, synthesising the evidence from studies of decisions outcomes that are not categorisable as better or worse (e.g., preferences for advertisements with more or less information) and the mechanisms underpinning observed age differences in decision-making would build upon the work of the present review. There is also a clear need for further in-depth reviews of the potential mechanisms, which, whilst beyond the scope of this work would help to further disentangle the results from the experimental studies and help in the design of future data collection.

Fourthly, the generation of prospective data will be necessary to understand how decision processes change with age on an intra-individual level. The inclusion of decision-making measures within ongoing longitudinal studies, albeit with the need to take account of learning effects, may be one efficient way to achieve this research goal. Lastly, consensus on terminology and operationalising decision-making across gerontological and psychological research will strengthen the interpretability and comparability of future findings.

## Statements and declarations

### Impact Statement

Age differences in decision-making have important implications for the ways that decision-making is supported, and the information that is provided to adults at different ages. This is particularly relevant for decisions that may have significant health, financial or social consequences. Currently our policies and practices do not take explicit account of the potential changes in decision-making ability with ageing. Our review speaks to the importance of taking this into account and highlights the need for further research.

### Ethical statement

An ethics statement was not required for this study type, no human or animal subjects or materials were used.

## Data availability statement

All data used or generated in this study are from previously published work. Further enquiries can be directed to the corresponding author.

