## Supplementary Information for "Ageing and Decision-making: A systematic review and meta-analysis"

a. School of Psychology, University of New South Wales, Sydney, Australia
b. Neuroscience Research Australia, Sydney, Australia
c. UNSW Ageing Futures Institute, University of New South Wales, Sydney, Australia
d. The George Institute for Global Health, Australia

***Corresponding author**:

Nicole Ee

### Supplementary Fig. I PRISMA flow diagram

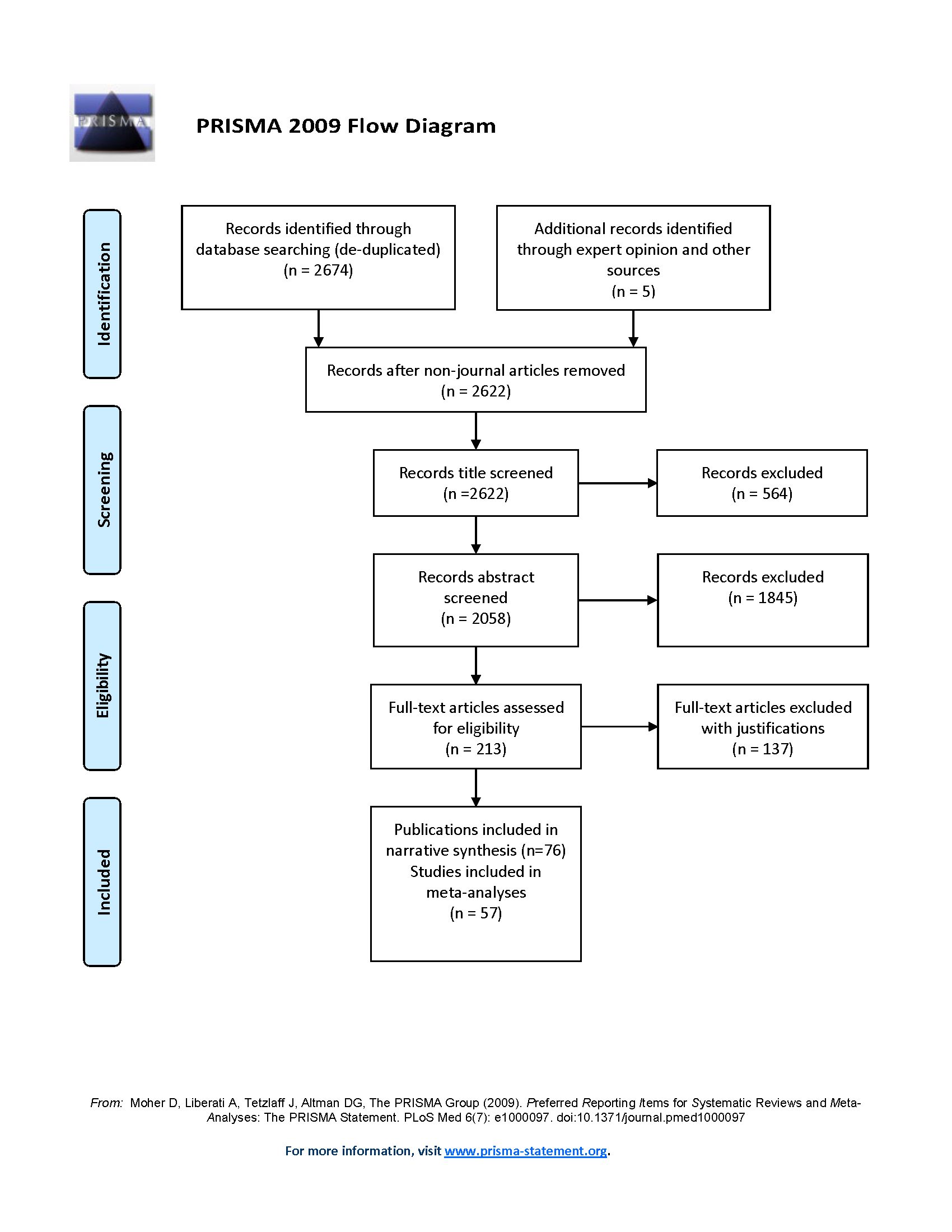

*Note.* Flow chart from Moher D, Liberati A, Tetzlaff J, Altman DG (2009) Preferred Reporting Items for Systematic Reviews and Meta-Analyses: the PRISMA statement. PLoS Med 6(7):e1000097. doi:10.1371/journal.pmed1000097.

Supplementary Fig. II
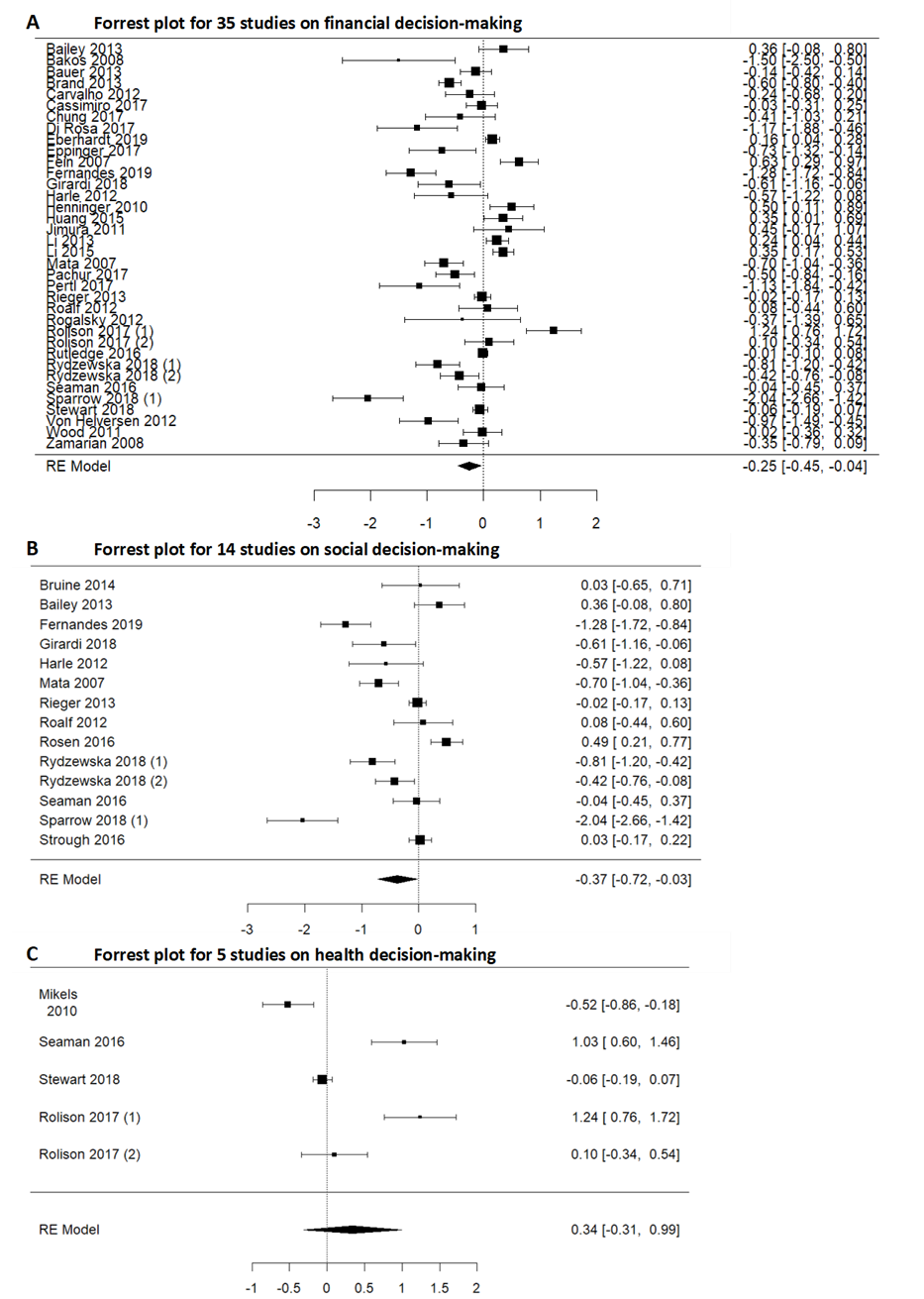
 Forrest plots showing individual and summary effects for financial (A), social (B) and health (C) decision-making studies

*Note.* Cohen’s d effects summary effects and 95% CI represented, with positive effects representing better decision making in older adults as compared to younger adults. A restricted maximum likelihood model was employed.

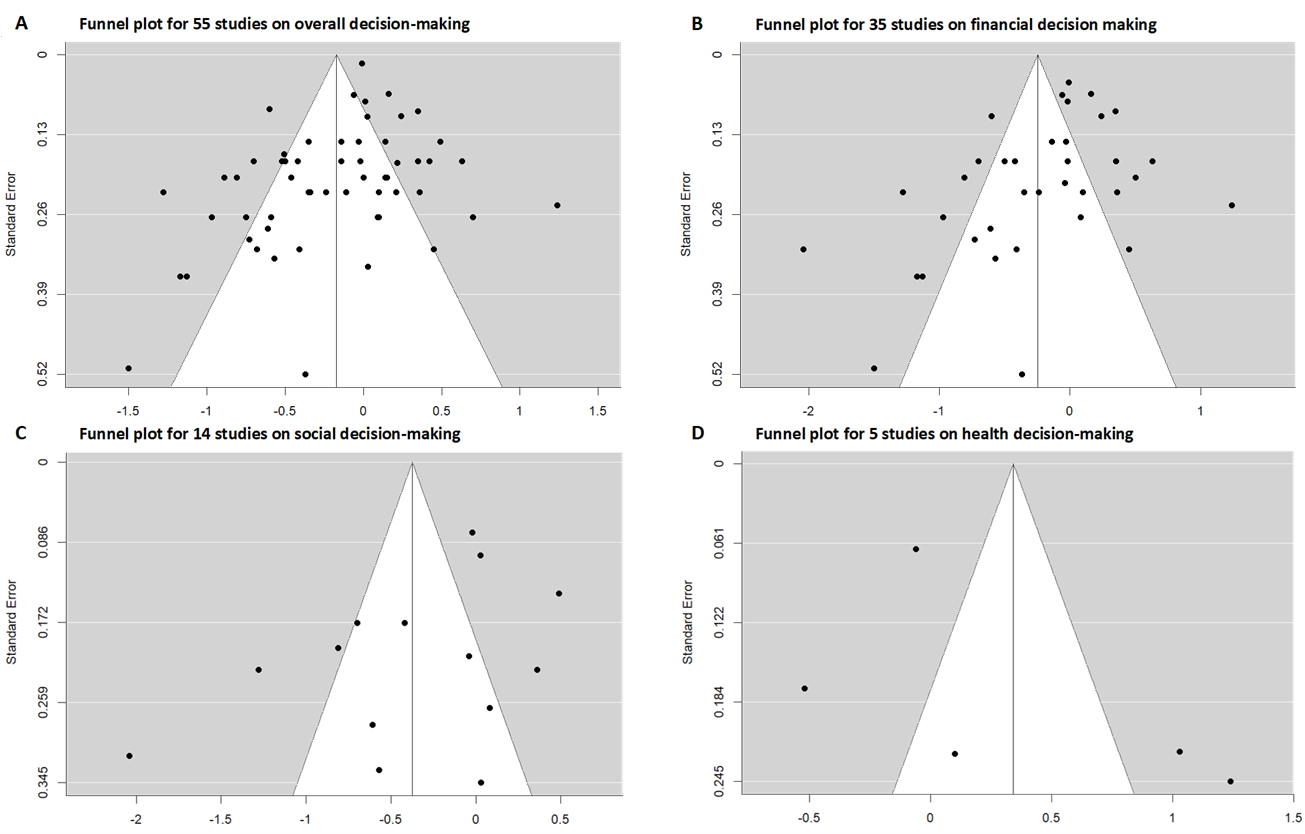

### Supplementary Fig. III Funnel plots for meta-analysis of overall (A), financial (B), social (C) and health decision-making (D)

*Note.* Cohen’s d effects summary effects and 95% CI represented, with positive effects representing better decision making in older adults as compared to younger adults. A restricted maximum likelihood model was employed.

### Supplementary Table I

### *Studies excluded and justification for exclusion*

| Reference | Justification |
| --- | --- |
| Basharat A, Adams MS, Staines WR, Barnett-Cowan M. (2018) Simultaneity and Temporal Order Judgments Are Coded Differently and Change with Age: an event-related potential study. Front Integr Neurosci 12(15):15. doi:10.3389/fnint.2018.00015 | Temporal order judgement and simultaneity judgements, decision-making outcomes not quantifiable as positive or negative. |
| Bedard G, Barnett-Cowan M. (2016) Impaired timing of audiovisual events in the elderly. Exp Brain Res 234(1):331-340. doi:10.1007/s00221-015-4466-7 | Temporal order judgement and simultaneity judgements, decision-making outcomes not quantifiable as positive or negative. |
| Beitz KM, Salthouse TA, Davis HP (2014) Performance on the Iowa Gambling Task: from 5 to 89 years of age. J Exp Psychol Gen 143(4):1677-1689. | Outcome is learning as a function of loss frequency and net outcome of each but decision-making outcomes not quantifiable as positive or negative. |
| Best R, Charness N (2015) Age differences in the effect of framing on risky choice: a meta-analysis. Psychol Aging 30(3):688. | Meta-analysis of effect of risky choice. |
| Bjälkebring P, Västfjäll D, Johansson B (2013) Regulation of Experienced and Anticipated Regret for Daily Decisions in Younger and Older Adults in a Swedish One-Week Diary Study GeroPsych 26(4):233-241. doi:10.1024/1662-9647/a000102 | Regret and regret regulation in relations to decisions, decision-making outcomes not quantifiable as positive or negative. |
| Boettger S, Bergman M, Jenewein J, Boettger S (2016) Advanced age and decisional capacity: the effect of age on the ability to make health care decisions. Arch Gerontol Geriatr 66:211-217. | Outcome is decision-making capacity but decision-making outcomes not quantifiable as positive or negative. |
| Brand M, Markowitsch HJ (2010) Aging and decision-making: a neurocognitive perspective. Gerontology 56(3):319-324. doi:10.1159/000248829 | Narrative review. |
| Breland S (2011) Later life decision-making: experiential adult learning and successful aging. The University of Southern Mississippi. | Dissertation, non-peer reviewed. |
| Brown SB, Ridderinkhof KR (2009) Aging and the neuroeconomics of decision making: a review. Cogn Affect Behav Neurosci 9(4):365-379. doi:10.3758/CABN.9.4.365 | Review. |
| Brown SW, Johnson TM, Sohl ME, Dumas MK (2015) Executive attentional resources in timing: effects of inhibitory control and cognitive aging. Journal of J Exp Psychol Hum Percept Perform 41(4):1063-1083. doi:10.1037/xhp0000078 | Decision-making outcomes not quantifiable as positive or negative. |
| Bruine de Bruin W, Parker AM, Fischhoff B (2012) Explaining adult age differences in decision‐making competence. J Behav Decis Mak 25(4):352-360. | On decision-making competence, decision-making outcomes not quantifiable as positive or negative. |
| Bruine de Bruin W, Parker AM, Strough J (2016) Choosing to be happy? Age differences in "maximizing" decision strategies and experienced emotional well-being. Psychol Aging 31(3):295-300. doi:10.1037/pag0000073 | On maximising and experience of emotional well-being, decision-making outcomes not quantifiable as positive or negative. |
| Carpenter SM, Peters E, Västfjäll D, Isen AM (2013) Positive feelings facilitate working memory and complex decision making among older adults. Cogn Emot 27(1):184-192. | No results by age or age comparison. |
| Castel AD, Friedman MC, McGillivray S, Flores CC, Murayama K, Kerr T, Drolet A (2016) I owe you: age-related similarities and differences in associative memory for gains and losses. Neuropsychol Dev Cogn B Aging Neuropsychol Cogn 23(5):549-565. doi:10.1080/13825585.2015.1130214 | Outcome is memory, decision-making outcomes not quantifiable as positive or negative. |
| Castel AD, Humphreys KL, Lee SS, Galván A, Balota DA, McCabe DP (2011) The development of memory efficiency and value-directed remembering across the life span: a cross-sectional study of memory and selectivity. Dev Psychol 47(6):1553. | Outcome is memory, decision-making outcomes not quantifiable as positive or negative. |
| Castel AD, Rossi AD, McGillivray S (2012) Beliefs about the "hot hand" in basketball across the adult life span. Psychol Aging 27(3):601-605. doi:10.1037/a0026991 | Outcome is memory, decision-making outcomes not quantifiable as positive or negative. |
| Chapman RM, Gardner MN, Mapstone M, Klorman R, Porsteinsson AP, Dupree HM (2016) ERP C250 shows the elderly (cognitively normal, Alzheimer's disease) store more stimuli in short-term memory than Young Adults do. Clin Neurophysiol 127(6):2423-2435. doi:10.1016/j.clinph.2016.03.006 | Outcome is memory, decision-making outcomes not quantifiable as positive or negative. |
| Chen X, Rutledge RB, Brown HR, Dolan RJ, Bestmann S, Galea JM. (2018) Age-dependent Pavlovian biases influence motor decision-making. PLoS Comput Biol 14(7):e1006304. doi:10.1371/journal.pcbi.1006304 | On motor decision-making, decision-making outcomes not quantifiable as positive or negative. |
| Chen Y, Ma X, Pethtel O (2011) Age differences in trade-off decisions: older adults prefer choice deferral. Psychol Aging 26(2):269-273. doi:10.1037/a0021582 | Outcome is decision avoidance, decision-making outcomes not quantifiable as positive or negative. |
| Clark R, Hazeltine E, Freedberg M, Voss MW (2018) Age differences in episodic associative learning. Psychol Aging 33(1):144-157. doi:10.1037/pag0000234 | Outcome is choice reaction time. |
| Crumlin J (2005) Aging and the influence of emotion on judgment. University of Denver. | Influence of mood on risk judgment, decision-making outcomes not quantifiable as positive or negative. |
| Del Missier F, Mäntylä T, Hansson P, Bruine de Bruin W, Parker AM, Nilsson LG (2013) The multifold relationship between memory and decision making: an individual-differences study. J Exper Psychol Learn Mem Cogn 39(5):1344. | decision-making outcomes not quantifiable as positive or negative. |
| Denburg NL, Recknor EC, Bechara A, Tranel D (2006) Psychophysiological anticipation of positive outcomes promotes advantageous decision-making in normal older persons. Int J Psychophysiol 61(1):19-25. doi:10.1016/j.ijpsycho.2005.10.021 | Only older adults’ sample, comparing impaired versus unimpaired older adults on the IGT. No age comparison. |
| Depping MK, Freund AM (2011) Normal Aging and Decision Making: the role of motivation. Human Development 54(6):349-367. doi:10.1159/000334396 | Review. |
| Depping MK, Freund AM (2013) When choice matters: task-dependent memory effects in older adulthood. Psychol Aging 28(4):923-936. doi:10.1037/a0034520 | On information processing and impact on memory, decision-making outcomes not quantifiable as positive or negative. |
| Dully J, McGovern DP, O’Connell RG (2018) The impact of natural aging on computational and neural indices of perceptual decision making: a review. Behav Brain Res 355:48-55. doi:10.1016/j.bbr.2018.02.001 | Review. |
| Dunlosky J, Kubat-Silman AK, Hertzog C (2003) Effects of aging on the magnitude and accuracy of quality-of-encoding judgments. Am J Psychol 116(3):431-454. | Outcome is quality of encoding judgements, decision-making outcomes not quantifiable as positive or negative. |
| Duverne S, Lemaire P (2005) Arithmetic split effects reflect strategy selection: an adult age comparative study in addition comparison and verification tasks. Can J Exp Psychol 59(4):262-278. | Outcome is split effects, decision-making outcomes not quantifiable as positive or negative. |
| Eitner A (1973) Influence of aging on information processing. Probleme und Ergebnisse der Psychologie 47:15-43. | On information processing, decision-making outcomes not quantifiable as positive or negative. |
| English T, Carstensen LL (2015) Does positivity operate when the stakes are high? Health status and decision making among older adults. Psychol Aging 30(2):348-355. | On positivity, decision-making outcomes not quantifiable as positive or negative. |
| Eppinger B, Hämmerer D, Li SC (2011) Neuromodulation of reward-based learning and decision making in human aging. Ann N Y Acad Sci 1235:1-17. | Review. |
| Eppinger B, Heekeren HR, Li SC (2015) Age-related prefrontal impairments implicate deficient prediction of future reward in older adults. Neurobiol Aging 36(8):2380-2390. doi:10.1016/j.neurobiolaging.2015.04.010 | Outcome is age-related learning in choice decision-making but decision-making outcomes not quantifiable as positive or negative. |
| Eppinger B, Nystrom L, Cohen JD (2011) Age-Related Changes in the Neural Correlates of Intertemporal Decision-Making and Reward-Based Learning. Journal of Psychophysiology 25 11-11. | Supplementary abstract only, no peer-review paper found. |
| Erber JT, Szuchman LT, Prager IG (2001) Ain't misbehavin': the effects of age and intentionality on judgments about misconduct. Psychol Aging 16(1):85-95. doi:Doi 10.1037/0882-7974.16.1.85 | Outcome is perception of older and younger individuals, decision-making outcomes not quantifiable as positive or negative. |
| Fern CS, Goncalves AR, Ferreira-Santos F, Barbosa F, Martins, IP, Marques-Teixeira J (2019) The Aging Social brain: neural and behavioral age-related changes in social cognition and decision-making. University of Porto. | Review. |
| Fernandes C, Pasion R, Goncalves AR, Ferreira-Santos F, Barbosa F, Martins IP, Marques-Teixeira J (2018) Age differences in neural correlates of feedback processing after economic decisions under risk. Neurobiol Aging 65:51-59. doi:10.1016/j.neurobiolaging.2018.01.003 | Outcome is risk aversion, decision-making outcomes not quantifiable as positive or negative. |
| Finucane ML, Gullion CM (2010) Developing a tool for measuring the decision-making competence of older adults. Psychol Aging 25(2):271-288. doi:10.1037/a0019106 | Outcome is decision-making competence, decision-making outcomes not quantifiable as positive or negative. |
| Finucane ML, Mertz CK, Slovic P, Schmidt ES (2005) Task complexity and older adults' decision-making competence. Psychol Aging 20(1):71-84. doi:10.1037/0882-7974.20.1.71 | Outcome is decision-making competency and comprehension, decision-making outcomes not quantifiable as positive or negative. |
| Finucane ML, Slovic P, Hibbard JH, Peters E, Mertz CK, MacGregor DG (2002) Aging and decision‐making competence: an analysis of comprehension and consistency skills in older versus younger adults considering health‐plan options. J Behav Decis Mak 15(2):141-164. | Outcome is decision-making competence, decision-making outcomes not quantifiable as positive or negative. |
| Fitzgerald JM (2000) Younger and older jurors: the influence of environmental supports on memory performance and decision making in complex trials. J Gerontol B Psychol Sci Soc Sci 55(6):P323-331. doi:10.1093/geronb/55.6.p323 | On juror memory, decision-making outcomes not quantifiable as positive or negative. |
| Forbes S, Hoffart N (1998) Elders' decision making regarding the use of long-term care services: a precarious balance. Qual Health Res 8(6):736-750. doi:10.1177/104973239800800602 | Decision-making qualitatively measured, decision-making outcomes not quantifiable as positive or negative. |
| Forstmann BU, Tittgemeyer M, Wagenmakers EJ, Derrfuss J, Imperati D, Brown S (2011) The speed-accuracy tradeoff in the elderly brain: a structural model-based approach. J Neurosci 31(47):17242-17249. doi:10.1523/JNEUROSCI.0309-11.2011 | Outcome is reaction time, decision-making outcomes not quantifiable as positive or negative. |
| Fu L, Maes JH, Varma S, Kessels RP, Daselaar SM (2017) Effortful semantic decision-making boosts memory performance in older adults. Memory 25(4):544-549. doi:10.1080/09658211.2016.1193204 | Outcome is episodic memory, decision-making outcomes not quantifiable as positive or negative. |
| Gajewski PD, Falkenstein M (2013) Decision Making and Error Monitoring in Elderly Employees: neurobehavioral correlates of work-related deterioration and training-induced improvement. J Psychophysiology 27:22-22. | No age comparison, only older adults. |
| Gamble KJ, Boyle PA, Yu L, Bennett DA (2015) Aging and Financial Decision Making. Management Science 61(11):2603-2610. doi:10.1287/mnsc.2014.2010 | Outcome is financial literacy and cognition, decision-making outcomes not quantifiable as positive or negative. |
| Gandini D, Lemaire P, Dufau S (2008) Older and younger adults' strategies in approximate quantification. Acta Psychol:129(1):175-189. doi:10.1016/j.actpsy.2008.05.009 | On estimation, decision-making outcomes not quantifiable as positive or negative. |
| Goh JO, Su YS, Tang YJ, McCarrey AC, Tereshchenko A, Elkins W, Resnick SM (2016) Frontal, Striatal, and Medial Temporal Sensitivity to Value Distinguishes Risk-Taking from Risk-Aversive Older Adults during Decision Making. J Neurosci 36(49):12498-12509. doi:10.1523/JNEUROSCI.1386-16.2016 | Outcome is decision threshold for risky decisions, decision-making outcomes not quantifiable as positive or negative. |
| Gold BT, Andersen AH, Jicha GA, Smith CD (2009) Aging influences the neural correlates of lexical decision but not automatic semantic priming. Cereb Cortex 19(11):2671-2679. doi:10.1093/cercor/bhp018 | Outcome is lexical decision-making, but decision-making outcomes not quantifiable as positive or negative. |
| Halfmann KM (2015) Emotion and decision-making in the aging brain. The University of Iowa. | Dissertation, non-peer reviewed. |
| Halfmann K, Hedgcock W, Bechara A, Denburg NL (2014) Functional neuroimaging of the Iowa Gambling Task in older adults. Neuropsychology 28(6):870-880. doi:10.1037/neu0000120 | No results by age or age comparison. |
| Han SD, Boyle PA, James BD, Yu L, Barnes LL, Bennett DA (2016) Discrepancies between cognition and decision making in older adults. Aging Clin Exp Res 28(1):99-108. doi:10.1007/s40520-015-0375-7 | Outcome is discrepancies between cognition and decision performance, no separate decision-making outcomes that are quantifiable as positive or negative. |
| Hannah SD, Allan LG, Young ME (2012) Age differences in contingency judgement linked to perceptual segregation. Q J Exp Psychol (Hove):65(6):1195-1213. doi:10.1080/17470218.2011.649293 | Outcome is contingency judgements, decision-making outcomes not quantifiable as positive or negative. |
| Hanoch Y, Wood S, Rice T (2007) Bounded rationality, emotions and older adult decision making: not so fast and yet so frugal. Human Development 50(6):333-358. | Discussion paper. |
| Harty S, Murphy PR, Robertson IH, O'Connell RG (2017) Parsing the neural signatures of reduced error detection in older age. NeuroImage 161:43-55. doi:10.1016/j.neuroimage.2017.08.032 | Outcome is error awareness, decision-making outcomes not quantifiable as positive or negative. |
| Hatta T, Hatta T, Iwahara A, Hatta J, Nagahara N, Ito E, Fujiwara K, Hotta C (2015) Relations among higher brain function, trust, and gullibility in middle-aged and elderly people. The Japanese Journal of Psychology 85(6):540-548. | Outcome is cognitive processing and susceptibility to deceit, decision-making outcomes not quantifiable as positive or negative. |
| Hertzog C, Sinclair SM, Dunlosky J (2010) Age differences in the monitoring of learning: cross-sectional evidence of spared resolution across the adult life span. Dev Psychol 46(4):939. | Outcome is learning, no separate decision-making outcomes that are quantifiable as positive or negative. |
| Hess TM, Smith BT (2014) Aging and the impact of irrelevant information on social judgments. Psychol Aging 29(3):542-553. doi:10.1037/a0036730 | Outcome is processing of relevant and irrelevant information in job evaluation but decision-making outcomes not quantifiable as positive or negative. |
| Hess TM, Leclerc CM, Swaim E, Weatherbee SR (2009) Aging and everyday judgments: the impact of motivational and processing resource factors. Psychol Aging 24(3):735. | Outcome is learning, decision-making outcomes not quantifiable as positive or negative. |
| Hess TM, Queen TL, Ennis GE (2012) Age and self-relevance effects on information search during decision making. J Geron B Psychol Sci Soc Sci 68(5):703-711. | Outcomes related to strategy use, decision-making outcomes not quantifiable as positive or negative. |
| Hoppmann CA, Blanchard-Fields F (2010) Goals and everyday problem solving: manipulating goal preferences in young and older adults. Dev Psychol 46(6):1433-1443. doi:10.1037/a0020676 | Outcome is link between goal and problem-solving strategy, no separate decision-making outcomes that are quantifiable as positive or negative. |
| Hosseini SH, Rostami M, Yomogida Y, Takahashi M, Tsukiura T, Kawashima R (2010) Aging and decision making under uncertainty: behavioral and neural evidence for the preservation of decision making in the absence of learning in old age. Neuroimage 52(4):1514-1520. | Outcome is self-reported decisional strategies, decision-making outcomes not quantifiable as positive or negative. |
| Hsieh S, Wu M (2010) Age differences in switching the relevant stimulus dimensions in a speeded same-different judgment paradigm. Acta Psychol (Amst):135(2):140-149. doi:10.1016/j.actpsy.2010.05.010 | Outcome is same-different judgement and reaction time, no separate decision-making outcomes that are quantifiable as positive or negative. |
| Isella V, Mapelli C, Morielli N, Pelati O, Franceschi, M, Appollonio IM (2008) Age-related quantitative and qualitative changes in decision making ability. Behav Neurol 19(1-2):59-63. | No results by age or age comparison in healthy adults. |
| Jacus JP, Bayard S, Raffard S, Bonnoron S, Gely-Nargeot MC (2012) [Decision-making in normal and pathological aging]. Geriatrie et psychologie neuropsychiatrie du vieillissement 10(4):437-444. doi:10.1684/pnv.2012.0371 | Discussion paper. |
| Johnson MM (1993) Thinking About Strategies during, before, and after Making a Decision. Psychol Aging 8(2):231-241. doi:Doi 10.1037/0882-7974.8.2.231 | Outcome is decision strategies, decision-making outcomes not quantifiable as positive or negative. |
| Kardos Z, Kóbor A, Takács Á, Tóth B, Boha R, File B, Molnár M (2016) Age-related characteristics of risky decision-making and progressive expectation formation. Behav Brain Res 312:405-414. doi:10.1016/j.bbr.2016.07.003 | On risk aversion and exploratory behaviour, decision-making outcomes not quantifiable as positive or negative. |
| Kim S, Hasher L (2005) The attraction effect in decision making: superior performance by older adults. Q J Exp Psychol A 58(1):120-133. doi:10.1080/02724980443000160 | Outcome is choice consistency, decision-making outcomes not quantifiable as positive or negative. |
| Kim S, Healey MK, Goldstein D, Hasher L, Wiprzycka UJ (2008) Age differences in choice satisfaction: a positivity effect in decision making. Psychol Aging 23(1):33-38. doi:10.1037/0882-7974.23.1.33 | Outcome is decision satisfaction, decision-making outcomes not quantifiable as positive or negative. |
| Kliegel M, Ramuschkat G, Martin M (2003) Exekutive funktionen und prospektive gedachtnisleistung im alter - Eine differentielle analyse von ereignisund zeitbasierter prospektiver gedachtnisleistung. Zeitschrift für Gerontologie und Geriatrie 36(1):35-41. doi:10.1007/s00391-003-0081-5 | Non-English copy of publication; English version excluded as outcome is memory, decision-making outcomes not quantifiable as positive or negative. |
| Kliegel M, Ramuschkat G, Martin M (2003) Executive functions and prospective memory performance in old age: an analysis of event-based and time-based prospective memory. Zeitschrift fur Gerontologie und Geriatrie 36(1):35-41. | Outcome is memory, decision-making outcomes not quantifiable as positive or negative. |
| Kovalchik S, Camerer CF, Grether DM, Plott CR, Allman JM (2005) Aging and decision making: a comparison between neurologically healthy elderly and young individuals. J Econ Behav Organ 58(1):79-94. doi:10.1016/j.jebo.2003.12.001 | No statistical analysis conducted on age group comparison or results by age. |
| Kubo-Kawai N, Kawai N (2010) Elimination of the enhanced Simon effect for older adults in a three-choice situation: ageing and the Simon effect in a go/no-go Simon task. Q J Exp Psychol (Hove):63(3):452-464. doi:10.1080/17470210902990829 | Go-no-go task testing Simon effect, decision-making outcomes not quantifiable as positive or negative. |
| Kurnianingsih YA, Sim SK, Chee MW, Mullette-Gillman OD (2015) Aging and loss decision making: increased risk aversion and decreased use of maximizing information, with correlated rationality and value maximization. Front Hum Neurosci 9:280. | Outcome risk aversion and choice strategy but decision-making outcomes not quantifiable as positive or negative. |
| Labudda K, Woermann FG, Mertens M, Pohlmann-Eden B, Markowitsch HJ, Brand M (2008) Neural correlates of decision making with explicit information about probabilities and incentives in elderly healthy subjects. Exp Brain Res 187(4):641-650. doi:10.1007/s00221-008-1332-x | Outcome is neural activation, decision-making outcomes not quantifiable as positive or negative. |
| Li Y, Baldassi M, Johnson EJ, Weber EU (2013) Complementary cognitive capabilities, economic decision making, and aging. Psychol Aging 28(3):595. | On temporal discounting loss aversion, financial literacy, anchoring but decision-making outcomes not quantifiable as positive or negative. |
| Lichtenberg PA, Ficker LJ, Rahman-Filipiak A (2016) Financial decision-making abilities and financial exploitation in older African Americans: preliminary validity evidence for the Lichtenberg Financial Decision Rating Scale (LFDRS) J Elder Abuse Negl 28(1):14-33. doi:10.1080/08946566.2015.1078760 | Study on validity of the Lichtenberg Financial Decision Rating Scale (LFDRS) in African Americans. Outcome is decision-making ability, decision-making outcomes not quantifiable as positive or negative. |
| Lim KT, Yu R (2015) Aging and wisdom: age-related changes in economic and social decision making. Front Aging Neurosci 7(120):120. doi:10.3389/fnagi.2015.00120 | Review. |
| Han-hui LI, Yan-yan AN, Hui-min LI, Zhen WE, Xing-ting ZH, Hui-jie LI (2014) Development and aging of decision-making rationality under risk framework. Contemporary Neurology & Neurosurgery 14(3):186-191. | On framing effect, decision-making outcomes not quantifiable as positive or negative. |
| Löckenhoff CE, Carstensen LL (2007) Aging, emotion, and health-related decision strategies: motivational manipulations can reduce age differences. Psychol Aging 22(1):134-146. doi:10.1037/0882-7974.22.1.134 | Decision-making outcomes not quantifiable as positive or negative. |
| Löckenhoff CE, Carstensen LL (2008) Decision strategies in health care choices for self and others: older but not younger adults make adjustments for the age of the decision target. J Gerontol B Psychol Sci Soc Sci 63(2):P106-P109. | Outcome is information review strategy and emotional response, decision-making outcomes not quantifiable as positive or negative. |
| Mares ML, Bartsch A, Bonus JA (2016) When meaning matters more: media preferences across the adult life span. Psychol Aging 31(5):513-531. doi:10.1037/pag0000098 | On media preferences across lifespan, decision-making outcomes not quantifiable as positive or negative. |
| Marschner A, Mell T, Wartenburger I, Villringer A, Reischies FM, Heekeren HR (2005) Reward-based decision-making and aging. Brain Res Bull 67(5):382-390. doi:10.1016/j.brainresbull.2005.06.010 | Review. |
| Mata R (2007) Understanding the aging decision maker. Human Development 50(6):359-366. doi:10.1159/000109836 | Narrative paper. |
| Mata R, Nunes L (2010) When less is enough: cognitive aging, information search, and decision quality in consumer choice. Psychol Aging 25(2):289-298. doi:10.1037/a0017927 | Meta-analysis. |
| Mata R, Pachur T, Von Helversen B, Hertwig R, Rieskamp J, Schooler L (2012) Ecological rationality: a framework for understanding and aiding the aging decision maker. Front Neurosci 6(6):19. doi:10.3389/fnins.2012.00019 | Review. |
| Mather M, Mazar N, Gorlick MA, Lighthall NR, Burgeno J, Schoeke A, Ariely D (2012) Risk preferences and aging: the “certainty effect” in older adults' decision making. Psychol Aging 27(4):801. | On certainty effect, decision-making outcomes not quantifiable as positive or negative. |
| Meyer BJ, Talbot AP, Ranalli C (2007) Why older adults make more immediate treatment decisions about cancer than younger adults. Psychol Aging 22(3):505-524. | Outcome is immediacy of medical decision-making, decision-making outcomes not quantifiable as positive or negative. |
| Moberg PJ, Rick JH (2008) Decision-making capacity and competency in the elderly: a clinical and neuropsychological perspective. NeuroRehabilitation 23(5):403-413. | Review. |
| Mohr PN, Li SC, Heekeren HR (2010) Neuroeconomics and aging: neuromodulation of economic decision making in old age. Neurosci Biobehav Rev 34(5):678-688. | Review. |
| Moreno GL (2016) The effects of stress on decision making and the prefrontal cortex among older adults. University of Iowa. | Effect of acute stress on age-related changes in decision-making; no results by age or with age comparison. |
| Moret‐Tatay C, Lemus‐Zúñiga LG, Tortosa DA, Gamermann D, Vázquez‐Martínez A, Navarro‐Pardo E, Conejero JA (2017) Age slowing down in detection and visual discrimination under varying presentation times. Scand J Psychol 58(4):304-311. | On emotion regulation, decision-making outcomes not quantifiable as positive or negative. |
| Morrison JD, Reilly J (1986) An assessment of decision-making as a possible factor in the age-related loss of contrast sensitivity. Perception 15(5):541-552. doi:10.1068/p150541 | Outcome is signal detection and discrimination, decision-making outcomes not quantifiable as positive or negative. |
| Morsch P, Mirandola AR, Caberlon IC, Bós ÂJG (2017) Factors associated with health-related decision-making in older adults from Southern Brazil. Geriatr Gerontol Int 17(5):798-803. doi:10.1111/ggi.12788 | Outcome is whether decisions are conducted in isolation or with others, decision-making outcomes not quantifiable as positive or negative. |
| Mohr PN, Li SC, Heekeren HR (2008) Health economic choices in old age: interdisciplinary perspectives on economic decisions and the aging mind. Adv Health Econ Health Serv Res 20:227-270. | Book chapter. |
| Oertelt-Prigione S, Kendel F, Kaltenbach M, Hetzer R, Regitz-Zagrosek V, Baretti R (2013) Detection of gender differences in incomplete revascularization after coronary artery bypass surgery varies with classification technique. Biomed Res Int 2013(2):108475. doi:10.1155/2013/108475 | On effect of gender and classification technique on revascularisation patterns and mortality. |
| Pasion R, Goncalves AR, Fernandes C, Ferreira-Santos F, Barbosa F, Marques-Teixeira J (2017) Meta-Analytic Evidence for a Reversal Learning Effect on the Iowa Gambling Task in Older Adults. Front Psychol 8:1785. doi:10.3389/fpsyg.2017.01785 | Meta-analysis. |
| Pate DS, Margolin DI, Friedrich FJ, Bentley EE (2007) Decision-making and attentional processes in ageing and in dementia of the Alzheimer's type. Cognitive Neuropsychology 11(3):321-339. doi:10.1080/02643299408251978 | On differences between AD and normal adults, no report of decision-making within healthy adults by age. |
| Plaud JJ, Gillund B, Ferraro FR (2000) Signal detection analysis of choice behavior and ageing. Journal of Clinical Geropsychology 6(1):73-81. | Outcome is signal detection, decision-making outcomes not quantifiable as positive or negative. |
| Pornpattananangkul N, Kok BC, Chai J, Huang Y, Feng L, Yu R (2018) Choosing for you: diminished self-other discrepancies in financial decisions under risk in the elderly. Psychol Aging 33(6):871-891. doi:10.1037/pag0000284 | Outcome is loss aversion and risk aversion and self-other discrepancies but decision-making outcomes not quantifiable as positive or negative. |
| Qian Z, Guye KN, Masiello DJ, Ginger DS (2017) Dynamic Optical Switching of Polymer/Plasmonic Nanoparticle Hybrids with Sparse Loading. J Phys Chem B 121(5):1092-1099. doi:10.1021/acs.jpcb.7b00013 | Outcomes are riskiness of decision and reaction time, decision-making outcomes not quantifiable as positive or negative. |
| Queen TL, Hess TM, Ennis GE, Dowd K, Grühn D (2012) Information search and decision making: effects of age and complexity on strategy use. Psychol Aging 27(4):817. | Outcome information search strategy, decision-making outcomes not quantifiable as positive or negative. |
| Ratcliff R, Thapar A, McKoon G (2006) Ageing and individual differences in rapid two-choice decisions. Psychon Bull Rev 13(4):626-635. | On signal detection and discrimination, decision-making outcomes not quantifiable as positive or negative. |
| Ratcliff R, Thapar A, McKoon G (2007) Application of the diffusion model to two-choice tasks for adults 75-90 years old. Psychol Aging 22(1):56-66. doi:10.1037/0882-7974.22.1.56 | On signal detection and two-choice discrimination, decision-making outcomes not quantifiable as positive or negative. |
| Reed AE, Mikels JA, Löckenhoff CE (2013) Preferences for choice across adulthood: age trajectories and potential mechanisms. Psychol Aging 28(3):625-632. doi:10.1037/a0031399 | Outcome is choice preferences in a decision-making task, decision-making outcomes not quantifiable as positive or negative. |
| Reed AE, Mikels JA, Simon KI (2014) Older adults prefer less choice than young adults. Translational Issues in Psychological Science 1(S):5–10. | Outcome is choice preference, decision-making outcomes not quantifiable as positive or negative. |
| Reifegerste J, Hauer F, Felser C (2017) Agreement processing and attraction errors in ageing: evidence from subject-verb agreement in German. Neuropsychol Dev Cogn B Ageing Neuropsychol Cogn 24(6):672-702. doi:10.1080/13825585.2016.1251550 | On lexical processing, decision-making outcomes not quantifiable as positive or negative. |
| Reike D, Schwarz W (2019) Ageing effects on symbolic number comparison: no deceleration of numerical information retrieval but more conservative decision-making. Psychol Aging 34(1):4-16. doi:10.1037/pag0000272 | On numerical comparison, decision-making outcomes not quantifiable as positive or negative. |
| Samanez-Larkin GR, Knutson B (2015) Decision making in the ageing brain: changes in affective and motivational circuits. Nat Rev Neurosci 16(5):278-289. doi:10.1038/nrn3917 | Review. |
| Scheibe S, Sheppes G, Staudinger UM (2015) Distract or reappraise? Age-related differences in emotion-regulation choice. Emotion 15(6):677-681. doi:10.1037/a0039246 | On emotion regulation, decision-making outcomes not quantifiable as positive or negative. |
| Seaman, KL, Green MA, Shu S, Samanez-Larkin GR (2018) Individual differences in loss aversion and preferences for skewed risks across adulthood. Psychol Aging 33(4):654-659. doi:10.1037/pag0000261 | Outcome is loss aversion but decision-making outcomes not quantifiable as positive or negative. |
| Seaman KL, Howard DV, Howard Jr JH (2014) Adult age differences in learning on a sequentially cued prediction task. J Gerontol B Psychol Sci Soc Sci 69(5):686-694. doi:10.1093/geronb/gbt057 | Outcome is performance on sequentially cued prediction task, decision-making outcomes not quantifiable as positive or negative. |
| Seaman KL, Leong JK, Wu CC, Knutson B, Samanez-Larkin GR (2017) Individual differences in skewed financial risk-taking across the adult life span. Cogn Affect Behav Neurosci 17(6):1232-1241. doi:10.3758/s13415-017-0545-5 | On acceptability of positively versus negatively skewed risk, decision-making outcomes not quantifiable as positive or negative. |
| Seaman KL, Stillman CM, Howard DV, Howard Jr JH (2015) Risky decision-making is associated with residential choice in healthy older adults. Front Psychol 6:1192. doi:10.3389/fpsyg.2015.01192 | Comparison groups are independent versus older adults in retirement housing, no report of performance by age, or age comparison. |
| Segerstrom SC, Geiger PJ, Combs HL, Boggero IA (2016) Time perspective and social preference in older and younger adults: effects of self-regulatory fatigue. Psychol Aging 31(6):594-604. doi:10.1037/pag0000104 | Outcome was preference of partner type (knowledgeable versus close), decision-making outcomes not quantifiable as positive or negative. |
| Shimoyama I, Yoshida A, Yugeta T, Saeki N, Hayashi F, Yoshizaki H, Shimizu R (2012) Right-Left-Discrimination: cognitive reaction time and ageing. International Medical Journal 19(2):106-108. | Outcome is reaction time, decision-making outcomes not quantifiable as positive or negative. |
| Smart CM, Krawitz A (2015) The impact of subjective cognitive decline on Iowa Gambling Task performance. Neuropsychology 29(6):971-987. doi:10.1037/neu0000204 | Comparison of decision-making between adults’ subjective cognitive decision versus adults with no subjective cognitive decline, but no report on age comparison. |
| Spaniol J, Bayen UJ (2005) Ageing and conditional probability judgments: a global matching approach. Psychol Aging 20(1):165-181. doi:10.1037/0882-7974.20.1.165 | On frequency judgement and probability judgements, decision-making outcomes not quantifiable as positive or negative. |
| Spaniol J, Wegier P (2012) Decisions from experience: adaptive information search and choice in younger and older adults. Front Neurosci 6:36. doi:10.3389/fnins.2012.00036 | Outcome is adaptive risk taking, decision-making outcomes not quantifiable as positive or negative. |
| Starns JJ, Ratcliff R (2010) The effects of ageing on the speed–accuracy compromise: boundary optimality in the diffusion model. Psychol Aging 25(2):377. | Outcome is speed-accuracy trade-off, decision-making outcomes not quantifiable as positive or negative. |
| Starns JJ, Ratcliff R (2012) Age-related differences in diffusion model boundary optimality with both trial-limited and time-limited tasks. Psychon Bull Rev 19(1):139-145. doi:10.3758/s13423-011-0189-3 | Outcome was optimal speed-accuracy trade-off, decision-making outcomes not quantifiable as positive or negative. |
| Stormer V, Eppinger B, Li SC (2014) Reward speeds up and increases consistency of visual selective attention: a lifespan comparison. Cogn Affect Behav Neurosci 14(2):659-671. doi:10.3758/s13415-014-0273-z | Outcome is selective attention, decision-making outcomes not quantifiable as positive or negative. |
| Strough J, de Bruin WB, Peters E (2015) New perspectives for motivating better decisions in older adults. Front Psychol 6:783. doi:10.3389/fpsyg.2015.00783 | Review. |
| Tentori K, Osherson D, Hasher L, May C (2001) Wisdom and ageing: irrational preferences in college students but not older adults. Cognition 81(3):B87-B96. | Outcomes are regularity of decision making, decision-making outcomes not quantifiable as positive or negative. |
| Valsecchi M, Billino J, Gegenfurtner KR (2018) Healthy ageing is associated with decreased risk-taking in motor decision-making. J Exp Psychol Hum Percept Perform 44(1):154-167. doi:10.1037/xhp0000436 | On motor variability between older and younger adults, decision-making outcomes not quantifiable as positive or negative. |
| Van Duijvenvoorde AC, Jansen BR, Bredman JC, Huizenga HM (2012) Age-related changes in decision making comparing informed and noninformed situations. Dev Psychol 48(1):192-203. doi:10.1037/a0025601 | Developmental sample which does not contain older adults. |
| Verhaegen C, Poncelet M (2013) Changes in naming and semantic abilities with ageing from 50 to 90 years. J Int Neuropsychol Soc 19(2):119-126. doi:10.1017/S1355617712001178 | Outcome is naming difficulties, decision-making outcomes not quantifiable as positive or negative. |
| Vu KP, Proctor RW (2008) Age differences in response selection for pure and mixed stimulus-response mappings and tasks. Acta Psychol (Amst) 129(1):49-60. doi:10.1016/j.actpsy.2008.04.006 | Outcome is mapping response, decision-making outcomes not quantifiable as positive or negative. |
| Wagner GP, Parente MA (2009) Performance of older adults in two versions of the Iowa Gambling Task. Jornal Brasileiro de Psiquiatria 25(3):425-433. | On role of visual reinforcement and attentional resources processing on risky decision-making, only older adults, no age comparison. |
| Ward EV, Dhami MK (2016) The Ageing Decision-Maker: advances in understanding the impact of cognitive change on decision-making. Front Psychol 7:1622. doi:10.3389/fpsyg.2016.01622 | Editorial. |
| Watanabe S, Shibutani H (2010) Ageing and decision making: differences in susceptibility to the risky-choice framing effect between older and younger adults in Japan. Japanese Psychological Research 52(3):163-174. | Outcome is effect of framing on decision, decision-making outcomes not quantifiable as positive or negative. |
| Wearne TA (2018) Elucidating the Role of the Ventrolateral Prefrontal Cortex in Economic Decision-Making. J Neurosci 38(17):4059-4061. doi:10.1523/JNEUROSCI.0330-18.2018 | Comments on an article. |
| Weierich MR, Kensinger EA, Munnell AH, Sass SA, Dickerson BC, Wright CI, Barrett LF (2010) Older and wiser? An affective science perspective on age-related challenges in financial decision making. Soc Cog Affect Neurosci 6(2):195-206. | Narrative paper. |
| Weller JA, Levin IP, Denburg NL (2011) Trajectory of Risky Decision Making for Potential Gains and Losses from Ages 5 to 85. J Behav Decis Mak 24(4):331-344. doi:10.1002/bdm.690 | Outcome is risk aversion, decision-making outcomes not quantifiable as positive or negative. |
| Westbrook A, Martins BS, Yarkoni T, Braver TS (2012) Strategic insight and age-related goal-neglect influence risky decision-making. Front Neurosci 6:68. doi:10.3389/fnins.2012.00068 | Outcome is the effect of training intervention on risky decision-making but decision-making outcomes not quantifiable as positive or negative. |
| Wiesiolek CC, Foss MP, Diniz PR (2014) Normal ageing and decision-making: a systematic review of the literature of the last 10 years. Journal Brasileiro de Psiquiatria 63(3):255-259. | Review. |
| Woodhead EL, Lynch EB, Edelstein BA (2011) Decisional strategy determines whether frame influences treatment preferences for medical decisions. Psychol Aging 26(2):285-294. doi:10.1037/a0021608 | Outcome is the decision strategy, and second outcome is treatment preference, but neither is better and thus decision-making outcomes not quantifiable as positive or negative. |
| Yoon C, Cole CA, Lee MP (2009) Consumer decision making and ageing: current knowledge and future directions. Journal of Consumer Psychology 19(1):2-16. doi:10.1016/j.jcps.2008.12.002 | Review. |

### Supplementary Table II

### *Sensitivity analysis on the impact of moderators on relationship between age and decision-making*

|  | Model | Effects Size, 95% CI, *I*2 | | Moderators, *p* | | | | | | | | | | | | | |
| --- | --- | --- | --- | --- | --- | --- | --- | --- | --- | --- | --- | --- | --- | --- | --- | --- | --- |
|  |  |  | | Publication year | | | Mean age | | Tasks differences | | + task demands | | IV type | | SES adjustment | | Converted effect size |
| All (*k* = 57) | | | *d* _random_ = -0.17, 95% CI -0.29, -0.04, *I^2^* = 92.92% | | .87 | .55 | | .16 to .99 | | **.03** to .97 | | .15 | | .56 | | .70 | |
| Moderator: intertemporal task demands | | | *d* _random =_ -0.23, 95% CI -0.37, -0.10, *I^2^* = 91.86% | |  |  | |  | |  | |  | |  | |  | |
| Exclusion of intertemporal tasks (*k* = 46) | | | *d* _random_ = -0.24, 95% CI -0.39, -0.09, *I^2^* = 93.38% | |  |  | |  | |  | |  | |  | |  | |
| Moderator: exploration/exploitation | | | *d* _random =_ -0.14, 95% CI -0.26, -0.08, *I^2^* = 92.43% | |  |  | |  | |  | |  | |  | |  | |
| Exclusion of exploration/exploitation tasks (*k* = 53) | | | *d* _random_ = -0.13, 95% CI -0.27, -0.01, *I^2^* = 93.1% | |  |  | |  | |  | |  | |  | |  | |
| Financial *(k =* 36) | | | *d* _random_ = -0.25, 95% CI -0.45, -0.44, *I^2^* = 94.68% | | .26 | .87 | | .50 to .96 | | .15 to .62 | | .29 | | .74 | | .59 | |
| Social *(k =* 14) | | | *d* _random_ = -0.38, 95% CI -0.72, -0.03, *I^2^* = 92.9% | | .25 | .28 | | .54 to .97 | | **.00** to .66 | | **.03** | | .90 | | .82 | |
| Moderator: risky task demands | | | *d* _random =_ -0.26, 95% CI -0.54, 0.25, *I^2^* = 88.80% | |  |  | |  | |  | |  | |  | |  | |
| Exclusion of risky tasks (*k* =13) | | | *d* _random_ = -0.26, 95% CI -0.54, 0.25, *I^2^* = 92.84% | |  |  | |  | |  | |  | |  | |  | |
| Moderator: IV type | | | *d* _random =_ 0.14, 95% CI -0.42, 0.69, *I^2^* = 90.02% | |  |  | |  | |  | |  | |  | |  | |
| Exclusion of continuous IV (*k* = 10) | | | *d* _random_ = -0.59, 95% CI -1.01, -0.18, *I^2^* = 88.5% | |  |  | |  | |  | |  | |  | |  | |
| Health *(k = 5*) | | | *k* = 5, *d* _random_ = 0.34, 95% CI -0.31, 0.99, *I^2^* = 95.02% | | 0.29 | . | | . | | .47 to .83 | | .57 | | . | | .26 | |

### Supplementary Table III

### *Proportion of studies with low, moderate, and high risk of bias in five assessed domains*

| Bias rating | Selection bias | Confounding bias | Attrition bias | Statistical Analysis | Study design |
| --- | --- | --- | --- | --- | --- |
| Low | 47.1% | 71.3% | 1.1% | 40.8% | 20.1% |
| Moderate | 21.3% | 16.1% | 98.8% | 30.5% | 43.1% |
| High | 31.6% | 12.6% | 0.0% | 28.7% | 36.8% |
| ^a^Each cell represented the proportion of items rated low, moderate, or high bias per bias domain. | | | | | |

### Supplementary Table IV

### *Risk of bias for included studies assessed by 11-Item quality rating tool*

| Reference | Selection bias | | | |  | Confounding bias | |  | Attrition bias |  | Statistical analyses | |  | Study design | |
| --- | --- | --- | --- | --- | --- | --- | --- | --- | --- | --- | --- | --- | --- | --- | --- |
|  | 1. Between group age ranges comparable? | 2. Comparable between group sample sizes? | 3. Females/males represented equally? | 4. Sample size considered? |  | 5. Consistency of task administration? | 6. Participant screening conducted? |  | 7. Attrition addressed? |  | 8. Appropriate analysis undertaken? | 9. Statistical control for confounders? |  | 10. Suitable for investigating age differences? | 11. Clearly described? |
| Bailey, Ruffman and Rendell 2013 | Mod | Low | High | High |  | Low | Low |  | Mod |  | Low | Mod |  | Mod | Mod |
| Bakos et al 2008 | Low | Low | High | High |  | Low | Low |  | Mod |  | Mod | High |  | High | Low |
| Bangma et al 2017 | Low | Low | Mod | High |  | Low | Low |  | Mod |  | Low | Mod |  | Mod | Mod |
| Bauer et al 2013  (Denburg 2009 sample) | Low | Low | High | High |  | Low | Low |  | Mod |  | Low | Mod |  | Mod | Mod |
| Beadle et al 2012 | Low | Low | Mod | High |  | Low | High |  | Mod |  | Low | High |  | High | Mod |
| Besedes et al 2012b | Low | Low | High | High |  | Low | High |  | Mod |  | Mod | High |  | High | Mod |
| Best and Freund 2018 | Low | Low | Low | High |  | Mod | High |  | Mod |  | Mod | Low |  | High | Mod |
| Blanco et al 2016 (1) | Mod | Low | Mod | High |  | Low | Low |  | Mod |  | Mod | High |  | High | Mod |
| Blanco et al 2016 (2) | Low | Low | High | High |  | Low | Mod |  | Mod |  | Mod | High |  | High | Mod |
| Boyle et al 2012 (1)  (RUSH sample) | Low | Low | Mod | High |  | Mod | Low |  | Mod |  | Mod | High |  | High | Low |
| Boyle et al 2012 (2)  (RUSH sample) | Low | Low | Mod | High |  | Mod | Low |  | Mod |  | Low | Low |  | Mod | Mod |
| Brand and Schiebener 2013 | Low | Low | Low | High |  | Low | Low |  | Mod |  | Low | Mod |  | Mod | Mod |
| Bruine de Bruin, Strough and Parker 2014 | Low | Low | Low | High |  | Low | Low |  | Mod |  | Mod | Mod |  | High | Low |
| Caird et al 2005 | Mod | Low | Mod | High |  | Low | Mod |  | Mod |  | Low | Low |  | Mod | Mod |
| Carvalho et al 2012 | Low | Low | Mod | High |  | Low | Low |  | Mod |  | Mod | High |  | High | Low |
| Cassimiro et al 2017 | Low | Low | High | High |  | Low | Low |  | Mod |  | Mod | High |  | High | Low |
| Cavanagh et al 2012 | High | Low | Mod | High |  | Low | Low |  | Mod |  | Low | High |  | High | Low |
| Chung, Tymula and Glimcher 2017 | Low | Low | Mod | High |  | Low | Low |  | Mod |  | Mod | High |  | High | Low |
| Cooper et al 2017 (1) | Mod | Low | High | High |  | Low | Mod |  | Mod |  | Low | High |  | High | High |
| Cooper et al 2017 (2) | Mod | Low | High | Mod |  | Low | High |  | Mod |  | Low | High |  | High | High |
| Cooper et al 2013 | High | High | High | High |  | Low | Low |  | Mod |  | Low | High |  | High | High |
| Cooper, Worthy and Maddox 2016 | Mod | Mod | High | High |  | Low | Mod |  | Mod |  | Low | High |  | High | High |
| Deakin et al 2004 | Low | Low | Mod | High |  | Low | High |  | Mod |  | Low | Mod |  | High | High |
| Denburg et al 2009 | Low | Low | Mod | High |  | Low | Mod |  | Mod |  | Low | Mod |  | Mod | Mod |
| Denburg et al 2007 (1) | Low | Low | Low | High |  | Low | Low |  | Mod |  | Mod | High |  | High | Mod |
| Denburg et al 2007 (2)  (Includes Denburg et al 2007 (1) sample) | High | Low | High | High |  | Low | Low |  | Mod |  | Mod | High |  | High | Mod |
| Denburg, Travel and Bechara 2005 | Low | Low | Low | High |  | Low | Low |  | Mod |  | Mod | High |  | High | Low |
| Di Rosa et al 2017 | Low | Low | Mod | High |  | Low | Low |  | Mod |  | Mod | High |  | High | Mod |
| Eberhardt, de Bruin and Strough 2019 | Low | Low | Low | Mod |  | Low | High |  | Low |  | Mod | Low |  | High | Low |
| Eppinger, Heekeren and Li 2017 | High | Low | Low | High |  | Low | Low |  | Mod |  | Low | High |  | High | Mod |
| Fein, McGillivray and Finn 2007 | Low | High | Mod | High |  | Low | Low |  | Mod |  | Low | Mod |  | Mod | Low |
| Fernandes et al 2019 | Low | Low | High | High |  | Low | Low |  | Mod |  | Mod | High |  | High | Low |
| Girardi, Sala and MacPherson 2018 | Mod | Low | High | Mod |  | Low | Low |  | Mod |  | Mod | High |  | High | Mod |
| Harle and Sanfey 2012 | Mod | Low | Mod | High |  | Low | Mod |  | Mod |  | Low | High |  | High | Mod |
| Henninger, Madden and Huettel 2010 | Low | Low | Low | Mod |  | Low | High |  | Mod |  | Mod | High |  | High | High |
| Hess et al 2018 (1) | Low | Low | Low | High |  | Low | High |  | Mod |  | Low | Mod |  | Mod | Mod |
| Hess et al 2018 (2) | Low | Low | Low | High |  | Low | High |  | Mod |  | Low | High |  | High | Mod |
| Hess, Queen and Patterson 2012 | Low | Low | High | High |  | Low | High |  | Mod |  | Mod | High |  | High | Mod |
| Huang et al 2015 | Mod | Low | Mod | Low |  | Low | Low |  | Mod |  | Low | Mod |  | Mod | Low |
| James et al 2012  (RUSH sample) | Low | Low | Mod | High |  | Low | Mod |  | Mod |  | Low | Mod |  | Mod | Low |
| Jimura et al 2011 | High | Low | High | High |  | Low | Mod |  | Mod |  | Low | High |  | High | Mod |
| Koscienlniak 2016 | Mod | Low | High | High |  | Low | Low |  | Mod |  | Low | High |  | High | Low |
| Li et al 2017 | Low | Mod | Mod | Low |  | Low | Low |  | Mod |  | Low | Mod |  | Mod | Low |
| Li et al 2013 | Mod | Mod | Mod | Mod |  | Low | Mod |  | Mod |  | Mod | High |  | Low | Mod |
| Li et al 2015 | Low | Low | Mod | Mod |  | Low | Mod |  | Mod |  | Mod | High |  | Low | Mod |
| Lobjos and Cavallo 2007 | Low | Low | Low | High |  | Low | Mod |  | Mod |  | Low | High |  | High | Mod |
| Lockenhoff et al 2016 (1) | High | Low | Low | High |  | Low | Low |  | Mod |  | Low | Low |  | Mod | Low |
| Lockenhoff et al 2016 (2) | Low | Low | Low | High |  | Low | Low |  | Mod |  | Low | Low |  | Mod | Low |
| Ma and Chen 2015 | High | Low | Mod | High |  | Low | High |  | Mod |  | Mod | High |  | High | Mod |
| MacPherson, Phillips and Sala 2002 | Low | Low | Low | High |  | Low | Low |  | Mod |  | Low | Mod |  | Mod | Low |
| Mata, Schooler and Rieskamp 2007 | Low | Low | Mod | High |  | Low | Mod |  | Mod |  | Low | Mod |  | Mod | Low |
| Mata, von Helversen and Rieskamp 2010 | Low | Low | Mod | High |  | Low | High |  | Mod |  | Low | High |  | High | Mod |
| Mikels et al 2013 | Low | Low | Mod | High |  | Low | High |  | Mod |  | Low | Mod |  | Mod | Mod |
| Mikels et al 2010 | Low | Low | Low | High |  | Low | Low |  | Mod |  | Low | Mod |  | Mod | Low |
| Pachur, Mata and Hertwig 2017 | Mod | Low | Mod | High |  | Low | High |  | Mod |  | Low | Mod |  | Mod | High |
| Pachur et al 2009 (1) | Low | Low | Mod | High |  | Low | Mod |  | Mod |  | Low | Low |  | Mod | Low |
| Pachur et al 2009 (2) | Low | Low | Low | High |  | Low | Mod |  | Mod |  | Low | High |  | High | Low |
| Pertl et al 2017 | Low | Low | High | High |  | Low | Low |  | Mod |  | Mod | High |  | High | Mod |
| Queen and Hess 2010 | Mod | Mod | Low | High |  | Low | High |  | Mod |  | Mod | High |  | High | Low |
| Rieger and Mata 2013 | Low | Low | High | High |  | Low | Mod |  | Mod |  | Low | Low |  | Mod | Low |
| Roalf et al 2012 | Low | Low | Low | High |  | Low | Mod |  | Mod |  | Low | Mod |  | Mod | Mod |
| Rogalsky et al 2012 | Low | Low | Mod | High |  | Low | Mod |  | Mod |  | Mod | High |  | High | Mod |
| Rolison, Hanoch and Wood 2012 | Low | Low | High | High |  | Low | Mod |  | Mod |  | Low | High |  | High | Mod |
| Rolison, Wood and Hanoch 2017 (1) | Low | Low | Mod | Mod |  | Low | Low |  | Mod |  | Low | Mod |  | Mod | Low |
| Rolison, Wood and Hanoch 2017 (2) | Low | Low | Mod | Mod |  | Low | Low |  | Mod |  | Low | Mod |  | Mod | Low |
| Rosen, Brand and Kalbe 2016 | Low | Low | Mod | Low |  | Low | Low |  | Mod |  | Mod | Low |  | Mod | Low |
| Rutledge et al 2016 | Low | Low | High | Mod |  | Low | High |  | Mod |  | Low | Mod |  | High | Mod |
| Rydzewska et al 2018 (1) | Low | Low | Mod | High |  | Low | High |  | Mod |  | Low | Mod |  | Mod | Mod |
| Rydzewska et al 2018 (2) | Low | Low | Mod | High |  | Low | High |  | Mod |  | Low | Mod |  | Mod | Mod |
| Schiebener and Brand 2017 (Includes Brand and Schiebener 2013 sample) | Low | Low | Mod | Mod |  | High | Mod |  | Mod |  | Mod | Mod |  | Mod | High |
| Seaman et al 2018 | Low | Low | Mod | High |  | Low | Mod |  | Mod |  | Low | High |  | High | High |
| Seaman et al 2016 | Low | Low | Mod | High |  | Low | Mod |  | Mod |  | Low | Low |  | Mod | Mod |
| Sparrow and Spaniol 2018 (1) | Low | Low | Mod | High |  | Low | Low |  | Mod |  | Mod | High |  | High | Low |
| Sparrow and Spaniol 2018 (2) | High | Mod | Mod | High |  | Low | Low |  | Mod |  | Mod | High |  | High | Low |
| Stewart et al 2018  (RUSH Sample) | Low | Low | Mod | High |  | Low | Low |  | Mod |  | Low | Low |  | Mod | Low |
| Strough et al. 2016 | Low | Low | Low | High |  | Low | Low |  | Mod |  | Low | Low |  | High | Low |
| Tymula et al 2013 | High | Low | Low | High |  | Low | Low |  | Mod |  | Low | Low |  | Mod | Low |
| Von Helversen and Mata 2012 | Low | Low | Mod | High |  | Low | High |  | Mod |  | Low | High |  | High | Mod |
| Walker, Fisk and Mcguire 1997 | Low | Low | High | High |  | Low | Low |  | Mod |  | Low | High |  | High | Mod |
| Wayde et al 2017 | Mod | Low | High | High |  | Low | Mod |  | Mod |  | Low | High |  | High | Mod |
| Wood et al 2005 | Low | Mod | High | High |  | Low | High |  | Mod |  | Low | High |  | High | Mod |
| Wood et al 2011 | High | Low | Mod | Low |  | Low | Mod |  | Mod |  | Low | Low |  | Mod | Mod |
| Worthy et al 2014 | High | Low | High | High |  | Low | High |  | Mod |  | Low | High |  | High | Mod |
| Worthy et al 2016 | Low | Low | Mod | High |  | Low | Low |  | Mod |  | Low | High |  | High | Mod |
| Worthy et al 2011 (1) | Low | Low | Mod | High |  | Low | Mod |  | Mod |  | High | High |  | High | High |
| Worthy et al 2011 (2) | Mod | Low | High | High |  | Low | Mod |  | Mod |  | Low | High |  | High | High |
| Zamarian et al 2008 | Low | Mod | Mod | Low |  | Low | Low |  | Mod |  | Low | High |  | High | Low |

^a^Mod, Moderate. For interrater consistency, the items were rated using the following guidance:

Item 1. Between group differences in age range: <10 years = Low, 10-15 years = Mod, >15 year = High. Rate as low if no between group comparison made.

Item 2. Between group differences in sample size: <10 = Low, 11-25 =Mod, >25 = High. Rate as low if no between group comparison made.

Item 3. Overall proportion of females: 45%-55% = Low, 56%-80% = Moderate, >80% = High.

Item 4. Consideration of sample size: presence of power calculation = Low, sample size considered but not estimated (e.g., based on previous literature) = Mod, no mention = High.

Item 5. Consistency of task administration: consider description and methods e.g., computerised tasks, multiple experimenters.

Item 6. Participant screening: well described and replicable = Low, present but difficult to replicate = Mod, no screening conducted/mentioned = High.

Item 7. Attrition: unlikely and addressed = Low, possible but not addressed = Mod, likely but not described (e.g., in longitudinal studies) = High.

Item 8. Statistical analysis: was robust and reported (e.g., ANOVA, t-test) = Low, conducted but not well described = Mod, not robust/described =High

Item 9. Confounders: considered and controlled for statistically (e.g., covariates included) = Low, some consideration but reporting unclear = Mod, not considered/statistically controlled for = High.

Item 10. Study design: suited to assess intraindividual change (i.e., longitudinal) = Low, suited to assess interindividual change accounting for cohort effects (i.e., cross-sectional and considered cohort effects), suited to assess interindividual change not accounting for cohort effects = High.

Item 11. Study description: detailed and replicable = Low, lacking some detail = Moderate, lacking considerable detail = High

### Supplementary Table VI

### *PRISMA Checklist*

| Section/topic | # | Checklist item | Reported on page # |
| --- | --- | --- | --- |
| **Title** | 1 | Identify the report as a systematic review, meta-analysis, or both. | 1 |
| **Abstract** | | |  |
| Structured summary | 2 | Provide a structured summary including, as applicable: background; objectives; data sources; study eligibility criteria, participants, and interventions; study appraisal and synthesis methods; results; limitations; conclusions and implications of key findings; systematic review registration number. | 2 |
| **Introduction** | | |  |
| Rationale | 3 | Describe the rationale for the review in the context of what is already known. | 3-6 |
| Objectives | 4 | Provide an explicit statement of questions being addressed with reference to participants, interventions, comparisons, outcomes, and study design (PICOS). | 5-6 |
| **Methods** | | |  |
| Protocol and registration | 5 | Indicate if a review protocol exists, if and where it can be accessed (e.g., Web address), and, if available, provide registration information including registration number. | 6 |
| Eligibility criteria | 6 | Specify study characteristics (e.g., PICOS, length of follow-up) and report characteristics (e.g., years considered, language, publication status) used as criteria for eligibility, giving rationale. | 7 |
| Information sources | 7 | Describe all information sources (e.g., databases with dates of coverage, contact with study authors to identify additional studies) in the search and date last searched. | 6 |
| Search | 8 | Present full electronic search strategy for at least one database, including any limits used, such that it could be repeated. | 6 |
| Study selection | 9 | State the process for selecting studies (i.e., screening, eligibility, included in systematic review, and, if applicable, included in the meta-analysis). | 6 |
| Data collection process | 10 | Describe method of data extraction from reports (e.g., piloted forms, independently, in duplicate) and any processes for obtaining and confirming data from investigators. | 8-10 |
| Data items | 11 | List and define all variables for which data were sought (e.g., PICOS, funding sources) and any assumptions and simplifications made. | 8-10 |
| Risk of bias in individual studies | 12 | Describe methods used for assessing risk of bias of individual studies (including specification of whether this was done at the study or outcome level), and how this information is to be used in any data synthesis. | 9 |
| Summary measures | 13 | State the principal summary measures (e.g., risk ratio, difference in means). | 10-11 |
| Synthesis of results | 14 | Describe the methods of handling data and combining results of studies, if done, including measures of consistency (e.g., *I*^2^) for each meta-analysis. | 11 |
| Risk of bias across studies | 15 | Specify any assessment of risk of bias that may affect the cumulative evidence (e.g., publication bias, selective reporting within studies). | 8 |
| Additional analyses | 16 | Describe methods of additional analyses (e.g., sensitivity or subgroup analyses, meta-regression), if done, indicating which were pre-specified. | 10 |
| **Results** | | | |
| Study selection | 17 | Give numbers of studies screened, assessed for eligibility, and included in the review, with reasons for exclusions at each stage, ideally with a flow diagram. | 10 |
| Study characteristics | 18 | For each study, present characteristics for which data were extracted (e.g., study size, PICOS, follow-up period) and provide the citations. | 10-11 |
| Risk of bias within studies | 19 | Present data on risk of bias of each study and, if available, any outcome level assessment (see item 12). | 10 |
| Results of individual studies | 20 | For all outcomes considered (benefits or harms), present, for each study: (a) simple summary data for each intervention group (b) effect estimates and confidence intervals, ideally with a forest plot. | 11 |
| Synthesis of results | 21 | Present results of each meta-analysis done, including confidence intervals and measures of consistency. |  |
| Risk of bias across studies | 22 | Present results of any assessment of risk of bias across studies (see Item 15). | 13 |
| Additional analysis | 23 | Give results of additional analyses, if done (e.g., sensitivity or subgroup analyses, meta-regression [see Item 16]). | 12-13 |
| **Discussion** | | | |
| Summary of evidence | 24 | Summarise the main findings including the strength of evidence for each main outcome; consider their relevance to key groups (e.g., healthcare providers, users, and policy makers). | 13-16 |
| Limitations | 25 | Discuss limitations at study and outcome level (e.g., risk of bias), and at review-level (e.g., incomplete retrieval of identified research, reporting bias). | 16-17 |
| Conclusions | 26 | Provide a general interpretation of the results in the context of other evidence, and implications for future research. | 13-17 |
| **Funding** | | | |
| Funding | 27 | Describe sources of funding for the systematic review and other support (e.g., supply of data); role of funders for the systematic review. | 17 |

*Note.* Checklist from Moher D, Liberati A, Tetzlaff J, Altman DG (2009) Preferred Reporting Items for Systematic Reviews and Meta-Analyses: the PRISMA statement. PLoS Med 6(7):e1000097. doi:10.1371/journal.pmed1000097

### Supplementary Table VII

### *Descriptions of key decision-making tasks*

| Tasks | Reference | Description |
| --- | --- | --- |
| Iowa Gambling Task (IGT) | Bechara A, Damasio AR, Damasio H, Anderson SW (1994) Insensitivity to future consequences following damage to human prefrontal cortex. Cognition 50:7-15. | The IGT is a card task which requires participant to choose 100 cards from four decks (A, B, C, D). The participant starts with $2000 of game credit and told to make a profit. They win or lose money with each card selection and are not aware of what each card would yield in advance. There is a 50% chance of winning or losing on each turn. Each card selection from Decks A and B either yields $100 or deducts $250 (higher risk) and each selection from Decks C and D either yields $50 or deducts $50 (low risk). The IGT assesses aversion to risk, and how long it takes people to decide before making a risky decision. |
| Ultimatum Game (UG) | Güth W, Schmittberger R, Schwarze B (1982) An experimental analysis of ultimatum bargaining. J Econ Behav Organ 3(4):367-388. | The UG is a two-player behavioural economics game. A set amount of money is available to be split between two players. The proposer suggests a division of the money between themselves and the responder. This occurs over several trials. If the responder accepts the offer, then both players receive the proposed amounts. If the responder rejects the offer, no one is given any money. Technically it would make sense for the responder to accept any amount offered, however they may reject offers they deem unfair in hopes of attaining a fairer offer. The UG has been used to assess reciprocity and generosity. |
| Game of Dice Task (GDT) | Brand M, Pawlikowski M, Labudda K, Laier C, von Rothkirch N, Markowitsch HJ (2009) Do amnesic patients with Korsakoff’s syndrome use feedback when making decisions under risky conditions? An experimental investigation with the Game of Dice Task with and without feedback. Brain Cogn 69(2):279-290. | The GDT is a computerised tasks which requires participants to predict the outcome of a dice roll. Participants are told to maximise their winnings and select options ranging from high probability but low monetary payoff to low probability but high monetary payoff. On 18 trials they select one, two, three or four dice in attempt to predict the result on the rolled dice. Selecting lower number of dice has the potential to yield higher pay offs and vice versa. Probabilities of wins and losses are made explicit on the screen. The GDT assesses participants' aversion/attraction to risky decision. |
| Balloon Analogue Risk Task (BART) | Lejuez CW, Read JP, Kahler CW, Richards JB, Ramsey SE, Stuart GL, Strong DR, Brown RA (2002) Evaluation of a behavioral measure of risk taking: the balloon analogue risk task (BART). J Exp Psychol App 8(2):75. | The BART is a computerised decision-making task that assesses risk-taking behaviour. Each participant is presented with 90 balloons of 3 different colours which appear one at a time. Participants click a button to inflate the balloon and accumulate 5 cents per pump into a temporary bank which they are unable to view. They may also choose to collect their winnings which are transferred into a permanent bank. If the balloon explodes the temporary bank resets to zero, and another balloon appears. If they collect their winnings before the balloon explodes, they will be able to see the amount earned on that balloon. |

^a^This table details the original versions of each task, but some studies have since used adapted versions.
